# Proof-of-principle neural network models for classification, attribution, creation, style-mixing, and morphing of image data for genetic conditions

**DOI:** 10.1101/2021.04.08.21255123

**Authors:** Dat Duong, Rebekah L. Waikel, Ping Hu, Cedrik Tekendo-Ngongang, Benjamin D. Solomon

**Author notes:** Corresponding author: Benjamin D. Solomon, MD, Clinical Director, National Human Genome Research Institute, Building 10, Suite 3-2551, 10 Center Drive, Bethesda, MD 20892, United States of America.

## Abstract

Neural networks have shown strong potential to aid the practice of healthcare. Mainly due to the need for large datasets, these applications have focused on common medical conditions, where much more data is typically available. Leveraging publicly available data, we trained a neural network classifier on images of rare genetic conditions with skin findings. We used approximately100 images per condition to classify 6 different genetic conditions. Unlike other work related to these types of images, we analyzed both preprocessed images that were cropped to show only the skin lesions, as well as more complex images showing features such as the entire body segment, patient, and/or the background. The classifier construction process included attribution methods to visualize which pixels were most important for computer-based classification. Our classifier was significantly more accurate than pediatricians or medical geneticists for both types of images. Next, we trained two generative adversarial networks to generate new images. The first involved all of the genetic conditions and was used for style-mixing to demonstrate how the diversity of small datasets can be increased. The second focused on different disease stages for one condition and depicted how morphing can illustrate the disease progression of this condition. Overall, our findings show how computational techniques can be applied in multiple ways to small datasets to enhance the study of rare genetic diseases.

## Introduction

Neural network models have demonstrated strong potential to improve the practice of healthcare. For example, “artificial intelligence” may help detect breast cancer via mammogram analysis or COVID-19 based on CT scans.^1,2^ In this type of computer vision approach, because medical datasets are typically small compared to other types of publicly available datasets, the neural network is first pretrained on a large, general dataset (e.g. ImageNet, http://www.image-net.org/) to help identify major features, such as edges or basic shapes. Next, the neural network is fine-tuned to address a more specific question, such as the ability to recognize certain diseases. For this approach to work well, the pretrained data must either be similar in type to the medical data or the size of the medical dataset must be relatively large.^3^

Due to these limitations, neural network applications in healthcare have focused on relatively common conditions, where sufficiently large datasets are more readily collected. Genetic conditions, though common in aggregate, are largely individually rare.^4^ A recent meta-analysis identified 82 studies comparing deep learning performance to that of health care professionals in disease detection using medical imaging. None of the conditions in this meta-analysis were genetic, though some (e.g., breast cancer) involve clear genetic underpinnings in a minority of patients.^5^

Despite this lack of representation, neural network approaches have been used in some areas of genetics.^6^ With efforts to collect adequate training data, these methods could be especially useful in clinical genetics, where there is a lack of trained individuals to help determine whether a patient may be affected by a genetic condition, what that condition may be, and what testing strategy and next management steps are indicated.^7,8^ With the accelerating expansion of genomics into diverse fields of medicine,^9^ an alternative strategy of training non-geneticist clinicians has not kept pace.^10^ Developing computational methods could help geneticists as well as other clinicians manage the large numbers of affected individuals.

To explore the use of these techniques in proof-of-principle exercises using small datasets, we collected images of a selected group of rare genetic conditions that manifest with characteristic skin findings. We chose clinically impactful conditions that can be nontrivial to diagnose. We built neural network classifiers both for images that were cropped to focus on the lesions of interests, similar to previous studies^11,12^ as well as uncropped images. These uncropped images can be more difficult for a neural network model to analyze, but may more closely mimic real-life, unprocessed images such as those a clinician might encounter in training or diagnostic situations. During the classifier construction process, we used an attribution method for image recognition to help our team visualize how the classifier weighted certain pixels in the image. We then compared the classifier performance to pediatricians and medical geneticists.

Beyond neural network-based classification, we trained generative adversarial networks (GAN) to demonstrate how style-mixing can generate more ancestrally diverse images and how morphing images can demonstrate the progression of NF1 manifestations. In using GAN, we empirically addressed two issues. First, although a few GAN models have been built for training smaller datasets, the dataset we used is approximately an order of magnitude smaller than those previously described.^13^ Second, the images that we used, which represent unique, unrelated patients at different disease stages, are heterogeneous, which is challenging for recreating realistic time series.

To summarize, our contributions include: 1) evaluation of a neural network classifier’s performance on a small dataset of esoteric genetic conditions; 2) comparison of how focused and panoramic images affect human and the neural network model’s accuracy; 3) new image generation to augment datasets and to show the progression of a selected condition.

## Materials and Methods

### Ethics review

The study was reviewed by National Human Genome Research Institute (NHGRI) bioethicists and the National Institutes of Health (NIH) Institutional Review Board (IRB). The main analyses were considered not human subjects research; a waiver of consent was granted by the NIH IRB for the work (NIH protocol: 000285) involving the surveys of medical professionals, as described below.

### Data collection

Using condition and gene names, we searched Google and PubMed to identify publicly available images showing the following conditions: Hypomelanosis of Ito (HMI), Incontinentia Pigmenti (IP), McCune-Albright Syndrome (MA), Neurofibromatosis Type 1 (NF1), Noonan Syndrome with Multiple Lentigines (ML; formally known as LEOPARD syndrome), and Tuberous sclerosis Complex (TSC) (see Supplemental Table 1 for more details on these conditions). We also selected images of other medical conditions (e.g., erythema migrans, hemangioma, etc.) that are unrelated to the genetic conditions, but which could be misclassified as one of the genetic conditions. Though difficult to quantify due to lack of available, comprehensive information for many images, we endeavored to collect images from individuals of diverse racial and ethnic backgrounds. This was done by manually reviewing images as well as ascertaining images from sources that focus on ancestrally diverse individuals (such as journals devoted to the presentation of medical conditions in diverse geographic locations).

As defined below (see the “Initial image processing” section), the images we used included focused images (N=1033 (total): 97 HMI, 115 IP, 123 MA, 102 ML, 230 NF1, 122 TSC, 244 other) and panoramic images (N=802 (total): 90 HMI, 84 IP, 103 MA, 79 ML, 120 NF1, 91 TSC, 235 other). Two board-certified clinicians (one medical geneticist and one genetic counselor) reviewed images and data in the source websites to help ensure accuracy of diagnoses based on clinical descriptions and the information described. For example, if a publication showed an image of a patient described as having NF1, our team reviewed the image and the description of the patient to ensure that the patient had strong evidence for the diagnosis, and that there was not contradictory evidence, such as a statement that the patient, on ultimate genetic testing, did not have findings consistent with the suspected diagnosis. All images and URLs used in classification are listed in Supplemental Table 2.

### Initial image processing

First, following the conventions of other large-scale neural network studies on images of skin cancer,^11,12,14^ we cropped images to contain just the lesions of interest. We refer to this dataset as *focused images*. For conditions with multiple skin or external findings, we focused on one main manifestation per condition. For example, for NF1, we focused on café-au-lait macules (CALMs) rather than cutaneous neurofibromas.^15^ We also used the photos as they were captured, some of which show an entire person’s face or body segment (e.g, an arm or the entire back) with the genetic conditions, and with other features such as clothes or a background, though we cropped out words, such as a heading that indicated the image number. We refer to this second dataset as *panoramic images*.

A single panoramic image can have multiple corresponding focused images. For example, as shown in Figure 1, an image of a person with NF1 may include multiple CALMs. We did not want the model to capture anything related to the test images during training. Hence, for the test set, from each of the 7 categories, we selected 20 panoramic images and corresponding focused images. Each panoramic test image has exactly one corresponding focused image. In total, the panoramic test set and their corresponding focused test images contained 140 images each. The remaining images were used to train the model.

**Figure 1.**
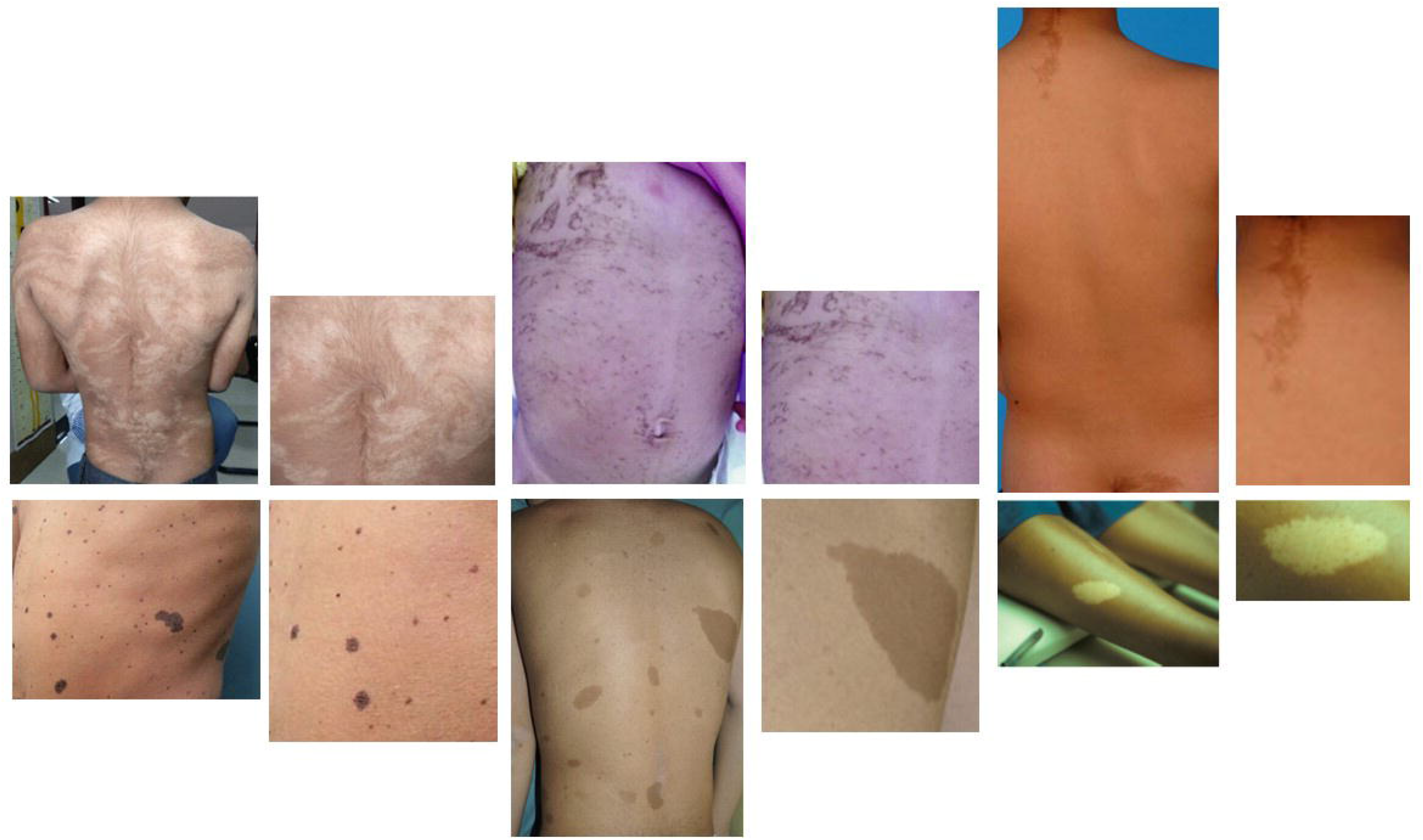
Example panoramic and focused images for each pair of conditions. Top row, from left to right: Hypomelanosis of Ito (HMI), Incontinentia Pigmenti (IP), McCune-Albright Syndrome (MA). Bottom row, from left to right: Neurofibromatosis Type 1 (NF1), Noonan Syndrome with Multiple Lentigines (ML; formally known as LEOPARD syndrome), Tuberous sclerosis Complex (TSC). See Supplemental Table 1 for more details on these conditions. Image sources (all used with appropriate permissions; see Supplemental data for permissions): HMI: https://casereports.bmj.com/content/12/4/e227693; IP: https://www.researchgate.net/figure/X-linked-Incontinentia-pigmenti-Pattern-type-1a-Blaschko-lines-narrow-bands_fig3_257074473; MA: https://www.researchgate.net/figure/Representative-Cafe-au-lait-Spots-Seen-in-McCune-Albright-Syndrome-A-spectrum-of-spots_fig4_225077911; ML: https://pmj.bmj.com/content/94/1116/605; NF1: https://www.sciencedirect.com/science/article/pii/S1578219016300853; TSC: https://www.sciencedirect.com/science/article/pii/B9780444627025000068?via%3Dihub

### Classifier

We chose the EfficientNet-B4 classifier which achieved good performance on the ImageNet data with a relatively low number of parameters.^16^ We initialized EfficientNet-B4 with the parameter values pretrained on ImageNet and continued training the entire model, not just the last few fully-connected layers.^3^ Combining and then jointly training a small dataset of interest with a larger auxiliary dataset often help the prediction accuracy.^17,18^ For the auxiliary dataset, we downloaded the publicly available SIIM-ISIC Melanoma Classification Challenge Dataset from 2018 to 2020.^11,19^ This dataset contains 58,459 images of 9 skin cancer diseases: actinic keratosis, basal cell carcinoma, benign keratosis, dermatofibroma, melanoma, melanocytic nevus, squamous cell carcinoma, vascular lesion, and other unknown skin cancer cases.

We trained our dataset with the SIIM-ISIC dataset where we classified an image as one of the 16 diseases (7 from our genetic + other disorders dataset and 9 from the SIIM-ISIC dataset). We conducted two experiments, the first one with our focused images and SIIM-ISIC images, and the second with our panoramic images and SIIM-ISIC images. Both experiments used 450 by 450 pixel images and the same data augmentation. Weighted loss was used in both image type trainings: focused images were weighted 5 times and panoramic images were weighted 10 times more than the SIIM-ISIC images. Training was done on one 11 GB Nvidia graphic card. Our code is available at: github.com/datduong/ClassifyNF1.

For our focused images, a 5-fold cross-validation was used to build 5 different classifiers (one for each fold). To create an ensemble predictor, we used each classifier to estimate the predicted probabilities for the labels of a test image. The average of these probabilities was calculated for the 5 classifiers. When averaging, we considered only the classifiers that produced a maximum predicted probability (over all the labels) of at least 0.5. The same procedure was used for training the model on our panoramic images. To visualize which parts of an image the classifier considered to be important, we applied Integrated Gradient to identify pixels of an image that most affect the classifier’s outcome.^20^

### Comparison to clinicians

We compared the classifier to board-certified or board-eligible medical geneticist physicians and pediatricians. We chose these specialties because, in our experience, these types of clinicians more frequently encounter these patients (versus, for example, dermatologists, who may more often assess other skin conditions).^21^ That is, a typical path involves an initial encounter by a pediatrician, followed by referral to a medical geneticist, if available.

We generated surveys using Qualtrics (Provo, Utah, United States of America). Each survey has 4 panoramic images and their corresponding focused versions for each of the 7 conditions (6 genetic conditions + other conditions). As there are 140 panoramic test images, it takes 5 surveys to cover all the test images. We created 6 sets of these 5 surveys, so that each test image would be seen 6 times. In each survey, we showed the focused images first, and then the panoramic images. We suspected that participants would more accurately classify panoramic images as these can include more diagnostic clues than a single focused image, and that presenting images in this order could result in more reliable, independent responses.

When responding to the survey, participants were directed not to use any external resources for help, and to select the genetic condition best represented by the image presented. The surveys also included 3 demographic questions (medical specialty, number of years in practice, and location of current practice), but number of years in practice and location were only used for verification purposes, rather than for analyses. See Supplemental materials for a copy of the survey.

Following previous methods,^11,12^ we estimated that 30 participants for each clinician type would provide a statistical power of 95% to detect a 10% difference. For each of the 30 surveys, one board-certified or board-eligible medical geneticist physician and one board-certified or board-eligible pediatrician was recruited via email. To identify survey respondents, we obtained email addresses through professional networks, departmental websites, journal publications, and other web-available lists. A total of 105 medical geneticists were contacted, 37 agreed to participate, and 32 completed the survey. A total of 379 pediatricians were contacted, 37 agreed to participate, and 32 completed the survey. Surveys were considered complete if >95% of the multiple-choice questions were answered. If multiple medical geneticists or pediatricians completed the same survey, only the first complete survey was used for analysis.

### Generative adversarial network

After classification, we used a subset of our dataset, and collected new images of later-stage disease to generate new images and to show how “morphing” could illustrate disease progression. We identified and used 107 early, 71 intermediate, and 103 late-stage NF1 images (see Supplemental Table 3). These were collected, labeled, and processed in the same way as the images for the classifier. We trained StyleGAN2-ada on our skin images of the NF1 stages.^13^ Stages were assigned by our study team (and reviewed by a genetic counselor and medical geneticist) based on patient age and clinical features according to the natural history of disease.^15^ We loaded the model weights pretrained on Flickr-Faces-HQ (FFHQ) dataset, changed the model objective into conditional GAN, where the labels correspond to the three NF1 stages, and then fine-tuned on our NF1 localized images with the same hyperparameters used for FFHQ dataset.^13^ During training, we kept the NF1 images in their original forms and did not apply any preprocessing, such as cropping and centering the images or background blurring, which are often done to FFHQ dataset. We chose not to preprocess in this way, as we wanted to explore a model that could be more readily applied to image sets without this additional step. Moreover, in many medical cases, all parts of an image may be important, and methods must be developed to analyze the skin findings in conditions like HMI or IP, where cropping and centering will not work since the lesions may not be small or discrete.

In this morphing experiment, we considered only focused images. This was because, with our small NF1 dataset, our exploratory work at generating realistic panoramic images did not produce realistic images, since the GAN not only has to generate the skin lesions but also the body parts where the lesions occur. To generate an image, conditional StyleGAN2-ada takes two key inputs: a random vector z and a 1-hot label vector.^13^ The random vector z is responsible for creating a random image from the label specified by the 1-hot vector. For our method, the three 1-hot label vectors are v_1_ = [1, 0, 0], v_2_ = [0, 1, 0] and v_3_ = [0, 0, 1] to denote that the NF1 image is of an early, intermediate or late stage. The 1-hot vector is then multiplied with the label embedding L ∈ R^MxD^, where M is the number of labels and D = 512 is the default setting. In our running example, we have M = 3, and Lv_1_ returns the vector representing the label “NF1 early stage.” We set the vector representing NF1 intermediate stage to be the average of the vectors representing NF1 early and late stages, that is Lv_2_ = 0.5(Lv_1_ + Lv_3_). L is a model parameter trained using our NF1 images. To generate later stages of an early stage NF1 image, we computed a linear interpolation between v_1_ and v_3_, and then passed these interpolated vectors with the same random vector z as inputs to StyleGAN2-ada. For style-mixing, we trained another StyleGAN2-ada to generate realistic images for the different genetic conditions. We also used style-mixing to transfer attributes of images (including skin and lesion colors) from one group of images to another to show how style-mixing can create more images for less represented groups in datasets.^13^ Our code is available at: github.com/datduong/stylegan2-ada-MorphNF1.

## Results

### Classifier

We first assessed the classifiers’ performances when jointly trained on our dataset and SIIM-ISIC dataset. None of our focused and panoramic test images were classified as one of the cancer conditions in SIIM-ISIC dataset. This was expected because (1) SIIM-ISIC diseases are very unrelated to our genetic conditions and (2) the pose-style and patient ethnicity (SIIM-ISIC represents individuals of mostly European descent) in the SIIM-ISIC images are very different from those in our images. The latter reason is especially true when jointly training the classifier on our panoramic images and SIIM-ISIC. When evaluated on the same 30 surveys described above, in “*Comparison to clinicians”*, the classifier trained on focused images and the classifier trained on panoramic images obtain the same accuracy (2-sided paired t-test, p = 1).

To determine how well our classifier performed relative to clinicians’ abilities to identify these conditions, we compared the output to that of medical geneticists and pediatricians. Group results for accuracy for classification are shown in Figure 2A. Overall, the computer classifier performed 24.6% (p= 1.55×10^−12^) and 15.6% (p = 9.27×10^−9^) better than medical geneticists for focused and panoramic images, respectively. The computer classifier performed 40.1% (p = 5.24×10^−14^) and 31.5% (p=1.85×10^−12^) better than pediatricians for focused and panoramic images, respectively.

**Figure 2A.**
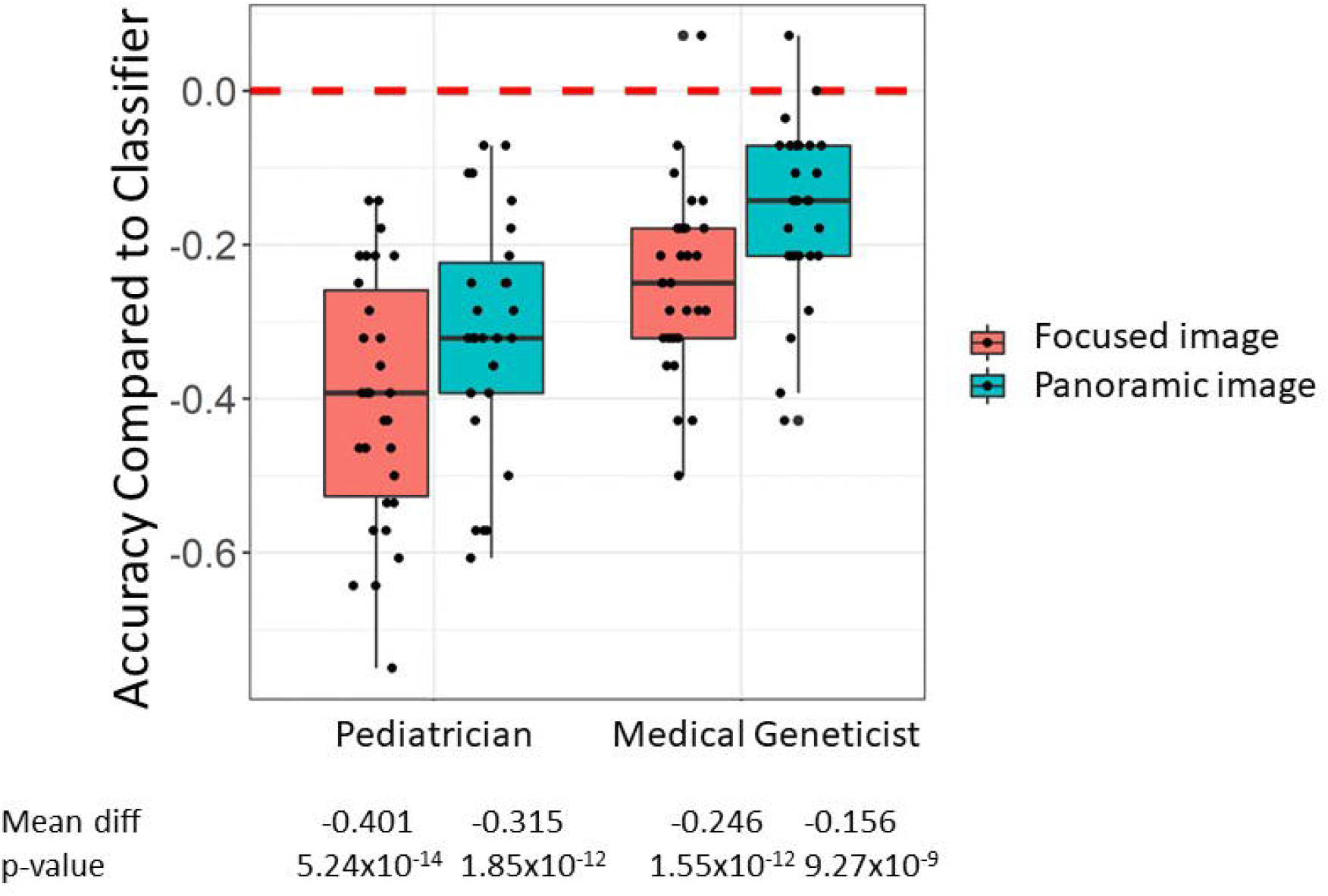
Performance of physicians compared to deep learning classifier. We trained two classifiers, one on focused images and the other on panoramic images. We compared the performance of the classifiers to that of pediatricians and medical geneticists. In the box plots, each point represents the accuracy difference between the classifier and the human performance for a single survey. The red line indicates the baseline accuracy for the classifier.

The results for each of the individual genetic conditions are shown in Figures 2B (focused images) and 2C (panoramic images). Clinicians performed better with panoramic than with focused images, though were on average still less accurate than the classifier. For the panoramic images, the condition with the lowest accuracy (80%) for the classifier was NF1, which was still higher than the average for the physicians (77.5% for medical geneticists and 61.7% for pediatricians). The most difficult condition for both medical geneticists and pediatricians to classify based on panoramic images was MA, with 49.2% and 28.3% accuracy respectively. The classifier identified 85% of MA panoramic images accurately.

**Figure 2B.**
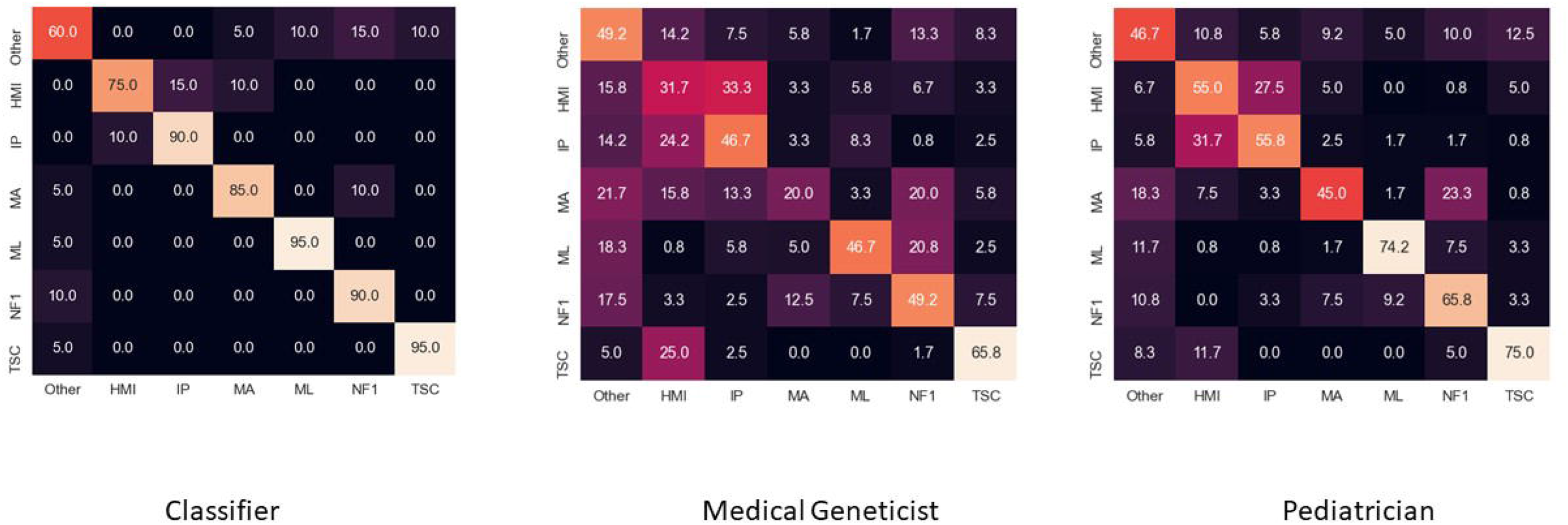
Confusion matrix comparing our classifier (right) versus the two different clinician types (middle and left) for classification of focused images. Rows represent the correct label, while columns represent the label chosen by the classifier or the clinicians. The diagonal numbers represent the percent accuracy for each category (the percentage of time the correct label was identified), while the off-diagonal numbers represent misclassifications, with the number corresponding to the percentage of time the label for a given image type was ascribed to another, incorrect category.

**Figure 2C.**
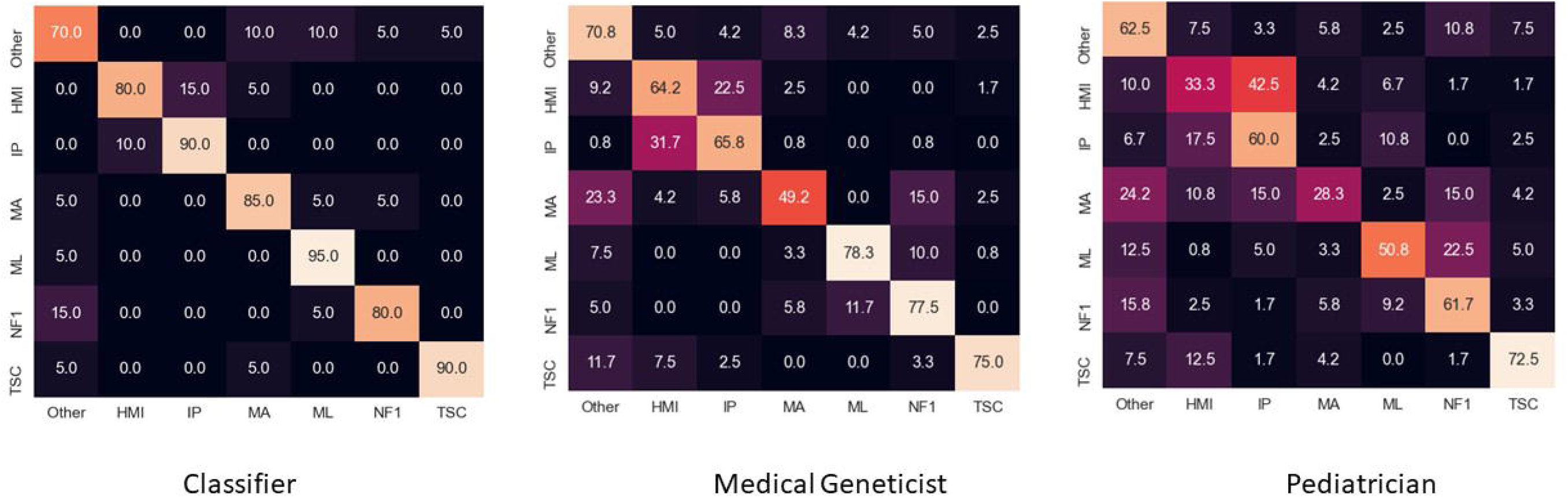
Confusion matrix comparing output of classifier versus the two different types of clinicians for classification of panoramic images. See Figure 2B legend for an explanation of the classifier.

We validated that our classifier obtains adequate accuracy because it “sees” important parts of an image. That is, the attribution method (Figure 3) helped us determine which pixels most affect the classifier’s decision making. This helped ensure that intuitively important pixels were being weighted more frequently than potential, common artifacts, such as recurrent background types or articles of clothing in the panoramic images.^22^

**Figure 3.**
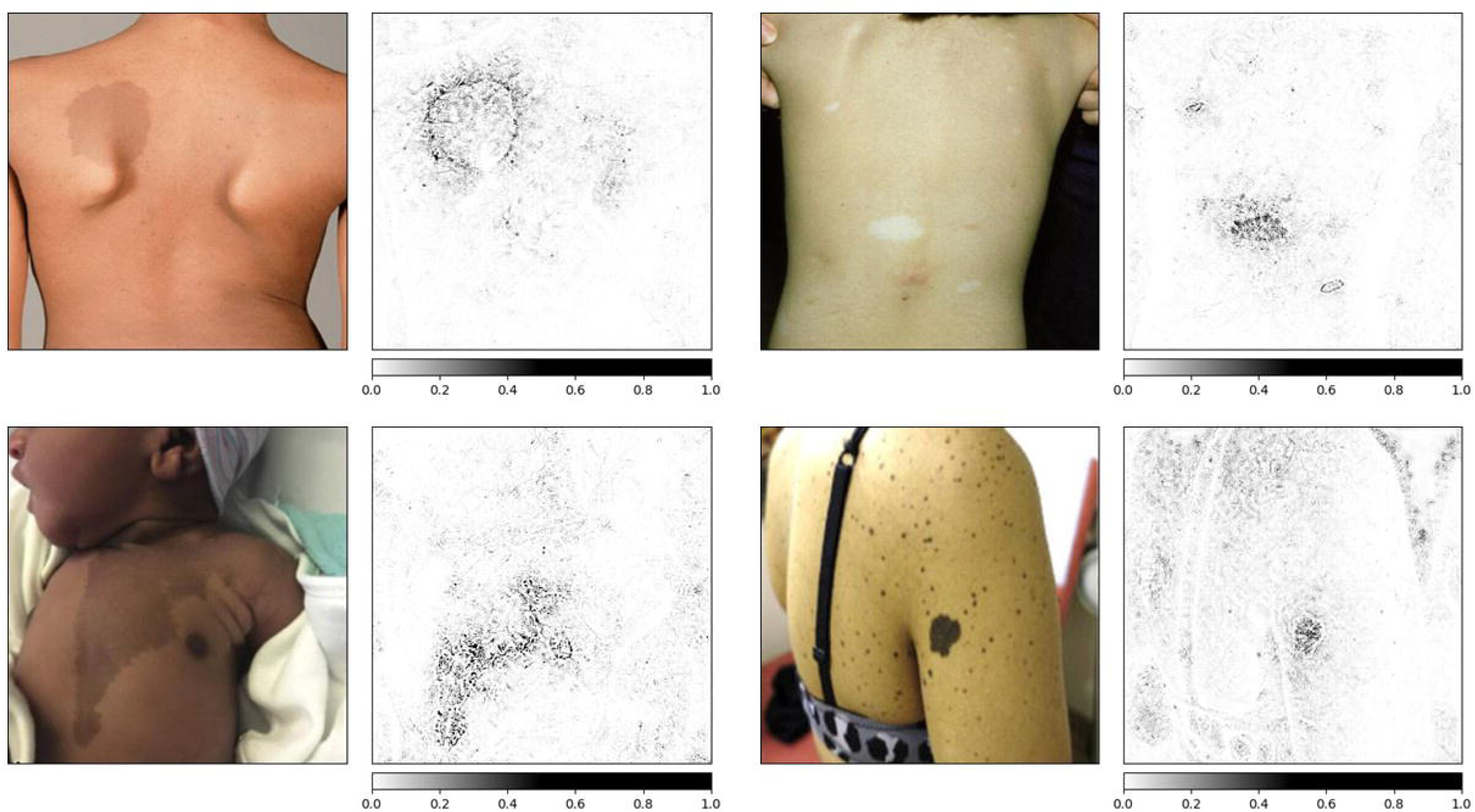
Attribution images, showing which pixels the classifier weights when “deciding” how to categorize. As shown, the classifier uses pixels involved in the skin finding, but may also use other pixels as well. Our research team examined these attribution methods during stages of classifier training and testing in order to determine how to improve performance, such as by incorporating other datasets for training, or when adjusting the neural network hyperparameters. Clockwise, from top left: Neurofibromatosis type 1 (NF1); Tuberous Sclerosis Complex (TSC); Noonan syndrome with Multiple Lentigines (ML); McCune-Albright syndrome (MA). Image sources (all used with appropriate permissions; see Supplemental data for permissions): NF1: https://pubmed.ncbi.nlm.nih.gov/24432075/#&gid=article-figures&pid=fig-2-uid-1 TSC: https://pubmed.ncbi.nlm.nih.gov/24143074/#&gid=article-figures&pid=figure-3-uid-2 ML: https://ars.els-cdn.com/content/image/1-s2.0-S000293431930347X-gr1_lrg.jpg MA: https://www.sciencedirect.com/science/article/pii/S235251261930044X?via%3Dihub#fig1

### Generative adversarial network, morphing, and style-mixing

Genetic conditions can have multiple manifestations, and these can change over time. In NF1, the earliest observable features are typically café-au-lait macules (as shown in Figure 1). Later, other features, such as cutaneous neurofibromas emerge.^15^ Our classifier focused on the former finding, since we were interested in identifying the condition in an early stage. However, we wanted to observe how these features could emerge and change over time. To do this, we trained a GAN to generate new images based on the collected dataset. Examples of images generated by our GAN are shown in Figure 4 and Supplemental Figure 1. Through the GAN, we also generated images to depict disease progression, as shown in Figure 5. New images generated through style-mixing are shown in Supplemental Figure 2. All newly generated images, and images obtained by morphing and style-mixing were shown to practicing genetics clinicians, who subjectively endorsed realistic output.

**Figure 4.**
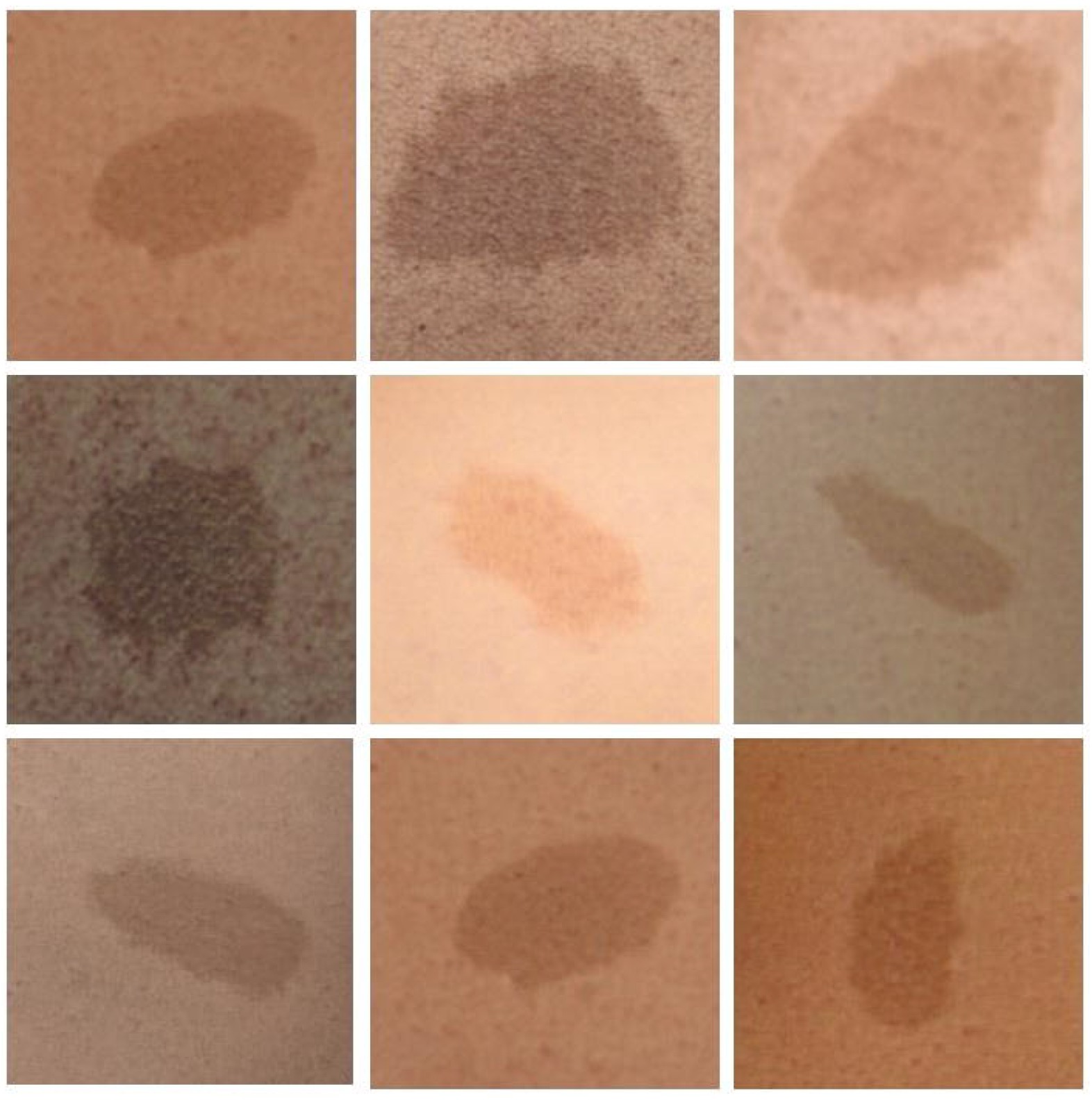
Examples of generated images of café-au-lait macules (CALMs) in Neurofibromatosis type 1 (NF1). Images, which were generated via a generative adversarial network (GAN),^13^ and which were felt to appear subjectively (by trained genetics clinicians) accurate, can be used for additional purposes, such as style-mixing or morphing.

**Figure 5.**
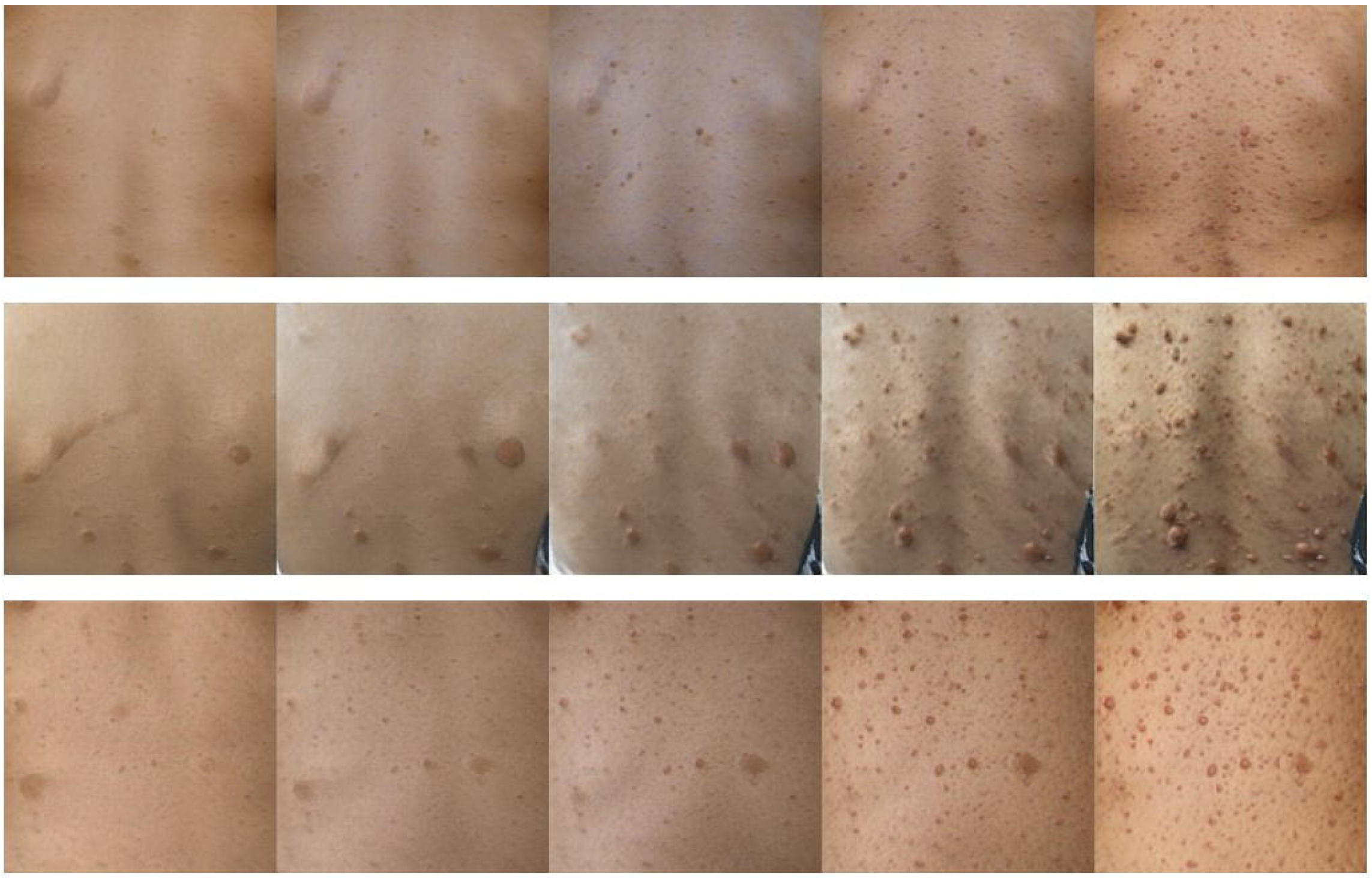
Examples of disease progression in Neurofibromatosis type 1 (NF1) generated via a generative adversarial network (GAN).^13^ These images show the accumulation of other features beyond café-au-lait macules (CALMs) such as cutaneous neurofibromas. Other features, such as the scapulae in some images, are also generated, since training images included features such as these.

## Discussion

Our overarching goal was to use neural networks to demonstrate how neural networks and related methods can be leveraged in potentially useful and interesting ways when applied to datasets involving genetic conditions. Our aim was not to build the most accurate classifier possible. We could achieve better accuracy with additional (computationally expensive) modifications, such as by further modifying the model’s hyperparameters or by incorporating larger or different datasets for training the model. We note that the availability of larger, centralized datasets relevant to genetic diseases such as those we analyzed is currently lacking compared to more common conditions.

Our classifier outperformed both pediatricians and geneticists for both focused and panoramic images. This is not meant to imply that the classifier can or should replace human experts. We examined a specific task (classifying images). This was done without incorporating other information that is often important in clinical practice, such as knowledge about family history and other clinical manifestations, like presence or absence of developmental delay or a history of certain types of cancer. These types of data could certainly be incorporated into a computer-based classifier, though such a model was not part of our objectives (including because we did not have uniform access to these data). Despite this, our experimental set-up did allow us to estimate how well the physicians perform when provided with the more holistic panoramic images as opposed to the focused lesion images. This estimation was not done in some other large-scale studies involving skin images.^19^

The fact that our classifier worked relatively well may demonstrate possible use cases. For example, this type of approach could help primary care doctors determine which patients should be prioritized for evaluation by subspecialists like geneticists. In settings – both in the United States and in other countries – with less access to specialists, these tools could also help identify the most efficient genetic testing strategy.

We observed a range of accuracy for the human experts. Several geneticists did extremely well, even though the average geneticist accuracy was lower than that of the classifier. This is logical: some geneticists may be more experienced with these specific conditions or may simply be more gifted at this type of task. The fact that the classifier does relatively well across different conditions – in addition to the overall accuracy, is notable. As an example, the confusion matrices (Figures 2B and 2C) show that the clinicians tend to do better with certain conditions than others. This may relate to the rarity of certain conditions that may be clinically important but may not be frequently encountered in clinical practice or medical training. This is one advantage of computerized methods, where a very rare condition can be included if adequate overall data can be gathered.

We built a potentially useful method using relatively small datasets – we were able to build the classifying algorithm using a minimum of about 100 images per condition. This approach is important when considering methods for conditions that are relatively rare. We also endeavored to include individuals of diverse ancestral backgrounds when collecting our training datasets. As the racial and ethnic background of most individuals whose images we used was not described in the primary literature, this was difficult to quantify, and requires further testing and attention in this type of work.^23,24^

The methods we built can also be readily modified. For example, other conditions of interest could be incorporated by collecting additional images and retraining our classifier, which can be done quickly using the code we provide.

Finally in relation to the classifier, one concern about neural network and related methods is that they are a “black box” that is opaque to human intuition or explanation. Our attribution methods show that one can – to an extent – correlate which features are important for the computer classifier to make decisions. This was useful during the classifier building process to ensure that pixels were weighted in what would be considered a logical fashion. This is not dissimilar to how a human might identify which condition a person has. That is, the human may pay more attention to certain, informative features, such as the shape of a skin lesion, or the angle of a bone on an x-ray, or the existence of an affected family member, to decide which condition might be most likely to affect a person. One of the skills physicians pick up during their career is knowing which features deserve attention, and which are less important. Using computer-based attribution methods can similarly help understand which features a model uses.

We also aimed to show how assembling and analyzing datasets via neural networks could be used beyond classification. For example, as mentioned above, creating new images could be useful for educational purposes, including to ensure that more images would be available regarding how conditions can appear in individuals of diverse ancestries. Our images, on expert assessment, appear to be realistic representations. However, this again demands further study, as approaches like style-mixing could inadvertently introduce or ignore differences due to factors that impact how a condition may manifest in a certain population. Separately, the morphing techniques may help depict the prognosis of a condition.

This could be also useful for the education of patients and clinicians or could be a valuable adjunct (when coupled with highly accurate computer-based measurements) for clinical trials.

While our work provides novel insights into the use of advanced computational approaches in the study of rare diseases, our study does have limitations. In collecting a large enough dataset, we were reliant on publicly available images and information. While our clinical team vetted each image, it is possible that some of our data was inaccurate. For example, it is possible that some depicted images were from a patients with more than one genetic condition,^25^ which could complicate the phenotype. The genetic conditions analyzed can have genetic heterogeneity (can occur due to different genetic causes) or can involve distinct genotype-phenotype correlations. As we treated each condition collectively (as a single entity), we were not able to parse out unique attributes to a given genetic variant or subset of a given condition. Additionally, our approach did not account for possible overlaps between the conditions, such as might occur in the two RASopathies: NF1 and ML. As our major focus of our approach is to build methods that can be useful with smaller datasets, our accuracy was not as high as it would be with a larger dataset. We anticipate that as publicly available data sets are established for rare diseases, coupled with our methods, work in this area will approach the accuracy of that described for more common conditions. Our work with morphing and style-mixing is highly exploratory. We plan to study how these and other techniques can aid healthcare practitioners in their practices. Lastly, we compared our classifier to two types of physicians, clinicians who most frequently encounter these types of conditions collectively. However, other clinicians may have different (better or worse) abilities to classify some conditions. For example, dermatologists, family practitioners, neonatologists, neurologists, and other specialists may yield different results. The point of our classification comparison was not to devise a head-to-head competition, but rather to use our approaches to illustrate and explore the utility of advanced analytics in rare genetic diseases.

## Supporting information

Supplemental material (survey)

NIH cover sheet

COI disclosure form

Image permission

Image permission

Image permission

Image permission

Image permission

Image permission

Image permission

Image permission

## Data Availability

All data, including code, have been made available, either through posting at GitHub, or via the links (URLs) available in the Supplementary materials.

https://github.com/datduong/ClassifyNF1

https://github.com/datduong/stylegan2-ada-MorphNF1

## Acknowledgments

This research was supported by the Intramural Research Program of the National Human Genome Research Institute, National Institutes of Health. The authors thank Vence L. Bonham, Jr, JD, Senior Advisor to the NHGRI Director on Genomics and Health Disparities and Head of the NHGRI Health Disparities Unit for his comments regarding considerations related to ancestral diversity and Daniel L. Kastner, MD, PhD, for his mentorship and guidance regarding research plans.

## Figure Legends

**Supplemental Figure 1**. Top row: examples of generated images of café-au-lait macules (CALMs) in Neurofibromatosis type 1 (NF1). Bottom row: examples of generated images of CALMs in McCune-Albright syndrome (MA).

**Supplemental Figure 2**. Generation of new images to illustrate a potential method to address issues related to diversity, especially in smaller datasets. Rows indicate source images (the styles to be transferred over), and columns indicate the target images. Skin and lesion colors are transferred to the source, but the shape of the lesions in the source remained unchanged. If both sources and targets are realistic, we can create larger sets of diverse images, which can be useful, but output must be carefully checked to ensure that the results are realistic and do not introduce new sources of bias, such as by ignoring features that may be more or less frequent in certain populations.

