## Supplementary figures and images for "Proof-of-principle neural network models for classification, attribution, creation, style-mixing, and morphing of image data for genetic conditions"

### Figure S1

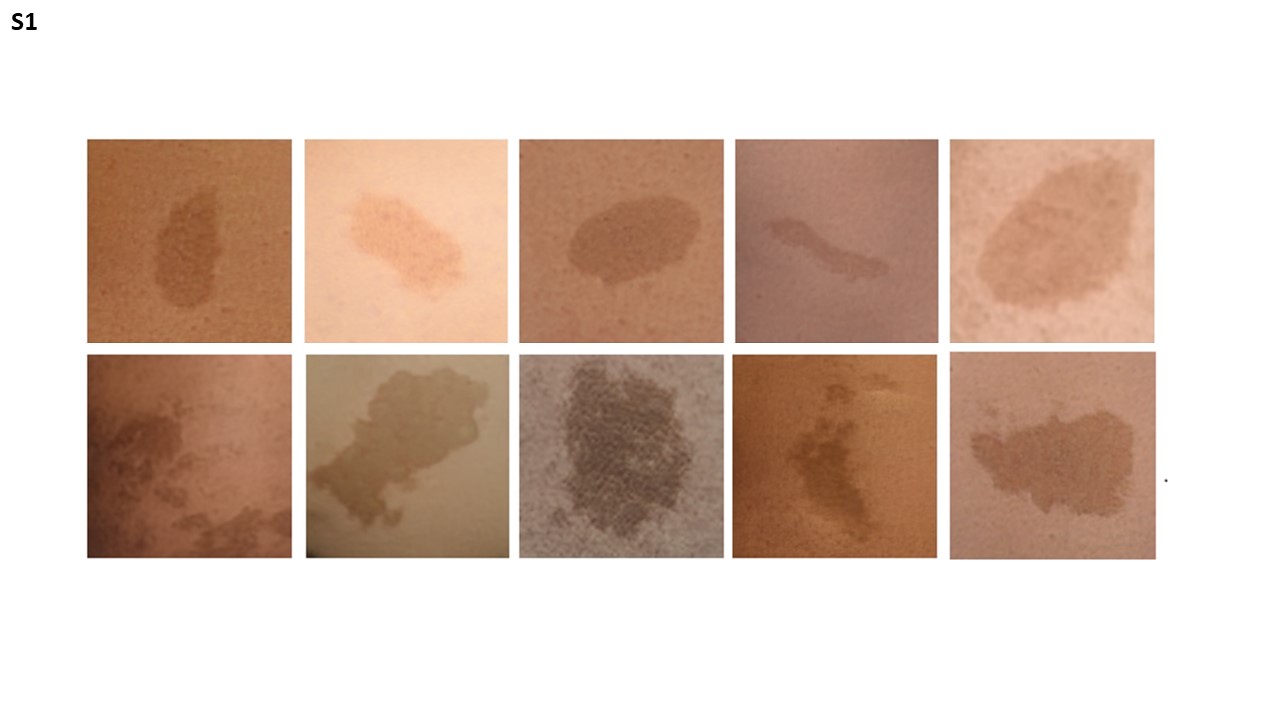

### Figure S2

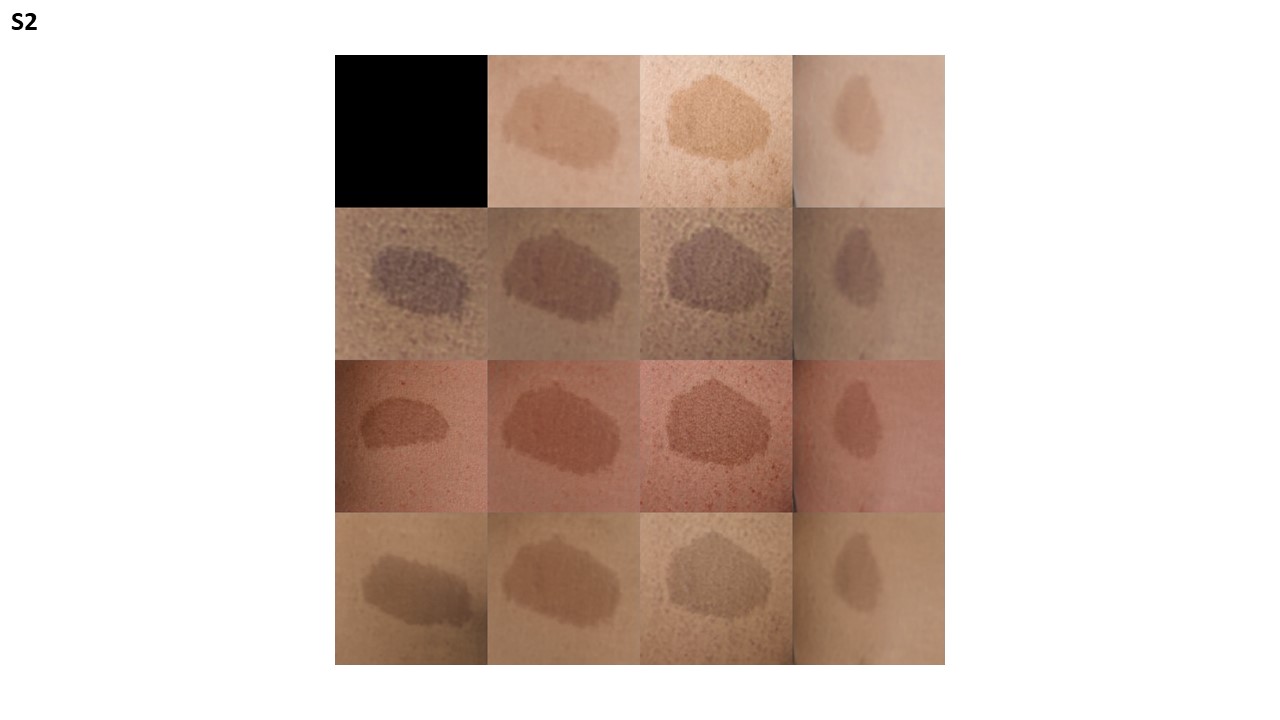
