## Supplemental Table 1 for "Proof-of-principle neural network models for classification, attribution, creation, style-mixing, and morphing of image data for genetic conditions"

**Supplemental Table 1.** Description of genetic conditions studied.

| **Syndrome/condition** | **Inheritance pattern** | **De novo rates** | **Gene** | **Prevalence per 100,000 (prevalence in population unless otherwise stated)^b^** | **Skin manifestations** | **Common extracutaneous syndrome/condition features (features can be highly variable, including because some conditions may have different molecular/genomic etiologies)** |
| --- | --- | --- | --- | --- | --- | --- |
| Hypomelanosis of Ito ^1,2^ | Likely Mosaic | Unknown | Multiple | 1.2 | Hypopigmentation following the lines of Blaschko*; alopecia; patchy depigmentation | Variable depending on underlying cytogenomic cause and affected tissures, but can include: seizures; scoliosis; chest wall deformity; digit abnormalities; hemihypertrophy; retinal hypopigmentation |
| Incontinentia Pigmenti ^3^ | X-linked Dominant, Mosaic | 65% | IKBKG | 1.2 (birth prevalence) | Blisters (Stage I), Wart-like rash (Stage 2) Swirling macular hyperpigmentation (Stage 3)^a^; Linear hypopigmentation (Stage 4) | Seizures; intellectual disability; male embryonic lethality; leukocytosis |
| McCune-Albright Syndrome ^4^ | Mosaic | 100% | GNAS | 0.55 | Café au lait pigmentation^a^ associated with midline of body, classically described as having jagged and irregular boarders | Fibrous dysplasia; scoliosis; hemihypertrophy; precocious puberty; excessive growth hormone; hyperthyroidism |
| Neurofibromatosis Type 1 ^5-7^ | Autosomal Dominant | 42%^8^ | NF1 | 33.3 (birth prevalence) | Café au lait macules (CALMs)^a^, classically described as having smooth borders; dermal neurofibromas; axillary and inguinal freckling; xanthogranuloma; nevus anemicus | Plexiform neurofibromas; Lisch nodules; choroidal freckling; optic gliomas |
| Noonan with Multiple Lentigines ^8^ | Autosomal Dominant | Unknown | PTPN11, RAF1, BRAF, MAP2K1 | Unknown | Multiple lentigine^a^; CALMs^a^ | Hypertrophic cardiomyopathy; pectus deformity; short stature; hypertelorism; dysmorphic facial features; intellectual disability |
| Tuberous Sclerosis Complex ^9,10^ | Autosomal Dominant | 80%^11^ | TSC1, TSC2 | 10.0 | Hypomelanotic macules (white ash leaf spots)^a^; confetti skin lesions; angiofibromas; shagreen patches, fibrous cephalic plaques; ungual fibromas | Seizures; intellectual disability; behavioral disturbances; subependymal nodules, cortical dysplasia, subependymal giant cell astrocytomas; renal disease; cardiac rhbdomyomas; lymphangioleimyomatosis |

^a^ Skin manifestation used in study.

^b^ Orphanet. Prevalence and incidence of rare diseases: Bibliographic data, January 2019, Number 01. 2019. https://www.orpha.net/orphacom/cahiers/docs/GB/Prevalence_of_rare_diseases_by_diseases.pdf. Accessed 16 March 2021.
