## Supplemental Table 2 for "Proof-of-principle neural network models for classification, attribution, creation, style-mixing, and morphing of image data for genetic conditions"

**Supplemental Table 2.** URLs for images used for classifier (links are subject to change). Conditions studied: Hypomelanosis of Ito (HMI), Incontinentia Pigmenti (IP), McCune-Albright Syndrome (MA), Neurofibromatosis Type 1 (NF1), Noonan Syndrome with Multiple Lentigines (ML; formally known as LEOPARD syndrome), Tuberous sclerosis Complex (TSC), and other (other skin conditions). For each row, an asterisk (*) indicates that the corresponding image was used in the testing set. URLs without an asterisk are to images that were used in the training set. Results of the comparison of human and ML classification of test set images are shown in Figure 2.

| URL | Condition |
| --- | --- |
| <https://casereports.bmj.com/content/12/4/e227693> | HMI |
| https://casereports.bmj.com/content/12/4/e227693 | HMI |
| <https://www.researchgate.net/figure/Hypomelanosis-of-Ito-Hypopigmentation-on-the-back-of-trunk-on-the-right-side-V-shaped_fig1_305991168> | HMI |
| <https://www.dermatologyadvisor.com/home/decision-support-in-medicine/dermatology/hypomelanosis-of-ito-incontinentia-pigmenti-achromians/> | HMI |
| <https://twitter.com/HITSWorldwide1/photo> | HMI |
| <https://ajns.paans.org/hypomelanosis-of-ito-a-case-report-from-east-africa/> | HMI* |
| <https://www.webmedcentral.com/article_view/3292> | HMI* |
| https://www.webmedcentral.com/article_view/3292 | HMI* |
| <https://cdn.ymaws.com/www.aocd.org/resource/resmgr/jaocd/contents/volume32/32-14.pdf> | HMI |
| https://cdn.ymaws.com/www.aocd.org/resource/resmgr/jaocd/contents/volume32/32-14.pdf | HMI |
| https://cdn.ymaws.com/www.aocd.org/resource/resmgr/jaocd/contents/volume32/32-14.pdf | HMI |
| <https://indianpediatrics.net/dec2000/dec-1386.htm> | HMI |
| <https://www.kinderaerztliche-praxis.de/a/hypomelanosis-ito-incontinentia-pigmenti-achromians-1693377> | HMI |
| <https://www.childrensmercy.org/health-care-providers/providers/connect-with-childrens-mercy/newsletter-the-link/whats-the-diagnosis-Dec-2018/> | HMI |
| <https://www.sciencedirect.com/science/article/pii/S0738081X04001804#fig2> | HMI* |
| <https://obgynkey.com/patterned-pigmentation-in-children/> | HMI* |
| <https://obgynkey.com/hypopigmentation-disorders/> | HMI |
| <https://obgynkey.com/hypopigmentation-disorders/> | HMI |
| <https://escholarship.org/uc/item/6324c2ww> | HMI |
| <https://plasticsurgerykey.com/hypomelanosis-of-ito/> | HMI |
| <https://plasticsurgerykey.com/hypomelanosis-of-ito/> | HMI |
| <https://www.facebook.com/231316036636/posts/-hypomelanosis-of-ito-is-a-sporadic-neurocutaneous-disorder-characterized-cutane/10152719440756637/> | HMI |
| <https://www.facebook.com/231316036636/posts/-hypomelanosis-of-ito-is-a-sporadic-neurocutaneous-disorder-characterized-cutane/10152719440756637/> | HMI |
| <https://www.semanticscholar.org/paper/Hypomelanosis-of-Ito-Sugathan-Grabowski/446431f694e2a5869845c2086819475fad58cd73> | HMI |
| <https://www.semanticscholar.org/paper/Hypomelanosis-of-Ito-Sugathan-Grabowski/446431f694e2a5869845c2086819475fad58cd73/figure/0> | HMI* |
| <https://www.sciencedirect.com/science/article/pii/B9780444627025000214> | HMI |
| https://www.sciencedirect.com/science/article/pii/B9780444627025000214 | HMI |
| <https://casereports.bmj.com/content/casereports/2018/bcr-2018-225055/F2.large.jpg> | HMI* |
| <https://www.semanticscholar.org/paper/Hypomelanosis-of-Ito-with-multiple-congenital-Yu-Kwon/a61fe2a7c0cac70acb3392384e110a0434074003> | HMI |
| https://www.semanticscholar.org/paper/Hypomelanosis-of-Ito-with-multiple-congenital-Yu-Kwon/a61fe2a7c0cac70acb3392384e110a0434074003 | HMI |
| https://www.semanticscholar.org/paper/Hypomelanosis-of-Ito-with-multiple-congenital-Yu-Kwon/a61fe2a7c0cac70acb3392384e110a0434074003 | HMI |
| <https://www.researchgate.net/publication/297756796_Total_Hemi-overgrowth_in_Pigmentary_Mosaicism_of_the_Hypomelanosis_of_Ito_Type_Eight_Case_Reports> | HMI |
| https://www.researchgate.net/publication/297756796_Total_Hemi-overgrowth_in_Pigmentary_Mosaicism_of_the_Hypomelanosis_of_Ito_Type_Eight_Case_Reports | HMI |
| <http://biblioteca.asmn.re.it/pubblicazioni/Pediatric%20Dermatol%202007%2024%203.pdf> | HMI |
| <https://onlinelibrary.wiley.com/doi/abs/10.1111/j.1525-1470.2007.00387.x> | HMI |
| <http://www.pigmentinternational.com/article.asp?issn=2349-5847;year=2015;volume=2;issue=1;spage=64;epage=66;aulast=Das> | HMI |
| http://www.pigmentinternational.com/article.asp?issn=2349-5847;year=2015;volume=2;issue=1;spage=64;epage=66;aulast=Das | HMI |
| <https://obgynkey.com/pigmentary-development-of-east-asian-skin/> | HMI |
| <https://link.springer.com/referenceworkentry/10.1007%2F978-3-642-02202-9_148> | HMI |
| https://link.springer.com/referenceworkentry/10.1007%2F978-3-642-02202-9_148 | HMI |
| <https://www.wikidoc.org/index.php/File:Incontinentia_pigmenti_achromians_of_Ito07.jpg> | HMI* |
| <https://www.wikidoc.org/index.php/File:Incontinentia_pigmenti_achromians_of_Ito14.jpg> | HMI* |
| https://www.wikidoc.org/index.php/File:Incontinentia_pigmenti_achromians_of_Ito03.jpg | HMI |
| <https://www.wikidoc.org/index.php/File:Incontinentia_pigmenti_achromians_of_Ito02.jpg> | HMI |
| <https://n.neurology.org/content/80/12/e130> | HMI |
| <https://jmedicalcasereports.biomedcentral.com/articles/10.1186/1752-1947-8-156/figures/3> | HMI* |
| <https://emedicine.medscape.com/article/909996-overview> | HMI* |
| <https://onlinelibrary.wiley.com/doi/full/10.1111/ajd.12032> | HMI |
| <https://www.researchgate.net/figure/Whorly-pattern-of-hypo-pigmentation-in-a-child-with-hypomelanosis-of-ito_fig3_335187206> | HMI |
| <https://casereports.bmj.com/content/casereports/2018/bcr-2018-225055/F1.large.jpg> | HMI* |
| <https://twitter.com/globaldermie/status/666223665038663680/photo/2> | HMI* |
| <https://twitter.com/globaldermie/status/666223665038663680/photo/4> | HMI |
| https://twitter.com/globaldermie/status/666223665038663680/photo/4 | HMI |
| <https://link.springer.com/content/pdf/10.1007/s004390050848.pdf> | HMI |
| https://link.springer.com/content/pdf/10.1007/s004390050848.pdf | HMI |
| <https://1library.net/document/lq5ogprz-study-on-linear-dermatoses.html> | HMI* |
| <https://file.scirp.org/Html/8-1460135_22990.htm> | HMI |
| https://file.scirp.org/Html/8-1460135_22990.htm | HMI |
| <https://www.wikidoc.org/index.php/File:Incontinentia_pigmenti_achromians_of_Ito15.jpg> | HMI |
| https://www.wikidoc.org/index.php/File:Incontinentia_pigmenti_achromians_of_Ito01.jpg | HMI |
| https://www.wikidoc.org/index.php/File:Incontinentia_pigmenti_achromians_of_Ito09.jpg | HMI |
| https://www.wikidoc.org/index.php/File:Incontinentia_pigmenti_achromians_of_Ito09.jpg | HMI |
| https://www.wikidoc.org/index.php/File:Incontinentia_pigmenti_achromians_of_Ito14.jpg | HMI |
| https://www.wikidoc.org/index.php/File:Incontinentia_pigmenti_achromians_of_Ito02.jpg | HMI |
| <https://www.aerzteblatt.de/app/print.asp?id=42493> | HMI |
| <https://www.sciencedirect.com/topics/medicine-and-dentistry/hypopigmentation> | HMI* |
| <https://radiologykey.com/70-17/> | HMI |
| <https://radiologykey.com/70-17/> | HMI |
| <https://onlinelibrary.wiley.com/doi/full/10.1002/mgg3.546> | HMI |
| <https://media.springernature.com/original/springer-static/image/chp%3A10.1007%2F978-3-319-18389-3_8/MediaObjects/318522_1_En_8_Fig13_HTML.jpg> | HMI* |
| <https://www.chegg.com/flashcards/genodermatoses-13a740ca-86ee-4397-9f94-527884bd2984/deck> | HMI |
| <https://www.thelancet.com/journals/lancet/article/PIIS0140-6736(03)14076-7/fulltext> | HMI |
| <https://www.thelancet.com/journals/lancet/article/PIIS0140-6736(03)14076-7/fulltext> | HMI |
| <http://medpics011.blogspot.com/2013/12/hypomelanosis-of-ito.html> | HMI |
| <http://medpics011.blogspot.com/2013/12/hypomelanosis-of-ito.html> | HMI |
| <http://medpics011.blogspot.com/2013/12/hypomelanosis-of-ito.html> | HMI |
| <https://www.scielo.br/scielo.php?pid=S1679-45082014000400529&script=sci_arttext&tlng=en> | HMI |
| <https://www.instagram.com/explore/tags/blaschko/> | HMI* |
| <https://www.instagram.com/explore/tags/blaschko/> | HMI* |
| <https://www.instagram.com/explore/tags/blaschko/> | HMI |
| <https://www.sciencedirect.com/science/article/pii/S0022347689803324?via%3Dihub> | HMI |
| <https://www.sciencedirect.com/science/article/pii/S0022347689803324?via%3Dihub> | HMI |
| <https://link.springer.com/article/10.1007/s00381-020-04758-5#Fig9> | HMI |
| <https://plasticsurgerykey.com/genodermatoses-and-congenital-anomalies/> | HMI* |
| <https://www.sciencedirect.com/science/article/pii/S0022347619314933?via%3Dihub> | HMI |
| <https://www.sciencedirect.com/science/article/pii/S0022347619314933?via%3Dihub> | HMI |
| <https://onlinelibrary.wiley.com/doi/full/10.1111/pde.13913> | HMI |
| <https://onlinelibrary.wiley.com/doi/full/10.1111/pde.13913> | HMI |
| <https://onlinelibrary.wiley.com/doi/full/10.1111/pde.13913> | HMI |
| <https://onlinelibrary.wiley.com/doi/full/10.1111/pde.13913> | HMI |
| <https://onlinelibrary.wiley.com/doi/full/10.1111/pde.13913> | HMI |
| <https://onlinelibrary.wiley.com/doi/full/10.1111/ijd.14393> | HMI |
| <https://onlinelibrary.wiley.com/doi/full/10.1002/ajmg.a.31940> | HMI |
| <https://onlinelibrary.wiley.com/doi/full/10.1002/ajmg.a.31940> | HMI |
| <https://onlinelibrary.wiley.com/doi/full/10.1002/ajmg.a.31940> | HMI |
| <https://onlinelibrary.wiley.com/doi/full/10.1111/jdv.13737> | HMI |
| <https://www.sciencedirect.com/science/article/pii/S0022347615006071?via%3Dihub#dfig1> | HMI |
| <https://www.scielo.br/scielo.php?script=sci_arttext&pid=S0004-282X2015000400366&lng=en&nrm=iso&tlng=en> | HMI |
| <https://www.scielo.br/scielo.php?script=sci_arttext&pid=S0004-282X2015000400366&lng=en&nrm=iso&tlng=en> | HMI |
| <https://link.springer.com/article/10.1007/s10072-014-2049-1/figures/1> | HMI |
| <https://www.ncbi.nlm.nih.gov/pmc/articles/PMC4030355/> | HMI |
| <https://thejns.org/pediatrics/view/journals/j-neurosurg-pediatr/10/3/article-p182.xml> | HMI |
| <https://www.sciencedirect.com/science/article/pii/S0887899411003572?via%3Dihub#fig1> | HMI |
| <https://onlinelibrary.wiley.com/doi/full/10.1111/j.1365-4632.2011.04974.x> | HMI |
| <https://onlinelibrary.wiley.com/doi/full/10.1111/j.1365-4632.2010.04639.x> | HMI |
| <https://onlinelibrary.wiley.com/doi/full/10.1111/j.1365-4632.2010.04639.x> | HMI |
| <https://onlinelibrary.wiley.com/doi/full/10.1111/j.1365-2230.2009.03208.x> | HMI* |
| <https://www.sciencedirect.com/science/article/pii/S0887899408005900?via%3Dihub#fig1> | HMI |
| <https://www.sciencedirect.com/science/article/pii/S107190910800082X?via%3Dihub#fig1> | HMI |
| <https://www.sciencedirect.com/science/article/pii/S107190910800082X?via%3Dihub#fig1> | HMI |
| <https://www.sciencedirect.com/science/article/pii/S107190910800082X?via%3Dihub#fig1> | HMI |
| <https://www.sciencedirect.com/science/article/pii/S107190910800082X?via%3Dihub#fig1> | HMI |
| <https://academic.oup.com/ndt/article/22/6/1796/1926821> | HMI |
| <https://www.huidziekten.nl/quiz/dermatologiequiz45q.htm> | HMI |
| <https://pedsinreview.aappublications.org/content/28/5/193> | HMI |
| <https://pedsinreview.aappublications.org/content/28/5/193> | HMI |
| <https://www.dermaamin.com/site/atlas-of-dermatology/7-h/688-hypomelanosis-of-ito-.html> | HMI |
| <https://aemende.wordpress.com/2015/10/27/the-disorder-incontinentia-pigmenti-or-bloch-sulzberger-syndrome/> | IP |
| <https://dermnetnz.org/topics/incontinentia-pigmenti/> | IP |
| <https://dermnetnz.org/topics/incontinentia-pigmenti/> | IP |
| <https://edsociety.co.uk/what-is-ed/types-of-ed/welcome-to-the-incontinentia-pigmenti-section/> | IP |
| <https://edsociety.co.uk/what-is-ed/types-of-ed/welcome-to-the-incontinentia-pigmenti-section/> | IP |
| <https://pedsinreview.aappublications.org/content/28/11/429> | IP* |
| <https://pedsinreview.aappublications.org/content/28/11/429> | IP |
| <https://pedsinreview.aappublications.org/content/28/11/429> | IP |
| <https://pedsinreview.aappublications.org/content/28/11/429> | IP |
| <https://pedsinreview.aappublications.org/content/28/11/429> | IP |
| <https://2017.prepsa.courses.aap.org/PrepSA/Modal/Media.aspx?key=a25275d4-daa7-4162-8e52-0afc5d8c4145> | IP* |
| <https://plasticsurgerykey.com/130-incontinentia-pigmenti/> | IP* |
| <https://www.researchgate.net/figure/X-linked-Incontinentia-pigmenti-Pattern-type-1a-Blaschko-lines-narrow-bands_fig3_257074473> | IP |
| <https://www.nfed.org/learn/types/incontinentia-pigmenti/> | IP |
| <https://www.elrincondelamedicinainterna.com/2017/03/incontinencia-pigmenti.html> | IP |
| <https://www.elrincondelamedicinainterna.com/2017/03/incontinencia-pigmenti.html> | IP |
| <https://www.elrincondelamedicinainterna.com/2017/03/incontinencia-pigmenti.html> | IP |
| <https://www.elrincondelamedicinainterna.com/2017/03/incontinencia-pigmenti.html> | IP |
| <https://pediatrics.aappublications.org/content/100/4/e6/tab-article-info> | IP |
| <https://www.elrincondelamedicinainterna.com/2019/12/incontinencia-pigmentariaa-proposito-de.html> | IP* |
| <https://www.elrincondelamedicinainterna.com/2019/12/incontinencia-pigmentariaa-proposito-de.html> | IP |
| <https://www.elrincondelamedicinainterna.com/2019/12/incontinencia-pigmentariaa-proposito-de.html> | IP |
| <https://www.elrincondelamedicinainterna.com/2019/12/incontinencia-pigmentariaa-proposito-de.html> | IP |
| <https://www.elrincondelamedicinainterna.com/2019/12/incontinencia-pigmentariaa-proposito-de.html> | IP |
| <https://www.sciencedirect.com/topics/pharmacology-toxicology-and-pharmaceutical-science/incontinentia-pigmenti> | IP |
| <https://www.sciencedirect.com/topics/pharmacology-toxicology-and-pharmaceutical-science/incontinentia-pigmenti> | IP |
| <http://juberos6.blogspot.com/2016/07/incontinencia-pigmenti.html> | IP* |
| <https://www.sciencedirect.com/science/article/abs/pii/S0891524517302663> | IP |
| <https://www.sciencedirect.com/science/article/abs/pii/S0891524517302663> | IP |
| <https://www.jpedhc.org/article/S0891-5245(17)30266-3/pdf> | IP |
| <https://healthjade.net/incontinentia-pigmenti/> | IP |
| <http://dxline.info/diseases/incontinentia-pigmenti/#prettyPhoto[pp_gal]/1/> | IP |
| <http://dxline.info/diseases/incontinentia-pigmenti/#prettyPhoto[pp_gal]/1/> | IP |
| <http://www.ijmr.org.in/article.asp?issn=0971-5916%3Byear=2011%3Bvolume=133%3Bissue=4%3Bspage=442%3Bepage=445%3Baulast=Thakur> | IP |
| <http://www.ijmr.org.in/article.asp?issn=0971-5916%3Byear=2011%3Bvolume=133%3Bissue=4%3Bspage=442%3Bepage=445%3Baulast=Thakur> | IP |
| <https://escholarship.org/uc/item/9dz2p5bk> | IP* |
| <https://www.sciencedirect.com/science/article/pii/S0929693X11002727> | IP |
| <https://www.google.com/search?q=incontinentia%20pigmenti&tbm=isch&hl=en&hl=en&tbs=rimg%3ACRaUPpRRsTFCYYaiVr-Ejwv8&rlz=1C1GCEA_enUS888US888&sa=X&ved=0CBsQuIIBahcKEwiwzo_95NzrAhUAAAAAHQAAAAAQBw&biw=1560&bih=1323#imgrc=vqQX513pCiL_iM> | IP |
| <https://www.semanticscholar.org/paper/Incontinentia-pigmenti%3A-a-newborn-with-skin-lesions-Azizzadeh-Rezaei/68544928f3fe3ac7f7f969398f8475a9e5da0b72/figure/0> | IP |
| <https://www.huidziekten.nl/zakboek/dermatosen/itxt/IncontinentiaPigmenti.htm> | IP |
| <https://www.huidziekten.nl/zakboek/dermatosen/itxt/IncontinentiaPigmenti.htm> | IP |
| <https://www.huidziekten.nl/zakboek/dermatosen/itxt/IncontinentiaPigmenti.htm> | IP* |
| <https://www.jaypeedigital.com/book/9788184480146/chapter/ch29> | IP |
| <https://www.medscape.com/viewarticle/776025_1> | IP |
| <http://pennstatehershey.adam.com/content.aspx?productId=112&pid=2&gid=2071> | IP |
| <http://pennstatehershey.adam.com/content.aspx?productId=112&pid=2&gid=2071> | IP |
| <https://www.wikidoc.org/index.php/File:Incontinentia_pigmenti05.jpg> | IP |
| <https://www.wikidoc.org/index.php/File:Incontinentia_pigmenti06.jpg> | IP |
| <https://emedicine.medscape.com/article/1176285-clinical#b4> | IP |
| <https://emedicine.medscape.com/article/1176285-clinical#b4> | IP |
| <http://www.ijo.in/viewimage.asp?img=IndianJOphthalmol_2011_59_3_255_81022_u1.jpg> | IP |
| <https://ghr.nlm.nih.gov/condition/incontinentia-pigmenti> | IP |
| <https://ghr.nlm.nih.gov/condition/incontinentia-pigmenti> | IP |
| <https://ghr.nlm.nih.gov/condition/incontinentia-pigmenti> | IP |
| <https://ghr.nlm.nih.gov/condition/incontinentia-pigmenti> | IP |
| <https://www.ncbi.nlm.nih.gov/pmc/articles/PMC3726911/> | IP* |
| <https://www.sochiderm.org/web/revista/28_2/6.pdf> | IP |
| <https://www.sochiderm.org/web/revista/28_2/6.pdf> | IP |
| <https://www.analesdepediatria.org/en-incontinentia-pigmenti-manifestaciones-iniciales-largo-articulo-resumen-S1695403308700353> | IP |
| <https://www.analesdepediatria.org/en-incontinentia-pigmenti-manifestaciones-iniciales-largo-articulo-resumen-S1695403308700353> | IP |
| <https://link.springer.com/referenceworkentry/10.1007%2F978-1-4939-2401-1_133> | IP |
| <https://link.springer.com/referenceworkentry/10.1007%2F978-1-4939-2401-1_133> | IP |
| <http://www.ijpd.in/viewimage.asp?img=IndianJPaediatrDermatol_2016_17_1_24_173154_f2.jpg> | IP |
| <http://www.ijpd.in/viewimage.asp?img=IndianJPaediatrDermatol_2016_17_1_24_173154_f2.jpg> | IP |
| <https://jkms.org/ViewImage.php?Type=F&aid=503520&id=F1&afn=63_JKMS_21_3_474&fn=jkms-21-474-g001_0063JKMS> | IP |
| <https://www.consultant360.com/exclusives/incontinentia-pigmenti-neonate> | IP |
| <https://www.flickr.com/photos/46651330@N00/234913378/> | IP |
| <https://www.clinicaladvisor.com/slideshow/clinical-quiz/derm-dx-linear-streaky-vesicles-at-birth-2/> | IP |
| <https://www.clinicaladvisor.com/slideshow/clinical-quiz/derm-dx-linear-streaky-vesicles-at-birth-2/> | IP |
| <https://www.clinicaladvisor.com/slideshow/clinical-quiz/derm-dx-linear-streaky-vesicles-at-birth-2/> | IP |
| <https://www.clinicaladvisor.com/slideshow/clinical-quiz/derm-dx-linear-streaky-vesicles-at-birth-2/> | IP |
| <https://www.clinicaladvisor.com/slideshow/clinical-quiz/derm-dx-linear-streaky-vesicles-at-birth-2/> | IP |
| <https://m2.healio.com/~/media/journals/osli/2013/1_january/10_3928_23258160_20121221_20/fig1.jpg> | IP |
| <https://m2.healio.com/~/media/journals/osli/2013/1_january/10_3928_23258160_20121221_20/fig1.jpg> | IP |
| <https://www.pedneur.com/article/S0887-8994(18)31051-8/abstract> | IP |
| <https://www.pedneur.com/article/S0887-8994(18)31051-8/abstract> | IP |
| <https://www.peertechz.com/articles/IJDCR-4-128.php> | IP |
| <https://escholarship.org/uc/item/6vq7m9zr> | IP* |
| <http://www.bmhim.com/files/bmhim_20_77_3_112-118.pdf> | IP |
| <http://www.bmhim.com/files/bmhim_20_77_3_112-118.pdf> | IP |
| <https://www.ijdvl.com/article.asp?issn=0378-6323;year=2020;volume=86;issue=4;spage=422;epage=424;aulast=Bishnoi> | IP |
| <https://www.ijdvl.com/article.asp?issn=0378-6323;year=2020;volume=86;issue=4;spage=422;epage=424;aulast=Bishnoi> | IP |
| <https://www.ijdvl.com/article.asp?issn=0378-6323;year=2020;volume=86;issue=4;spage=422;epage=424;aulast=Bishnoi> | IP |
| <https://www.ijdvl.com/article.asp?issn=0378-6323;year=2020;volume=86;issue=4;spage=422;epage=424;aulast=Bishnoi> | IP |
| <https://www.ijdvl.com/article.asp?issn=0378-6323;year=2020;volume=86;issue=4;spage=422;epage=424;aulast=Bishnoi> | IP |
| <https://www.ncbi.nlm.nih.gov/pmc/articles/PMC7195961/> | IP |
| <https://www.ncbi.nlm.nih.gov/pmc/articles/PMC7195961/> | IP |
| <https://www.ncbi.nlm.nih.gov/pmc/articles/PMC7195961/> | IP |
| <https://www.sciencedirect.com/science/article/pii/S015196381931083X?via%3Dihub#fig0010> | IP |
| <https://www.sciencedirect.com/science/article/pii/S015196381931083X?via%3Dihub#fig0010> | IP |
| <https://onlinelibrary.wiley.com/doi/full/10.1111/ped.13964> | IP |
| <https://onlinelibrary.wiley.com/doi/full/10.1111/ped.13964> | IP |
| <https://onlinelibrary.wiley.com/doi/full/10.1111/ped.13964> | IP |
| <https://onlinelibrary.wiley.com/doi/full/10.1111/bjd.17319> | IP |
| <https://journals.lww.com/amjdermatopathology/Fulltext/2019/09000/A_Female_Infant_With_Linear_Erythema_and.22.aspx> | IP* |
| <https://journals.lww.com/amjdermatopathology/Fulltext/2019/09000/A_Female_Infant_With_Linear_Erythema_and.22.aspx> | IP |
| <https://www.sciencedirect.com/science/article/pii/S0738081X19301221?via%3Dihub#f0015> | IP |
| <https://www.sciencedirect.com/science/article/pii/S0738081X19301221?via%3Dihub#f0015> | IP |
| <https://www.sciencedirect.com/science/article/pii/S0738081X19301221?via%3Dihub#f0015> | IP |
| <https://www.sciencedirect.com/science/article/pii/S0738081X19301221?via%3Dihub#f0015> | IP |
| <https://www.sciencedirect.com/science/article/pii/S0738081X19301221?via%3Dihub#f0015> | IP* |
| <https://journals.lww.com/continuum/Fulltext/2018/02000/Neurocutaneous_Disorders.8.aspx> | IP* |
| <https://www.jle.com/en/revues/ejd/e-docs/detection_of_hpv_15_in_painful_subungual_tumors_of_incontinentia_pigmenti_successful_topical_therapy_with_retinoic_acid_280922/article.phtml> | IP |
| <https://www.ijo.in/article.asp?issn=0301-4738;year=2019;volume=67;issue=6;spage=940;epage=942;aulast=Rishi;type=3> | IP |
| <https://link.springer.com/article/10.1007/s00431-012-1855-9> | IP |
| <https://link.springer.com/article/10.1007/s00431-012-1855-9> | IP |
| <https://www.sciencedirect.com/science/article/pii/S1695403319301705?via%3Dihub#fig0005> | IP* |
| <https://www.sciencedirect.com/science/article/pii/S0001731018304903?via%3Dihub#fig0010> | IP |
| <https://www.sciencedirect.com/science/article/pii/S0001731018304903?via%3Dihub#fig0010> | IP |
| <https://www.sciencedirect.com/science/article/pii/S0001731018304903?via%3Dihub#fig0010> | IP |
| <https://onlinelibrary.wiley.com/doi/full/10.1111/1346-8138.14002> | IP |
| <https://onlinelibrary.wiley.com/doi/full/10.1111/1346-8138.14002> | IP |
| <https://onlinelibrary.wiley.com/doi/full/10.1111/j.1365-4632.2007.03365.x> | IP |
| <https://onlinelibrary.wiley.com/cms/asset/b5f926bd-1fbd-49a8-8a9a-9fc629882872/ijd_2156_f3.gif> | IP* |
| <https://link.springer.com/article/10.1007/s13555-019-00336-z> | IP |
| <https://link.springer.com/article/10.1007/s13555-019-00336-z> | IP |
| <https://www.dermaamin.com/site/atlas-of-dermatology/8-i/709-incontinentia-pigmenti-.html> | IP |
| <https://www.dermaamin.com/site/atlas-of-dermatology/8-i/709-incontinentia-pigmenti-.html> | IP |
| <https://www.dermaamin.com/site/atlas-of-dermatology/8-i/709-incontinentia-pigmenti-.html> | IP |
| <https://www.dermaamin.com/site/atlas-of-dermatology/8-i/709-incontinentia-pigmenti-.html> | IP* |
| <https://www.dermaamin.com/site/atlas-of-dermatology/8-i/709-incontinentia-pigmenti-.html> | IP |
| <https://www.dermaamin.com/site/atlas-of-dermatology/8-i/709-incontinentia-pigmenti-.html> | IP |
| <https://www.dermaamin.com/site/atlas-of-dermatology/8-i/709-incontinentia-pigmenti-.html> | IP* |
| <https://www.dermaamin.com/site/atlas-of-dermatology/8-i/709-incontinentia-pigmenti-.html> | IP* |
| <https://www.dermaamin.com/site/atlas-of-dermatology/8-i/709-incontinentia-pigmenti-.html> | IP* |
| <https://www.dermaamin.com/site/atlas-of-dermatology/8-i/709-incontinentia-pigmenti-.html> | IP |
| <https://www.dermaamin.com/site/atlas-of-dermatology/8-i/709-incontinentia-pigmenti-.html> | IP |
| <https://www.dermaamin.com/site/atlas-of-dermatology/8-i/709-incontinentia-pigmenti-.html> | IP* |
| <https://www.dermaamin.com/site/atlas-of-dermatology/8-i/709-incontinentia-pigmenti-.html> | IP |
| <https://www.dermaamin.com/site/atlas-of-dermatology/8-i/709-incontinentia-pigmenti-.html> | IP |
| <https://www.dermaamin.com/site/atlas-of-dermatology/8-i/709-incontinentia-pigmenti-.html> | IP |
| <https://www.dermaamin.com/site/atlas-of-dermatology/8-i/709-incontinentia-pigmenti-.html> | IP |
| <https://www.dermaamin.com/site/atlas-of-dermatology/8-i/709-incontinentia-pigmenti-.html> | IP* |
| <https://www.dermaamin.com/site/atlas-of-dermatology/8-i/709-incontinentia-pigmenti-.html> | IP |
| <https://www.dermaamin.com/site/atlas-of-dermatology/8-i/709-incontinentia-pigmenti-.html> | IP |
| <https://www.medscape.com/answers/127233-155901/which-physical-findings-are-characteristic-of-caf-au-lait-pigmentation-in-mccune-albright-syndrome-mas> | MA |
| <https://www.medscape.com/answers/127233-155901/which-physical-findings-are-characteristic-of-caf-au-lait-pigmentation-in-mccune-albright-syndrome-mas> | MA |
| <https://www.medscape.com/answers/127233-155901/which-physical-findings-are-characteristic-of-caf-au-lait-pigmentation-in-mccune-albright-syndrome-mas> | MA |
| <https://www.medscape.com/answers/127233-155901/which-physical-findings-are-characteristic-of-caf-au-lait-pigmentation-in-mccune-albright-syndrome-mas> | MA |
| <https://www.consultant360.com/articles/caf-au-lait-spots> | MA |
| <https://www.wikidoc.org/index.php/File:Cafe_au_lait_(with_MR,_it%27s_McCune_Albright).jpg> | MA |
| <https://www.consultant360.com/articles/photo-essay-hyperpigmented-macules> | MA |
| <https://www.consultant360.com/articles/photo-essay-hyperpigmented-macules> | MA |
| <https://www.sciencedirect.com/topics/nursing-and-health-professions/cafe-au-lait-spot> | MA |
| <https://www.jcdr.net/ReadXMLFile.aspx?id=11564> | MA |
| <https://clinicalgate.com/disorders-of-hyperpigmentation-and-melanocytes/> | MA |
| <https://accesspediatrics.mhmedical.com/content.aspx?bookid=2674&sectionid=220537705> | MA |
| <https://www.researchgate.net/figure/Representative-Cafe-au-lait-Spots-Seen-in-McCune-Albright-Syndrome-A-spectrum-of-spots_fig4_225077911> | MA |
| <https://www.researchgate.net/figure/Representative-Cafe-au-lait-Spots-Seen-in-McCune-Albright-Syndrome-A-spectrum-of-spots_fig4_225077911> | MA |
| <https://www.researchgate.net/figure/Representative-Cafe-au-lait-Spots-Seen-in-McCune-Albright-Syndrome-A-spectrum-of-spots_fig4_225077911> | MA |
| <https://www.researchgate.net/figure/Representative-Cafe-au-lait-Spots-Seen-in-McCune-Albright-Syndrome-A-spectrum-of-spots_fig4_225077911> | MA |
| <https://www.researchgate.net/figure/Representative-Cafe-au-lait-Spots-Seen-in-McCune-Albright-Syndrome-A-spectrum-of-spots_fig4_225077911> | MA |
| <https://www.researchgate.net/figure/Representative-Cafe-au-lait-Spots-Seen-in-McCune-Albright-Syndrome-A-spectrum-of-spots_fig4_225077911> | MA |
| <https://www.semanticscholar.org/paper/%5BMcCune-Albright-syndrome-revealed-by-caf%C3%A9-au-lait-Jung-Soskin/18526db965064109d9fbf909517af04853098b28> | MA* |
| [https://www.semanticscholar.org/paper/[McCune-Albright-syndrome-revealed-by-caf%C3%A9-au-lait-Jung-Soskin/18526db965064109d9fbf909517af04853098b28/figure/1](https://www.semanticscholar.org/paper/%5bMcCune-Albright-syndrome-revealed-by-caf%C3%A9-au-lait-Jung-Soskin/18526db965064109d9fbf909517af04853098b28/figure/1) | MA |
| <https://www.slideserve.com/catori/cutaneous-mosaicism-a-molecular-and-clinical-point-of-view> | MA |
| <https://www.slideserve.com/catori/cutaneous-mosaicism-a-molecular-and-clinical-point-of-view> | MA |
| <https://www.mja.com.au/journal/2006/185/11/mccune-albright-syndrome> | MA |
| <http://img.medcubic.com/index/category/38> | MA |
| <https://twitter.com/VisualDx/status/1191729131681042432> | MA |
| <https://ojrd.biomedcentral.com/articles/10.1186/s13023-019-1102-9/figures/1> | MA |
| <https://ojrd.biomedcentral.com/articles/10.1186/s13023-019-1102-9/figures/1> | MA |
| <https://ojrd.biomedcentral.com/articles/10.1186/s13023-019-1102-9/figures/1> | MA |
| <https://ojrd.biomedcentral.com/articles/10.1186/s13023-019-1102-9/figures/1> | MA |
| <http://archivos.pap.es/Empty/PAP/front/Articulos/Imprimir/_OrCjUxDG4croFblaIuWJHzVPjyNsJ3us94oWRCvxdfM> | MA |
| <http://archivos.pap.es/Empty/PAP/front/Articulos/Imprimir/_OrCjUxDG4croFblaIuWJHzVPjyNsJ3us94oWRCvxdfM> | MA |
| <https://www.huidziekten.nl/zakboek/dermatosen/atxt/Albright.htm> | MA* |
| https://www.huidziekten.nl/zakboek/dermatosen/atxt/Albright.htm | MA |
| https://www.huidziekten.nl/zakboek/dermatosen/atxt/Albright.htm | MA |
| <https://onlinelibrary.wiley.com/doi/pdf/10.1111/odi.12563> | MA |
| <https://www.eurorad.org/case/14477> | MA* |
| <https://link.springer.com/article/10.1186/1757-1626-2-9376> | MA |
| <https://neoreviews.aappublications.org/content/suppl/2003/10/01/4.10.e263.DC1> | MA |
| <https://www.magicfoundation.org/Growth-Disorders/McCune-Albright-Syndrome-or-Fibrous-Dysplasia/> | MA |
| <https://link.springer.com/article/10.1007/s00223-019-00550-z/figures/8> | MA |
| <http://www.jgid.org/viewimage.asp?img=JGlobalInfectDis_2012_4_4_215_103901_u3.jpg> | MA |
| <http://www.jgid.org/viewimage.asp?img=JGlobalInfectDis_2012_4_4_215_103901_u3.jpg> | MA |
| <https://www.semanticscholar.org/paper/Mazabraud-syndrome-associated-with-McCune-Albright-Biazzo-Bernardo/ede60247c79f21a495224a1fb5702acd943eca33/figure/0> | MA |
| <https://www.researchgate.net/figure/Cafe-au-lait-macules-of-an-18-month-old-girl-with-McCune-Albright-syndrome-MAS_fig1_326131321> | MA |
| <http://medpics011.blogspot.com/2014/02/mccune-albright-syndrome.html> | MA |
| <https://www.semanticscholar.org/paper/McCune-Albright-syndrome-without-endocrine-Case-in-Sami-Pankaj/dd9a27ed7a174798c44e51c84796d6d0bab1d31c> | MA |
| <https://www.semanticscholar.org/paper/McCune-Albright-syndrome-without-endocrine-Case-in-Sami-Pankaj/dd9a27ed7a174798c44e51c84796d6d0bab1d31c> | MA |
| <https://www.sciencedirect.com/science/article/pii/S0003426616000032> | MA |
| <https://ghr.nlm.nih.gov/condition/mccune-albright-syndrome> | MA |
| <https://ghr.nlm.nih.gov/condition/mccune-albright-syndrome> | MA |
| <https://www.researchgate.net/publication/327958494_Fibrous_Dysplasia/figures> | MA* |
| <https://www.researchgate.net/publication/327958494_Fibrous_Dysplasia/figures> | MA |
| <https://www.researchgate.net/publication/327958494_Fibrous_Dysplasia/figures> | MA |
| <https://www.researchgate.net/publication/327958494_Fibrous_Dysplasia/figures> | MA |
| <http://www.repdevmed.org/article.asp?issn=2096-2924;year=2018;volume=2;issue=4;spage=252;epage=255;aulast=Guan> | MA |
| <http://www.ijpd.in/viewimage.asp?img=IndianJPaediatrDermatol_2015_16_4_272_165674_f1.jpg> | MA |
| <http://www.ijpd.in/viewimage.asp?img=IndianJPaediatrDermatol_2015_16_4_272_165674_f1.jpg> | MA |
| <http://www.hormones.gr/156/article/article.html> | MA |
| <http://www.hormones.gr/156/article/article.html> | MA |
| <https://www.orthobullets.com/pathology/8038/fibrous-dysplasia> | MA |
| <https://link.springer.com/chapter/10.1007/978-3-319-99817-6_11> | MA |
| <https://link.springer.com/chapter/10.1007/978-3-319-99817-6_11> | MA |
| <https://en.wikipedia.org/wiki/McCune%E2%80%93Albright_syndrome> | MA |
| <https://www.researchgate.net/figure/Image-of-a-child-with-McCune-Albright-syndrome-with-light-brown-spots-on-the-skin-and-a_fig4_339831172> | MA |
| <https://www.semanticscholar.org/paper/Neonatal-McCune%E2%80%93Albright-syndrome-with-systemic-a-Louren%C3%A7o-Dias/9e817ba5522d06a2acd94afd213b69231b40f66f> | MA |
| <https://twitter.com/JuliaDemanett/status/1192859229994049536/photo/1> | MA* |
| <https://pubmed.ncbi.nlm.nih.gov/20157193/> | MA |
| <https://pubmed.ncbi.nlm.nih.gov/20157193/> | MA |
| <https://www.medscape.com/answers/925446-182295/which-conditions-are-associated-with-gigantism> | MA |
| <https://online.boneandjoint.org.uk/doi/full/10.1007/s11832-007-0006-8> | MA |
| <https://www.facebook.com/endopractice/photos/a.1732984706972205/2503956279875040/?type=3&theater> | MA* |
| <https://www.cancertherapyadvisor.com/home/decision-support-in-medicine/obstetrics-and-gynecology/precocious-puberty/> | MA |
| <https://www.cancertherapyadvisor.com/home/decision-support-in-medicine/obstetrics-and-gynecology/precocious-puberty/> | MA |
| <https://casereports.bmj.com/content/2018/bcr-2018-225709> | MA |
| <https://musculoskeletalkey.com/oncology-and-pathology/> | MA |
| <http://www.medibone.cn/Wenxianlist.aspx?key=&Type_ID=47&page=13> | MA* |
| <https://link.springer.com/article/10.1186/1750-1172-7-S1-S4> | MA |
| <https://surgicalneurologyint.com/surgicalint-articles/mccune-albright-syndrome-with-craniofacial-dysplasia-clinical-review-and-surgical-management/> | MA |
| <http://saspjournals.com/wp-content/uploads/2015/06/SJMCR-35402-406.pdf> | MA |
| <https://eje.bioscientifica.com/view/journals/eje/182/5/EJE-19-0969.xml> | MA |
| <https://eje.bioscientifica.com/view/journals/eje/182/5/EJE-19-0969.xml> | MA |
| <https://online.boneandjoint.org.uk/doi/full/10.1007/s11832-007-0006-8> | MA |
| <https://online.boneandjoint.org.uk/doi/full/10.1007/s11832-007-0006-8> | MA |
| <https://online.boneandjoint.org.uk/doi/full/10.1007/s11832-007-0006-8> | MA |
| <https://www.google.com/url?sa=i&url=https%3A%2F%2Fwww.ijrcog.org%2Findex.php%2Fijrcog%2Farticle%2Fdownload%2F6352%2F4510&psig=AOvVaw1G-ajGwKLDBD12EF2sIPrW&ust=1601750093895000&source=images&cd=vfe&ved=0CAMQjB1qFwoTCODqnJ_GluwCFQAAAAAdAAAAABAn> | MA |
| <https://oncohemakey.com/skeletal-dysplasias/> | MA |
| <https://oncohemakey.com/skeletal-dysplasias/> | MA |
| <https://pubmed.ncbi.nlm.nih.gov/32849305/#&gid=article-figures&pid=figure-1-uid-0> | MA |
| <https://pubmed.ncbi.nlm.nih.gov/32849305/#&gid=article-figures&pid=figure-1-uid-0> | MA |
| <https://pubmed.ncbi.nlm.nih.gov/32190188/> | MA* |
| <https://casereports.bmj.com/content/12/7/e229141.long> | MA* |
| <https://www.jaadcasereports.org/article/S2352-5126(19)30044-X/pdf> | MA |
| <https://pubmed.ncbi.nlm.nih.gov/29991465/> | MA |
| <https://www.karger.com/Article/FullText/473878> | MA |
| <https://www.ncbi.nlm.nih.gov/pmc/articles/PMC4933654/> | MA |
| <https://www.sciencedirect.com/science/article/pii/S0929693X1100580X?via%3Dihub#fig0005> | MA |
| <https://www.sciencedirect.com/science/article/pii/S0929693X1100580X?via%3Dihub#fig0005> | MA |
| <https://www.ncbi.nlm.nih.gov/pmc/articles/PMC4140081/> | MA |
| <https://www.ncbi.nlm.nih.gov/pmc/articles/PMC4140081/> | MA |
| <https://www.ncbi.nlm.nih.gov/pmc/articles/PMC3121281/> | MA |
| <https://www.sciencedirect.com/science/article/pii/S0151963810004862?via%3Dihub#fig0005> | MA |
| <https://insights.ovid.com/pubmed?pmid=21272121> | MA |
| <https://www.karger.com/Article/FullText/284365> | MA |
| <https://link.springer.com/article/10.1007%2Fs12020-007-0015-x> | MA |
| <https://www.sciencedirect.com/science/article/pii/S1530891X20414491?via%3Dihub> | MA |
| <https://www.ncbi.nlm.nih.gov/pmc/articles/PMC2687599/> | MA |
| <https://link.springer.com/article/10.1385%2FENDO%3A18%3A2%3A121> | MA |
| <https://onlinelibrary.wiley.com/doi/full/10.1046/j.1525-1470.2001.1862003.x?sid=nlm%3Apubmed> | MA |
| <https://onlinelibrary.wiley.com/doi/full/10.1046/j.1525-1470.2001.1862003.x?sid=nlm%3Apubmed> | MA |
| <https://www.scielo.br/scielo.php?script=sci_arttext&pid=S0103-64402008000200014&lng=en&nrm=iso&tlng=en> | MA |
| <https://www.scielo.br/scielo.php?script=sci_arttext&pid=S0103-64402008000200014&lng=en&nrm=iso&tlng=en> | MA |
| <https://media.springernature.com/original/springer-static/image/chp%3A10.1007%2F978-3-319-99817-6_11/MediaObjects/340511_1_En_11_Fig4_HTML.jpg> | MA |
| <https://ourorthopaedics.blogspot.com/2008/01/51-mccune-albright-syndrome-mas.html> | MA* |
| <https://www.sciencedirect.com/science/article/pii/S2212440313002782#fig1> | MA |
| <https://oncohemakey.com/gonadotropin-independent-precocious-puberty/> | MA |
| <https://academic.oup.com/edrv/article/41/2/345/5610851> | MA |
| <https://academic.oup.com/edrv/article/41/2/345/5610851> | MA |
| <https://academic.oup.com/edrv/article/41/2/345/5610851> | MA* |
| <https://academic.oup.com/edrv/article/41/2/345/5610851> | MA |
| <https://www.ijdvl.com/article.asp?issn=0378-6323;year=2010;volume=76;issue=6;spage=723;epage=724;aulast=Patel> | MA |
| <https://www.ijdvl.com/article.asp?issn=0378-6323;year=2010;volume=76;issue=6;spage=723;epage=724;aulast=Patel> | MA |
| <https://www.ijdvl.com/article.asp?issn=0378-6323;year=2010;volume=76;issue=6;spage=723;epage=724;aulast=Patel> | MA* |
| <https://austinpublishinggroup.com/gynecology-case-reports/fulltext/agcr-v1-id1007.php> | MA |
| <https://austinpublishinggroup.com/gynecology-case-reports/fulltext/agcr-v1-id1007.php> | MA |
| <https://jebmh.com/latest_articles/19> | MA* |
| <https://www.semanticscholar.org/paper/McCune-Albright-syndrome-presenting-with-unilateral-Khanna-Kantawala/1abd69eb7161d248e8964f1bdfd050288bcbb4aa> | MA* |
| <https://www.liebertpub.com/doi/pdf/10.1089/thy.1997.7.433> | MA |
| <https://www.liebertpub.com/doi/pdf/10.1089/thy.1997.7.433> | MA |
| <https://www.consultant360.com/articles/hyperpigmented-macules> | MA |
| <https://www.consultant360.com/articles/hyperpigmented-macules> | MA |
| <https://www.ncbi.nlm.nih.gov/books/NBK279155/figure/gigantism_f_gigantism_figure2/> | MA* |
| <https://www.ncbi.nlm.nih.gov/pmc/articles/PMC5035212/pdf/nihms-808453.pdf> | MA |
| <https://www.karger.com/Article/PDF/504802> | MA* |
| <https://www.karger.com/Article/PDF/504802> | MA* |
| <https://casereports.bmj.com/content/12/7/e229141.long> | MA |
| <https://www.sciencedirect.com/science/article/pii/S0151963810004862?via%3Dihub> | MA* |
| <https://www.sciencedirect.com/science/article/pii/S0151963810004862?via%3Dihub> | MA* |
| <https://www.ncbi.nlm.nih.gov/pmc/articles/PMC4560899/pdf/13256_2015_Article_689.pdf> | MA |
| <https://www.ncbi.nlm.nih.gov/pmc/articles/PMC5329869/pdf/med-2016-0082.pdf> | MA |
| <https://www.ncbi.nlm.nih.gov/pmc/articles/PMC4523984/pdf/tpa-50-2-114.pdf> | MA |
| <https://www.ncbi.nlm.nih.gov/pmc/articles/PMC5111499/pdf/cm-89-559.pdf> | MA |
| <https://www.ncbi.nlm.nih.gov/pmc/articles/PMC5111499/pdf/cm-89-559.pdf> | MA* |
| <https://link.springer.com/chapter/10.1007/978-3-319-44824-4_20> | MA |
| <https://bmcmedgenet.biomedcentral.com/articles/10.1186/1471-2350-15-44> | ML |
| <https://dermnetnz.org/topics/noonan-syndrome-with-multiple-lentigines/> | ML |
| <https://dermnetnz.org/topics/noonan-syndrome-with-multiple-lentigines/> | ML |
| <https://www.consultant360.com/articles/leopard-syndrome-progressively-increasing-pigmented-macules-8-year-old-boy> | ML |
| <https://www.dermatologyadvisor.com/home/decision-support-in-medicine/dermatology/leopard-syndrome-lentigines-ekg-abnormalities-ocular-hypertelorism-pulmonary-stenosis-abnormal-genitalia-retarded-growth-deafness-also-known-as-multiple-lentigines-syndrome-and-cardiocutaneous/> | ML |
| <https://pmj.bmj.com/content/94/1116/605> | ML |
| <https://pmj.bmj.com/content/94/1116/605> | ML |
| <https://pmj.bmj.com/content/94/1116/605> | ML |
| <https://onlinelibrary.wiley.com/doi/abs/10.1111/j.1525-1470.2008.00734.x> | ML |
| <https://onlinelibrary.wiley.com/doi/abs/10.1111/j.1525-1470.2008.00734.x> | ML |
| <https://onlinelibrary.wiley.com/doi/abs/10.1111/j.1525-1470.2008.00734.x> | ML |
| <https://onlinelibrary.wiley.com/doi/abs/10.1111/j.1525-1470.2008.00734.x> | ML |
| <https://www.semanticscholar.org/paper/LEOPARD-syndrome-(PTPN11%2C-T468M)-in-three-boys-type-Carcavilla-Pinto/bf4edc24a922b4ba349b4c847a258ab23cf78ee7> | ML |
| <https://medicalpicturesinfo.com/leopard-syndrome/> | ML* |
| <https://ojrd.biomedcentral.com/articles/10.1186/1750-1172-3-13/figures/2> | ML |
| <https://ojrd.biomedcentral.com/articles/10.1186/1750-1172-3-13/figures/2> | ML |
| <https://www.visualdx.com/visualdx/diagnosis/multiple+lentigines+syndrome?diagnosisId=51984&moduleId=101> | ML |
| <https://www.crutchfielddermatology.com/caseofthemonth/studies/2009/l_2009_010.asp> | ML* |
| <https://www.crutchfielddermatology.com/caseofthemonth/studies/2009/l_2009_010.asp> | ML |
| <https://www.crutchfielddermatology.com/caseofthemonth/studies/2009/l_2009_010.asp> | ML |
| <https://www.crutchfielddermatology.com/caseofthemonth/studies/2009/l_2009_010.asp> | ML |
| <https://www.crutchfielddermatology.com/caseofthemonth/studies/2009/l_2009_010.asp> | ML |
| <http://medical-dictionary.thefreedictionary.com/_/viewer.aspx?path=MosbyMD&name=leopard-syndrome.jpg&url=http%3A%2F%2Fmedical-dictionary.thefreedictionary.com%2FLEOPARD%2Bsyndrome> | ML |
| <http://medical-dictionary.thefreedictionary.com/_/viewer.aspx?path=MosbyMD&name=leopard-syndrome.jpg&url=http%3A%2F%2Fmedical-dictionary.thefreedictionary.com%2FLEOPARD%2Bsyndrome> | ML |
| <http://medical-dictionary.thefreedictionary.com/_/viewer.aspx?path=MosbyMD&name=leopard-syndrome.jpg&url=http%3A%2F%2Fmedical-dictionary.thefreedictionary.com%2FLEOPARD%2Bsyndrome> | ML |
| <https://syndromespedia.com/leopard-syndrome.html> | ML |
| <https://www.semanticscholar.org/paper/PTPN11-mutation-manifesting-as-LEOPARD-syndrome-and-Spatola-Wider/c9f79bcac8b8930a07d70fe74921ef1d1b208874> | ML |
| <https://www.semanticscholar.org/paper/PTPN11-mutation-manifesting-as-LEOPARD-syndrome-and-Spatola-Wider/c9f79bcac8b8930a07d70fe74921ef1d1b208874> | ML |
| <https://medlineplus.gov/genetics/condition/noonan-syndrome-with-multiple-lentigines/> | ML |
| <https://medlineplus.gov/genetics/condition/noonan-syndrome-with-multiple-lentigines/> | ML |
| <https://plasticsurgerykey.com/genodermatoses-and-congenital-anomalies/> | ML* |
| <https://www.ijdvl.com/article.asp?issn=0378-6323;year=2013;volume=79;issue=6;spage=821;epage=824;aulast=Ghosh> | ML |
| <https://www.ijdvl.com/article.asp?issn=0378-6323;year=2013;volume=79;issue=6;spage=821;epage=824;aulast=Ghosh> | ML |
| <https://www.ijdvl.com/article.asp?issn=0378-6323;year=2013;volume=79;issue=6;spage=821;epage=824;aulast=Ghosh> | ML |
| <https://www.ijdvl.com/article.asp?issn=0378-6323;year=2013;volume=79;issue=6;spage=821;epage=824;aulast=Ghosh> | ML |
| <https://dermaamin.com/site/images/clinical-pic/L/leopard-syndrome/leopard-syndrome24.jpg> | ML |
| <https://dermaamin.com/site/images/clinical-pic/L/leopard-syndrome/leopard-syndrome4.jpg> | ML |
| <https://dermaamin.com/site/images/clinical-pic/L/leopard-syndrome/leopard-syndrome4.jpg> | ML |
| <https://dermaamin.com/site/images/clinical-pic/L/leopard-syndrome/leopard-syndrome5.jpg> | ML* |
| <https://dermaamin.com/site/images/clinical-pic/L/leopard-syndrome/leopard-syndrome25.jpg> | ML |
| <https://dermaamin.com/site/images/clinical-pic/L/leopard-syndrome/leopard-syndrome10.jpg> | ML |
| <https://dermaamin.com/site/images/clinical-pic/L/leopard-syndrome/leopard-syndrome15.jpg> | ML* |
| <https://dermaamin.com/site/images/clinical-pic/L/leopard-syndrome/leopard-syndrome16.jpg> | ML* |
| <https://dermaamin.com/site/images/clinical-pic/L/leopard-syndrome/leopard-syndrome13.jpg> | ML |
| <https://dermaamin.com/site/images/clinical-pic/L/leopard-syndrome/leopard-syndrome13.jpg> | ML |
| <https://dermaamin.com/site/images/clinical-pic/L/leopard-syndrome/leopard-syndrome18.jpg> | ML |
| <https://onlinelibrary.wiley.com/doi/full/10.1111/bjd.17404> | ML |
| <https://onlinelibrary.wiley.com/doi/full/10.1111/bjd.17404> | ML |
| <https://www.huidziekten.nl/zakboek/dermatosen/ltxt/LeopardSyndroom.htm> | ML |
| <https://www.huidziekten.nl/zakboek/dermatosen/ltxt/LeopardSyndroom.htm> | ML |
| <https://reader.elsevier.com/reader/sd/pii/S0002914920308845?token=46566723012DF5556BC12DD4459D3D1BBB52F98C1F1BFD778EDC8456880BD57D839587B204B721332568BAF035FECE7B> | ML |
| <https://www.sciencedirect.com/science/article/pii/S000173102030168X?via%3Dihub#fig0005> | ML |
| <https://www.sciencedirect.com/science/article/pii/S000173102030168X?via%3Dihub#fig0005> | ML |
| <https://lib.ossn.ru/jour/article/view/944/641> | ML |
| <https://lib.ossn.ru/jour/article/view/944/641> | ML |
| <https://lib.ossn.ru/jour/article/view/944/641> | ML |
| <https://onlinelibrary.wiley.com/doi/epdf/10.1111/ddg.13880> | ML* |
| <https://www.sciencedirect.com/science/article/pii/S000293431930347X?via%3Dihub> | ML* |
| <https://onlinelibrary.wiley.com/doi/full/10.1002/ajmg.c.31692> | ML |
| <https://onlinelibrary.wiley.com/doi/full/10.1002/ajmg.c.31692> | ML* |
| <https://www.tandfonline.com/doi/full/10.1080/17843286.2018.1467531> | ML |
| <https://onlinelibrary.wiley.com/doi/full/10.1111/jdv.14573> | ML |
| <https://onlinelibrary.wiley.com/doi/full/10.1111/jdv.14573> | ML |
| <https://onlinelibrary.wiley.com/doi/full/10.1111/1346-8138.13960> | ML |
| <https://www.ncbi.nlm.nih.gov/pmc/articles/PMC5312195/> | ML |
| <https://www.medicaljournals.se/acta/content_files/files/pdf/97/4/4858.pdf> | ML |
| <https://onlinelibrary.wiley.com/doi/epdf/10.1111/cge.12728> | ML |
| <https://onlinelibrary.wiley.com/doi/epdf/10.1111/cge.12728> | ML |
| <https://onlinelibrary.wiley.com/doi/epdf/10.1111/cge.12728> | ML |
| <https://onlinelibrary.wiley.com/doi/epdf/10.1111/cge.12728> | ML |
| <https://onlinelibrary.wiley.com/doi/epdf/10.1111/cge.12728> | ML |
| <https://www.sciencedirect.com/science/article/pii/S176972121630088X?via%3Dihub#fig1> | ML* |
| <https://link.springer.com/article/10.1007/s00403-015-1597-4> | ML |
| <https://jmg.bmj.com/content/53/2/123.long> | ML |
| <https://jmg.bmj.com/content/53/2/123.long> | ML |
| <https://jmg.bmj.com/content/53/2/123.long> | ML |
| <https://jmg.bmj.com/content/53/2/123.long> | ML |
| <https://www.sciencedirect.com/science/article/pii/S0738081X14003137?via%3Dihub#f0010> | ML |
| <https://www.jisppd.com/viewimage.asp?img=JIndianSocPedodPrevDent_2015_33_1_57_149008_f2.jpg> | ML |
| <https://www.sciencedirect.com/science/article/pii/S0167527314008419?via%3Dihub#f0005> | ML |
| <https://www.ncbi.nlm.nih.gov/pmc/articles/PMC3778821/figure/F3/> | ML |
| <https://jpma.org.pk/article-details/4129?article_id=4129> | ML* |
| <https://www.sciencedirect.com/science/article/pii/S0190962212010237?via%3Dihub#fig1> | ML |
| <https://www.sciencedirect.com/science/article/pii/S0190962212010237?via%3Dihub#fig1> | ML |
| <https://www.ncbi.nlm.nih.gov/pmc/articles/PMC3507272/> | ML |
| <https://www.ncbi.nlm.nih.gov/pmc/articles/PMC3507272/> | ML* |
| <https://www.karger.com/Article/FullText/335995> | ML |
| <https://www.karger.com/Article/FullText/335995> | ML |
| <https://www.sciencedirect.com/science/article/pii/S0887899409000897?via%3Dihub#fig1> | ML |
| <https://www.jstage.jst.go.jp/article/internalmedicine/47/21/47_21_1925/_pdf/-char/en> | ML |
| <https://www.ncbi.nlm.nih.gov/pmc/articles/PMC1735627/pdf/v041p0e117.pdf> | ML |
| <https://www.ncbi.nlm.nih.gov/pmc/articles/PMC1735195/pdf/v039p00571.pdf> | ML |
| <http://atlasgeneticsoncology.org/Kprones/LeopardID10084.html> | ML* |
| <https://www.studyblue.com/notes/note/n/pediatric-dermatology-part-2/deck/11185956> | ML |
| <https://journals.lww.com/continuum/Fulltext/2018/02000/Neurocutaneous_Disorders.8.aspx> | ML* |
| <http://drugline.org/medic/term/leopard-syndrome> | ML |
| <http://drugline.org/medic/term/leopard-syndrome> | ML |
| <http://drugline.org/medic/term/leopard-syndrome> | ML* |
| <http://drugline.org/medic/term/leopard-syndrome> | ML |
| <https://www.ahajournals.org/doi/10.1161/CIRCULATIONAHA.108.792861> | ML* |
| <https://www.researchgate.net/publication/38043290_PTPN11_mutations_in_LEOPARD_syndrome_report_of_four_cases_in_Taiwan> | ML |
| <https://www.rcpjournals.org/content/clinmedicine/20/2/231> | ML |
| <https://escholarship.org/content/qt2d55s0t1/qt2d55s0t1.pdf> | ML |
| <https://escholarship.org/content/qt2d55s0t1/qt2d55s0t1.pdf> | ML |
| <https://escholarship.org/content/qt2d55s0t1/qt2d55s0t1.pdf> | ML |
| <https://www.karger.com/Article/Pdf/164839> | ML |
| <https://upload.wikimedia.org/wikipedia/commons/3/3a/LEOPARD_Synd2.jpg> | ML |
| <https://www.clevelandclinicmeded.com/medicalpubs/diseasemanagement/dermatology/dermatologic-signs-of-systemic-disease/> | ML |
| <https://www.sciencedirect.com/science/article/pii/S1578219015000566> | ML* |
| <https://www.sciencedirect.com/science/article/pii/S1578219015000566> | ML |
| <https://www.medicaljournals.se/acta/content/html/10.2340/00015555-2575> | ML* |
| <https://www.medicaljournals.se/acta/content/html/10.2340/00015555-2575> | ML |
| <https://www.medicaljournals.se/acta/content/html/10.2340/00015555-2575> | ML |
| <https://www.bioscience.org/2000/v5/d/stratak/fulltext.htm> | ML |
| <https://www.medicinenet.com/image-collection/multiple_lentigines_syndrome_back_picture/picture.htm> | ML |
| <https://www.medicinenet.com/image-collection/multiple_lentigines_syndrome_back_picture/picture.htm> | ML |
| <https://www.medicinenet.com/image-collection/multiple_lentigines_syndrome_back_picture/picture.htm> | ML |
| <https://www.revespcardiol.org/en-cardiac-abnormalities-in-leopard-syndrome-articulo-S1885585710000599> | ML |
| <https://www.medicinenet.com/image-collection/multiple_lentigines_syndrome_face_picture/picture.htm> | ML* |
| <https://link.springer.com/chapter/10.1007/978-1-4614-6654-3_3> | ML |
| <https://link.springer.com/chapter/10.1007/978-1-4614-6654-3_3> | ML* |
| <https://link.springer.com/chapter/10.1007/978-1-4614-6654-3_3> | ML |
| <https://www.dermatologyadvisor.com/home/decision-support-in-medicine/dermatology/cafe-au-lait-macules/> | NF1 |
| <https://www.dermatologyadvisor.com/home/decision-support-in-medicine/dermatology/cafe-au-lait-macules/> | NF1 |
| <https://www.dermatologyadvisor.com/home/decision-support-in-medicine/dermatology/cafe-au-lait-macules/> | NF1 |
| <https://www.sciencedirect.com/science/article/pii/S1578219016300853> | NF1 |
| <https://www.sciencedirect.com/science/article/pii/S1578219016300853> | NF1 |
| <https://www.sciencedirect.com/science/article/pii/S1578219016300853> | NF1 |
| <http://www.scielo.br/scielo.php?pid=S0034-72802013000200013&script=sci_arttext&tlng=en> | NF1 |
| <http://www.scielo.br/scielo.php?pid=S0034-72802013000200013&script=sci_arttext&tlng=en> | NF1 |
| <http://www.scielo.br/scielo.php?pid=S0034-72802013000200013&script=sci_arttext&tlng=en> | NF1* |
| <http://www.e-ijd.org/viewimage.asp?img=IndianJDermatol_2011_56_4_375_84721_u4.jpg> | NF1 |
| <http://www.e-ijd.org/viewimage.asp?img=IndianJDermatol_2011_56_4_375_84721_u4.jpg> | NF1 |
| <https://www.medicalhomeportal.org/diagnoses-and-conditions/neurofibromatosis-type-1> | NF1* |
| <https://www.consultant360.com/exclusives/infant-caf-au-lait-macules-vitiligo-and-hemangioma-case-neurofibromatosis-1> | NF1 |
| <https://www.webpathology.com/image.asp?case=1022&n=4> | NF1 |
| <https://www.researchgate.net/figure/Nevrofibromatosis-with-a-characteristic-cafe-au-lait-spot_fig2_256292178> | NF1* |
| <https://pedsinreview.aappublications.org/content/22/3/82/tab-figures-data> | NF1 |
| <http://www.jnsbm.org/viewimage.asp?img=JNatScBiolMed_2015_6_2_436_160029_u2.jpg> | NF1* |
| <https://entokey.com/systemic-hamartomatoses-phakomatoses/> | NF1 |
| <https://www.mdedge.com/dermatology/article/194498/pigmentation-disorders/neurofibromatosis-type-1-setting-systemic-lupus> | NF1* |
| <https://www.sciencedirect.com/science/article/pii/S0190962209012511> | NF1 |
| <https://www.sciencedirect.com/science/article/pii/S0190962209012511> | NF1 |
| <https://www.sciencedirect.com/science/article/pii/S0190962209012511> | NF1 |
| <https://www.sciencedirect.com/science/article/pii/S0190962209012511> | NF1 |
| <https://www.sciencedirect.com/science/article/pii/S0190962209012511> | NF1 |
| <https://www.sciencedirect.com/science/article/pii/S0190962209012511> | NF1 |
| <https://www.sciencedirect.com/science/article/pii/S0190962209012511> | NF1 |
| <https://www.sciencedirect.com/science/article/pii/S0190962209012511> | NF1 |
| <https://www.sciencedirect.com/science/article/pii/S0190962209012511> | NF1 |
| <https://www.consultant360.com/articles/photo-essay-hyperpigmented-macules> | NF1 |
| <https://www.consultant360.com/articles/photo-essay-hyperpigmented-macules> | NF1 |
| <https://pedsinreview.aappublications.org/content/22/3/82/tab-supplemental> | NF1 |
| <https://pedsinreview.aappublications.org/content/22/3/82/tab-supplemental> | NF1 |
| <https://www.aafp.org/afp/2012/1101/p826.html> | NF1 |
| <https://www.aafp.org/afp/2012/1101/p826.html> | NF1 |
| <https://pedsinreview.aappublications.org/content/30/5/182> | NF1 |
| <https://pedsinreview.aappublications.org/content/30/5/182> | NF1 |
| <https://www.researchgate.net/figure/Cafe-au-lait-macule-on-the-neck_fig4_274679988> | NF1 |
| <https://www.researchgate.net/figure/Huge-cafe-au-lait-macule-on-the-back_fig3_274679988> | NF1 |
| <https://emedicine.medscape.com/article/1112001-overview> | NF1* |
| <https://www.aafp.org/afp/2003/1115/p1955.html> | NF1 |
| <https://onlinelibrary.wiley.com/doi/full/10.1111/pde.13126> | NF1 |
| <https://onlinelibrary.wiley.com/doi/full/10.1111/pde.13126> | NF1 |
| <https://onlinelibrary.wiley.com/doi/full/10.1111/pde.13126> | NF1 |
| <https://clinicalgate.com/neurofibromatosis-and-tuberous-sclerosis/> | NF1 |
| <https://clinicalgate.com/neurofibromatosis-and-tuberous-sclerosis/> | NF1 |
| <https://clinicalgate.com/neurofibromatosis-and-tuberous-sclerosis/> | NF1 |
| <https://clinicalgate.com/neurofibromatosis-and-tuberous-sclerosis/> | NF1 |
| <http://www.pigmentinternational.com/showBackIssue.asp?issn=2349-5847;year=2018;volume=5;issue=1;month=January-June> | NF1 |
| <http://www.pigmentinternational.com/showBackIssue.asp?issn=2349-5847;year=2018;volume=5;issue=1;month=January-June> | NF1 |
| https://media.cheggcdn.com/media/55d/55d48fb9-90d1-4e39-88f6-cdc47a11fee6/patch1327202279170.jpg | NF1* |
| https://media.cheggcdn.com/media/55d/55d48fb9-90d1-4e39-88f6-cdc47a11fee6/patch1327202279170.jpg | NF1 |
| https://media.cheggcdn.com/media/55d/55d48fb9-90d1-4e39-88f6-cdc47a11fee6/patch1327202279170.jpg | NF1 |
| <https://dceg.cancer.gov/news-events/news/2015/doug-stewart-profile> | NF1 |
| <https://dceg.cancer.gov/news-events/news/2015/doug-stewart-profile> | NF1 |
| <https://dceg.cancer.gov/news-events/news/2015/doug-stewart-profile> | NF1 |
| <https://accesspediatrics.mhmedical.com/Content.aspx?bookid=1443&sectionid=79850513> | NF1 |
| <https://accesspediatrics.mhmedical.com/Content.aspx?bookid=1443&sectionid=79850513> | NF1 |
| <https://www.memorangapp.com/flashcards/129716/Genodermatosis/> | NF1* |
| <https://pedsinreview.aappublications.org/content/38/3/119/tab-figures-data> | NF1 |
| <https://pedsinreview.aappublications.org/content/38/3/119/tab-figures-data> | NF1 |
| <http://www.ijpd.in/viewimage.asp?img=IndianJPaediatrDermatol_2013_14_3_92_122178_f1.jpg> | NF1 |
| <https://www.verywellhealth.com/neurofibromatosis-type-1-nf1-2860837> | NF1 |
| <https://www.sciencedirect.com/topics/nursing-and-health-professions/cafe-au-lait-spot> | NF1* |
| <https://www.cureus.com/articles/12949-enlarging-plexiform-tumor-in-a-pregnant-patient-with-neurofibromatosis-type-one> | NF1 |
| <https://www.jaad.org/article/S0190-9622(15)01719-3/pdf> | NF1 |
| <https://clinicalgate.com/disorders-of-hyperpigmentation-and-melanocytes/> | NF1 |
| <https://clinicalgate.com/disorders-of-hyperpigmentation-and-melanocytes/> | NF1 |
| <https://clinicalgate.com/disorders-of-hyperpigmentation-and-melanocytes/> | NF1 |
| <https://www.touchophthalmology.com/neurofibromatosis-type-1-associated-unilateral-pediatric-glaucoma-and-proptosis-a-case-report/> | NF1 |
| <https://www.consultant360.com/articles/what-are-woman-s-widespread-asymptomatic-nodules?page=1> | NF1 |
| <https://drpanossian.com/neurofibromatosis/types-of-neurofibromatosis/neurofibromatosis-type-1/> | NF1 |
| <https://www.facebook.com/nffphilippines/photos/a.130776596963136/1378693642171419/?type=3&theater> | NF1 |
| <https://www.facebook.com/nffphilippines/photos/a.130776596963136/1378693642171419/?type=3&theater> | NF1 |
| <https://www.facebook.com/nffphilippines/photos/a.130776596963136/1378693642171419/?type=3&theater> | NF1 |
| <https://www.facebook.com/nffphilippines/photos/a.130776596963136/1378693642171419/?type=3&theater> | NF1 |
| <https://www.facebook.com/nffphilippines/photos/a.130776596963136/1378693642171419/?type=3&theater> | NF1 |
| <https://www.facebook.com/nffphilippines/photos/a.130776596963136/1378693642171419/?type=3&theater> | NF1 |
| <https://www.facebook.com/nffphilippines/photos/a.130776596963136/1378693642171419/?type=3&theater> | NF1 |
| <https://www.memorangapp.com/flashcards/189734/MS3+Neuro%3A++Neurocutaneous+Syndromes/> | NF1 |
| <https://bmcmedgenet.biomedcentral.com/articles/10.1186/s12881-018-0615-8/figures/2> | NF1 |
| <https://bmcmedgenet.biomedcentral.com/articles/10.1186/s12881-018-0615-8/figures/2> | NF1 |
| <https://bmcmedgenet.biomedcentral.com/articles/10.1186/s12881-018-0615-8/figures/2> | NF1 |
| <https://bmcmedgenet.biomedcentral.com/articles/10.1186/s12881-018-0615-8/figures/2> | NF1 |
| <https://bmcmedgenet.biomedcentral.com/articles/10.1186/s12881-018-0615-8/figures/2> | NF1 |
| <https://bmcmedgenet.biomedcentral.com/articles/10.1186/s12881-018-0615-8/figures/2> | NF1 |
| <https://www.healthplexus.net/keyword/neurofibromatosis-type-1> | NF1 |
| <https://www.healthplexus.net/keyword/neurofibromatosis-type-1> | NF1 |
| <https://medlibes.com/entry/neurofibromatosis-type-1> | NF1 |
| <https://medlibes.com/entry/neurofibromatosis-type-1> | NF1 |
| <https://www.dovepress.com/the-investigation-for-potential-modifier-genes-in-patients-with-neurof-peer-reviewed-article-OTT> | NF1* |
| <https://www.dovepress.com/the-investigation-for-potential-modifier-genes-in-patients-with-neurof-peer-reviewed-article-OTT> | NF1* |
| <https://emedicine.medscape.com/article/1112001-overview> | NF1* |
| <http://www.mjdrdypu.org/viewimage.asp?img=MedJDYPatilUniv_2016_9_1_143_168004_f1.jpg> | NF1 |
| <http://www.mjdrdypu.org/viewimage.asp?img=MedJDYPatilUniv_2016_9_1_143_168004_f1.jpg> | NF1 |
| <http://www.mjdrdypu.org/viewimage.asp?img=MedJDYPatilUniv_2016_9_1_143_168004_f1.jpg> | NF1 |
| <http://www.mjdrdypu.org/viewimage.asp?img=MedJDYPatilUniv_2016_9_1_143_168004_f1.jpg> | NF1 |
| <http://www.idoj.in/viewimage.asp?img=IndianDermatolOnlineJ_2012_3_1_51_93506_u1.jpg> | NF1 |
| <https://www.uab.edu/medicine/nfprogram/learn/neurofibromatosis-type-1-nf1/symptoms-features> | NF1 |
| <https://www.uab.edu/medicine/nfprogram/learn/neurofibromatosis-type-1-nf1/symptoms-features> | NF1 |
| <https://www.cancer.net/cancer-types/neurofibromatosis-type-1> | NF1 |
| <https://www.cancer.net/cancer-types/neurofibromatosis-type-1> | NF1 |
| <https://www.cancer.net/cancer-types/neurofibromatosis-type-1> | NF1 |
| <https://www.nfmidwest.org/wp-content/uploads/2016/03/Neurofibromas.pdf> | NF1 |
| <http://rc.rcjournal.com/content/56/11/1844/tab-pdf> | NF1 |
| http://rc.rcjournal.com/content/56/11/1844/tab-pdf | NF1 |
| <https://bestpractice.bmj.com/topics/en-us/410> | NF1 |
| https://www.ncbi.nlm.nih.gov/pmc/articles/PMC4372231/ | NF1 |
| https://www.ncbi.nlm.nih.gov/pmc/articles/PMC4372231/ | NF1 |
| <http://www.pediatricneurosciences.com/viewimage.asp?img=JPediatrNeurosci_2010_5_1_59_66684_f2.jpg> | NF1 |
| <https://www.merckmanuals.com/professional/pediatrics/neurocutaneous-syndromes/neurofibromatosis> | NF1 |
| <http://www.mrcophth.com/eyeanddermatology/neurofibromatosis.html> | NF1 |
| <http://www.mrcophth.com/eyeanddermatology/neurofibromatosis.html> | NF1 |
| <http://www.mrcophth.com/eyeanddermatology/neurofibromatosis.html> | NF1 |
| <https://www.studyblue.com/notes/note/n/neuro-goljan-audio/deck/6330973> | NF1 |
| <http://square.umin.ac.jp/nf1guideline/index.html> | NF1 |
| <http://square.umin.ac.jp/nf1guideline/index.html> | NF1 |
| <http://square.umin.ac.jp/nf1guideline/index.html> | NF1 |
| <http://square.umin.ac.jp/nf1guideline/index.html> | NF1 |
| <https://www.webconsultas.com/salud-al-dia/neurofibromatosis/sintomas-de-la-neurofibromatosis-tipo-1-nf1-7062> | NF1* |
| <https://elviento365.com/vida/3829/> | NF1 |
| <https://docplayer.nl/105041835-Poli-bijzonder-klinische-genetica-nf1-radiologie-simvastatin-nf1-simcoda-tsc-rapit-everolimus-paramedici-logopedie-navis.html> | NF1 |
| <https://docplayer.fr/111770669-Pigmentations-cutanes-de-l-enfant.html> | NF1 |
| <https://docplayer.fr/111770669-Pigmentations-cutanes-de-l-enfant.html> | NF1 |
| <https://docplayer.fr/111770669-Pigmentations-cutanes-de-l-enfant.html> | NF1 |
| <https://docplayer.com.br/82099396-Assistente-hospitalar-de-pediatria-consultant-of-pediatry-hospital-do-barreiro-centro-hospitalar-do-barreiro-montijo-barreiro-portugal-3.html> | NF1 |
| <https://www.thinglink.com/scene/447609901259161601> | NF1 |
| <https://www.neurofibromatose.fr/questions-reponses/> | NF1 |
| <https://www.neurofibromatose.fr/questions-reponses/> | NF1 |
| <https://www.neurofibromatose.fr/questions-reponses/> | NF1 |
| <https://www.neurofibromatose.fr/questions-reponses/> | NF1 |
| <https://www.researchgate.net/figure/Figuur-2-Huid-kenmerken-bij-NF1-Bij-het-kind-zijn-cafe-au-lait-vlekken-op-d-e-romp_fig2_271915261> | NF1 |
| <https://www.researchgate.net/figure/Figuur-2-Huid-kenmerken-bij-NF1-Bij-het-kind-zijn-cafe-au-lait-vlekken-op-d-e-romp_fig2_271915261> | NF1 |
| <https://www.researchgate.net/figure/Figuur-2-Huid-kenmerken-bij-NF1-Bij-het-kind-zijn-cafe-au-lait-vlekken-op-d-e-romp_fig2_271915261> | NF1 |
| <http://dermatoweb2.udl.es/ampliacio.php?idfoto=400507> | NF1 |
| <http://dermatoweb2.udl.es/ampliacio.php?idfoto=400507> | NF1 |
| <https://www.researchgate.net/figure/Figura-2-Manchas-cafe-con-leche-en-paciente-con-neurofibromatosis-tipo-1_fig3_228495366> | NF1 |
| <https://www.researchgate.net/figure/Figura-2-Manchas-cafe-con-leche-en-paciente-con-neurofibromatosis-tipo-1_fig3_228495366> | NF1 |
| <http://dermatologiaymascosas.blogspot.com/2011/11/neurofibromatosis.html> | NF1 |
| <http://dermatologiaymascosas.blogspot.com/2011/11/neurofibromatosis.html> | NF1 |
| http://dermatologiaymascosas.blogspot.com/2011/11/neurofibromatosis.html | NF1 |
| <http://endoneurocirugia.es/endoneurocirugia/divulgacion%20neurofibromatosis.Diagnostico.html> | NF1 |
| <https://www.sciencedirect.com/science/article/pii/S1761289619428173> | NF1 |
| <https://www.sciencedirect.com/science/article/pii/S1761289619428173> | NF1 |
| <https://www.sciencedirect.com/science/article/pii/S1761289619428173> | NF1 |
| <https://www.elsevier.es/es-revista-medicina-integral-63-articulo-neurofibromatosis-13015324> | NF1 |
| <https://search.ppsimages.co.jp/cgi-bin/search.cgi?rm=results&site=Meta&do_search=1&form_name=metageneral_and_metahistory&per_page=60&keyword_and=%E3%83%AC%E3%83%83%E3%82%AF%E3%83%AA%E3%83%B3%E3%82%B0%E3%83%8F%E3%82%A6%E3%82%BC%E3%83%B3%E7%97%85> | NF1* |
| <https://www.webmd.com/skin-problems-and-treatments/picture-of-neurofibromatosis> | NF1 |
| <https://www.semanticscholar.org/paper/Neurofibromatosis-type-I-(von-Recklinghausen%E2%80%99s-A-of-Kosti%C4%87-Dini%C4%87/f0be7d005feeab487287e66a807e2d1d42854676/figure/2> | NF1 |
| <https://www.semanticscholar.org/paper/Neurofibromatosis-type-I-(von-Recklinghausen%E2%80%99s-A-of-Kosti%C4%87-Dini%C4%87/f0be7d005feeab487287e66a807e2d1d42854676/figure/2> | NF1 |
| <https://www.semanticscholar.org/paper/Neurofibromatosis-type-I-(von-Recklinghausen%E2%80%99s-A-of-Kosti%C4%87-Dini%C4%87/f0be7d005feeab487287e66a807e2d1d42854676/figure/0> | NF1 |
| <https://link.springer.com/article/10.2165/00128071-200809010-00007> | NF1 |
| <https://link.springer.com/article/10.2165/00128071-200809010-00007> | NF1 |
| <https://somepomed.org/articulos/contents/mobipreview.htm?26/36/27211> | NF1 |
| <https://somepomed.org/articulos/contents/mobipreview.htm?26/36/27211> | NF1 |
| <https://somepomed.org/articulos/contents/mobipreview.htm?26/36/27211> | NF1 |
| <https://somepomed.org/articulos/contents/mobipreview.htm?26/36/27211> | NF1* |
| <https://www.pinterest.com/pin/99923685454392600/> | NF1 |
| <https://www.pinterest.com/pin/99923685454392600/> | NF1 |
| <https://www.pinterest.com/pin/99923685454392600/> | NF1 |
| <https://www.memorangapp.com/flashcards/189734/MS3+Neuro%3A++Neurocutaneous+Syndromes/> | NF1 |
| <https://www.uab.edu/medicine/news/latest/item/956-uab-researchers-work-to-unravel-the-complex-genetic-disease-neurofibromatosis-type-1> | NF1 |
| <https://www.uab.edu/medicine/news/latest/item/956-uab-researchers-work-to-unravel-the-complex-genetic-disease-neurofibromatosis-type-1> | NF1 |
| <https://www.uab.edu/medicine/news/latest/item/956-uab-researchers-work-to-unravel-the-complex-genetic-disease-neurofibromatosis-type-1> | NF1 |
| <https://quizlet.com/gb/161129334/paces-skin-2-neurofibromatosis-flash-cards/> | NF1 |
| <https://surgicalcasereports.springeropen.com/articles/10.1186/s40792-015-0107-4> | NF1* |
| <https://surgicalcasereports.springeropen.com/articles/10.1186/s40792-015-0107-4> | NF1* |
| <https://www.jaypeedigital.com/book/9788184482843/chapter/ch10> | NF1 |
| <https://www.scielo.br/scielo.php?script=sci_arttext&pid=S0004-282X2014000300241> | NF1 |
| <https://www.scielo.br/scielo.php?script=sci_arttext&pid=S0004-282X2014000300241> | NF1 |
| <https://www.scielo.br/scielo.php?script=sci_arttext&pid=S0004-282X2014000300241> | NF1 |
| <https://medihelp.life/neurofibromatosis-type-i-von-recklinghausens-disease-a-family-case-report-and-literature-review/> | NF1 |
| <https://medihelp.life/neurofibromatosis-type-i-von-recklinghausens-disease-a-family-case-report-and-literature-review/> | NF1 |
| <https://medihelp.life/neurofibromatosis-type-i-von-recklinghausens-disease-a-family-case-report-and-literature-review/> | NF1 |
| <https://medihelp.life/neurofibromatosis-type-i-von-recklinghausens-disease-a-family-case-report-and-literature-review/> | NF1 |
| <https://jnnp.bmj.com/content/66/4/417> | NF1 |
| <https://plasticsurgerykey.com/the-neurofibromatoses/> | NF1 |
| <https://plasticsurgerykey.com/the-neurofibromatoses/> | NF1 |
| <http://ceo.medword.net/?signs=neurofibromatosis-type-1-nf-1-skin-scapula-alata> | NF1 |
| <http://ceo.medword.net/?signs=neurofibromatosis-type-1-nf-1-skin-scapula-alata> | NF1 |
| <https://www.scielo.br/scielo.php?pid=S0365-05962013000300329&script=sci_arttext&tlng=en> | NF1 |
| <https://www.mitchmedical.us/physical-diagnosis/info-cza.html> | NF1 |
| <https://www.atlasophthalmology.net/photo.jsf;jsessionid=32004E0FCAB78FAB81C1692F508D50EB?node=9826&locale=de> | NF1 |
| <https://www.sciencedirect.com/science/article/pii/S1761289619428173> | NF1 |
| <https://www.sciencedirect.com/science/article/pii/S1761289619428173> | NF1 |
| <https://www.sciencedirect.com/science/article/pii/S1761289610703277> | NF1 |
| <https://obgynkey.com/disorders-of-hyperpigmentation-and-melanocytes/> | NF1 |
| [https://obgynkey.com/disorders-of-hyperpigmentation-and-melanocytes/](https://obgynkey.com/selected-hereditary-diseases/) | NF1 |
| [https://obgynkey.com/disorders-of-hyperpigmentation-and-melanocytes/](https://obgynkey.com/selected-hereditary-diseases/) | NF1 |
| <https://obgynkey.com/selected-hereditary-diseases/> | NF1 |
| <https://obgynkey.com/selected-hereditary-diseases/> | NF1 |
| <https://obgynkey.com/selected-hereditary-diseases/> | NF1 |
| <https://yixueshu.gitee.io/nelson/text00218.html> | NF1 |
| <https://yixueshu.gitee.io/nelson/text00218.html> | NF1 |
| <https://yixueshu.gitee.io/nelson/text00218.html> | NF1 |
| <https://yixueshu.gitee.io/nelson/text00218.html> | NF1 |
| <http://www.pcds.org.uk/clinical-guidance/neurofibromatosis-type-1> | NF1 |
| <http://www.pcds.org.uk/clinical-guidance/neurofibromatosis-type-1> | NF1 |
| <https://www.youtube.com/watch?v=Rh0ejpGasp0> | NF1 |
| <http://scielo.isciii.es/scielo.php?pid=S1130-05582008000300005&script=sci_arttext&tlng=en> | NF1 |
| <https://neurofibromatose.nl/kenmerken-van-nf1/huid-en-zwellingen> | NF1 |
| <https://neurofibromatose.nl/kenmerken-van-nf1/huid-en-zwellingen> | NF1 |
| <https://neurofibromatose.nl/kenmerken-van-nf1/huid-en-zwellingen> | NF1 |
| <https://neurofibromatose.nl/kenmerken-van-nf1/huid-en-zwellingen> | NF1 |
| <https://www.huidziekten.nl/zakboek/dermatosen/ntxt/Neurofibromatose.htm> | NF1 |
| <https://www.huidziekten.nl/zakboek/dermatosen/ntxt/Neurofibromatose.htm> | NF1 |
| <https://www.huidziekten.nl/zakboek/dermatosen/ntxt/Neurofibromatose.htm> | NF1 |
| <https://www.huidziekten.nl/zakboek/dermatosen/ntxt/Neurofibromatose.htm> | NF1 |
| <https://www.huidziekten.nl/zakboek/dermatosen/ntxt/Neurofibromatose.htm> | NF1 |
| <https://www.huidziekten.nl/zakboek/dermatosen/ntxt/Neurofibromatose.htm> | NF1 |
| <https://www.huidziekten.nl/zakboek/dermatosen/ntxt/Neurofibromatose.htm> | NF1 |
| <https://www.huidziekten.nl/zakboek/dermatosen/ntxt/Neurofibromatose.htm> | NF1 |
| <https://www.huidziekten.nl/zakboek/dermatosen/ntxt/Neurofibromatose.htm> | NF1 |
| <https://www.huidziekten.nl/zakboek/dermatosen/ntxt/Neurofibromatose.htm> | NF1 |
| <https://www.huidziekten.nl/zakboek/dermatosen/ntxt/Neurofibromatose.htm> | NF1 |
| <https://www.huidziekten.nl/zakboek/dermatosen/ntxt/Neurofibromatose.htm> | NF1 |
| <https://www.huidziekten.nl/zakboek/dermatosen/ntxt/Neurofibromatose.htm> | NF1 |
| <https://www.huidziekten.nl/zakboek/dermatosen/ntxt/Neurofibromatose.htm> | NF1 |
| <https://www.kinderaerztliche-praxis.de/a/neurofibromatose-typ-und-typ-1693419> | NF1 |
| <https://www.kinderaerztliche-praxis.de/a/neurofibromatose-typ-und-typ-1693419> | NF1 |
| <https://link.springer.com/article/10.1007/s00105-019-4416-6> | NF1 |
| <https://link.springer.com/article/10.1007/s00105-019-4416-6> | NF1 |
| <https://link.springer.com/article/10.1007/s00105-019-4416-6> | NF1 |
| <https://link.springer.com/article/10.1007/s00105-019-4416-6> | NF1 |
| <https://link.springer.com/article/10.1007/s00105-019-4416-6> | NF1 |
| <https://pubs.rsna.org/doi/full/10.1148/rg.252045176> | NF1 |
| <https://link.springer.com/referenceworkentry/10.1007%2F978-3-642-02202-9_148> | NF1 |
| <https://link.springer.com/referenceworkentry/10.1007%2F978-3-642-02202-9_148> | NF1 |
| <https://link.springer.com/referenceworkentry/10.1007%2F978-3-642-02202-9_148> | NF1 |
| <https://www.longdom.org/open-access/its-not-mccunealbright-syndrome-its-neurofibromatosis1-2157-7412-1000i101.pdf> | NF1* |
| <https://www.longdom.org/open-access/its-not-mccunealbright-syndrome-its-neurofibromatosis1-2157-7412-1000i101.pdf> | NF1 |
| <https://www.longdom.org/open-access/its-not-mccunealbright-syndrome-its-neurofibromatosis1-2157-7412-1000i101.pdf> | NF1 |
| <https://www.dermatologyadvisor.com/home/decision-support-in-medicine/dermatology/cafe-au-lait-macules/> | NF1* |
| <https://link.springer.com/article/10.1007/s00381-020-04758-5#Fig9> | NF1 |
| <https://www.sciencedirect.com/science/article/pii/S0738081X14003137?via%3Dihub#f0005> | NF1* |
| <https://www.ncbi.nlm.nih.gov/pmc/articles/PMC2716546/> | NF1 |
| <https://www.ncbi.nlm.nih.gov/pmc/articles/PMC2716546/> | NF1 |
| <https://www.nature.com/articles/gim20101> | NF1 |
| <https://www.nature.com/articles/gim20101> | NF1 |
| <https://www.thelancet.com/journals/lancet/article/PIIS0140-6736(20)31909-7/fulltext?rss=yes> | NF1 |
| <https://www.thelancet.com/journals/lancet/article/PIIS0140-6736(20)31909-7/fulltext?rss=yes> | NF1 |
| <https://pediatrics.aappublications.org/content/143/5/e20190660/tab-figures-data> | NF1 |
| <https://pediatrics.aappublications.org/content/143/5/e20190660/tab-figures-data> | NF1 |
| <https://www.longdom.org/open-access/autosomal-dominant-diseases-are-too-often-overlooked-in-the-parents-of-affected-children-report-of-six-cases-2157-7412.1000190.pdf> | NF1 |
| <https://www.longdom.org/open-access/autosomal-dominant-diseases-are-too-often-overlooked-in-the-parents-of-affected-children-report-of-six-cases-2157-7412.1000190.pdf> | NF1 |
| <https://link.springer.com/chapter/10.1007/978-3-030-50823-4_11> | NF1 |
| <https://link.springer.com/chapter/10.1007/978-3-030-50823-4_11> | NF1 |
| <https://link.springer.com/chapter/10.1007/978-3-030-50823-4_11> | NF1 |
| <https://www.consultant360.com/exclusives/infant-caf-au-lait-macules-vitiligo-and-hemangioma-case-neurofibromatosis-1> | NF1 |
| <https://journals.lww.com/continuum/Fulltext/2018/02000/Neurocutaneous_Disorders.8.aspx> | NF1 |
| <https://journals.lww.com/continuum/Fulltext/2018/02000/Neurocutaneous_Disorders.8.aspx> | NF1 |
| <https://plasticsurgerykey.com/ash-leaf-macules/> | TSC |
| https://plasticsurgerykey.com/ash-leaf-macules/ | TSC |
| <https://medlineplus.gov/ency/imagepages/2538.htm> | TSC |
| <https://www.merckmanuals.com/en-ca/home/children-s-health-issues/neurocutaneous-syndromes-in-children/tuberous-sclerosis-complex> | TSC |
| <http://makaayo.blogspot.com/2012/03/ash-leaf-spots-new-treatments-excellent_07.html> | TSC |
| <https://www.memorangapp.com/flashcards/91586/Neurologic+Systems/> | TSC |
| <https://www.memorangapp.com/flashcards/91586/Neurologic+Systems/> | TSC |
| <https://www.memorangapp.com/flashcards/91586/Neurologic+Systems/> | TSC |
| <https://www.researchgate.net/publication/258062315_Everolimus_in_the_treatment_of_subependymal_giant_cell_astrocytomas_angiomyolipomas_and_pulmonary_and_skin_lesions_associated_with_tuberous_sclerosis_complex/figures?lo=1> | TSC* |
| <https://step1.medbullets.com/neurology/113051/tuberous-sclerosis> | TSC |
| <https://step1.medbullets.com/neurology/113051/tuberous-sclerosis> | TSC |
| <https://obgynkey.com/selected-hereditary-diseases/> | TSC |
| <https://www.bjmp.org/content/skin-and-seizures-tuberous-sclerosis-complex-pictorial-essay> | TSC |
| <https://step2.medbullets.com/neurology/120327/tuberous-sclerosis> | TSC |
| <https://www.aafp.org/afp/2007/0401/p1053.html> | TSC |
| <https://doctorlib.info/medical/fitzpatrick-atlas-dermatology/19.html> | TSC |
| <https://doctorlib.info/medical/fitzpatrick-atlas-dermatology/19.html> | TSC |
| <https://doctorlib.info/medical/fitzpatrick-atlas-dermatology/19.html> | TSC |
| <http://www.idoj.in/article.asp?issn=2229-5178;year=2015;volume=6;issue=2;spage=142;epage=143;aulast=George> | TSC |
| <https://www.jaypeedigital.com/book/9789351526278/chapter/ch7> | TSC* |
| <https://www.huidziekten.nl/quiz/dermatologiequiz134qd.htm> | TSC* |
| <https://obgynkey.com/pigmentary-development-of-east-asian-skin/> | TSC* |
| <https://www.kinderaerztliche-praxis.de/a/tsc-tuberoese-sklerose-komplex-klinisches-bild-1984470> | TSC |
| <https://deximed.de/home/b/neurologie/krankheiten/erbkrankheiten/tuberoese-sklerose/> | TSC |
| <https://link.springer.com/article/10.1007/s00105-008-1633-9> | TSC* |
| <https://www.sciencedirect.com/science/article/pii/S1761289605442685> | TSC |
| <https://www.tuasaude.com/es/manchas-blancas-en-la-piel/> | TSC |
| <https://www.tuasaude.com/es/manchas-blancas-en-la-piel/> | TSC |
| <https://www.massgeneral.org/neurology/tsc/patient-stories/> | TSC |
| <https://twitter.com/harkerdavid/status/1138976907255582721> | TSC |
| <https://twitter.com/harkerdavid/status/1138976907255582721> | TSC |
| <https://pulpbits.net/6-ash-leaf-macules-pictures/ash-leaf-macules-tuberous-sclerosis/> | TSC |
| <https://pulpbits.net/6-ash-leaf-macules-pictures/ash-leaf-macules-tuberous-sclerosis/> | TSC |
| <http://dermchallenge.blogspot.com/2012/07/ash-leaf-macule-of-tuberous-sclerosis.html> | TSC |
| <https://www.researchgate.net/figure/Age-distribution-of-tuberous-sclerosis-patients-in-our-study_fig4_232351019> | TSC |
| <http://what-when-how.com/acp-medicine/coetaneous-manifestations-of-systemic-diseases-part-4/> | TSC |
| <https://www.memorangapp.com/flashcards/41902/Genetic+Diseases+pictures/> | TSC* |
| <https://www.dermcoll.edu.au/atoz/tuberous-sclerosis-complex/> | TSC |
| <http://swisstscnetwork.ch/wp-content/uploads/tsc-flyer-english-21.06.2016.pdf> | TSC |
| <https://www.sciencedirect.com/science/article/pii/S0975962X13000713#fig3> | TSC |
| <https://www.sciencedirect.com/science/article/pii/S0975962X13000713#fig3> | TSC* |
| <https://www.researchgate.net/figure/Multiple-small-hypopigmented-confetti-macules-on-abdomen-and-upper-limbs-in-a-young-boy_fig3_41453655> | TSC |
| <https://www.dovepress.com/clinical-features-epidemiology-and-therapy-of-lymphangioleiomyomatosis-peer-reviewed-fulltext-article-CLEP> | TSC |
| <https://www.dovepress.com/clinical-features-epidemiology-and-therapy-of-lymphangioleiomyomatosis-peer-reviewed-fulltext-article-CLEP> | TSC |
| <https://www.pediatriconcall.com/articles/genetics/tuberous-sclerosis/tuberous-sclerosis-patient-education> | TSC |
| <https://www.pediatriconcall.com/articles/genetics/tuberous-sclerosis/tuberous-sclerosis-patient-education> | TSC |
| <https://www.wikidoc.org/index.php/Tuberous_sclerosis_history_and_symptoms> | TSC* |
| <http://www.angelfire.com/la3/tsc/pictpage_ashleaf.html> | TSC |
| <https://studyhippo.com/dermatology-pathology-slides-from-lecture/> | TSC |
| <https://studyhippo.com/dermatology-pathology-slides-from-lecture/> | TSC |
| <https://www.jaypeedigital.com/book/9789385891212/chapter/ch47> | TSC |
| <https://www.jaypeedigital.com/book/9789385891212/chapter/ch47> | TSC |
| <http://www.pcds.org.uk/clinical-guidance/tuberous-sclerosis> | TSC* |
| <http://www.ijo.in/article.asp?issn=0301-4738;year=2019;volume=67;issue=3;spage=433;epage=435;aulast=Rajasekaran> | TSC |
| <https://pedsinreview.aappublications.org/content/38/3/119> | TSC* |
| <https://www.dermatologyadvisor.com/home/decision-support-in-medicine/dermatology/tuberous-sclerosis-complex-bourneville-disease-bourneville-phakomatosis/> | TSC |
| <https://www.dermatologyadvisor.com/home/decision-support-in-medicine/dermatology/tuberous-sclerosis-complex-bourneville-disease-bourneville-phakomatosis/> | TSC |
| <https://www.dermatologyadvisor.com/home/decision-support-in-medicine/dermatology/tuberous-sclerosis-complex-bourneville-disease-bourneville-phakomatosis/> | TSC |
| <https://www.dermatologyadvisor.com/home/decision-support-in-medicine/dermatology/tuberous-sclerosis-complex-bourneville-disease-bourneville-phakomatosis/> | TSC |
| <https://www.scielo.br/scielo.php?script=sci_arttext&pid=S0365-05962012000200001> | TSC* |
| <https://www.scielo.br/scielo.php?script=sci_arttext&pid=S0365-05962012000200001> | TSC |
| <https://www.scielo.br/scielo.php?script=sci_arttext&pid=S0365-05962012000200001> | TSC* |
| <http://www.ijdvl.com/article.asp?issn=0378-6323;year=2015;volume=81;issue=1;spage=97;epage=101;aulast=Viswanath> | TSC |
| <https://www.researchgate.net/figure/Ash-leave-macules-and-shagreen-patch-in-the-back-in-a-child-with-tuberous-sclerosis_fig10_273389178> | TSC |
| <http://www.ijpd.in/viewimage.asp?img=IndianJPaediatrDermatol_2019_20_3_219_261870_f7.jpg> | TSC |
| <https://dermnetnz.org/topics/tuberous-sclerosis/> | TSC |
| <https://dermnetnz.org/topics/tuberous-sclerosis/> | TSC |
| <https://dermnetnz.org/topics/tuberous-sclerosis/> | TSC |
| <https://dermnetnz.org/topics/tuberous-sclerosis/> | TSC |
| <https://obgynkey.com/tuberous-sclerosis/> | TSC |
| <https://obgynkey.com/tuberous-sclerosis/> | TSC |
| <https://obgynkey.com/tuberous-sclerosis/> | TSC |
| <https://obgynkey.com/tuberous-sclerosis/> | TSC |
| <https://www.medicalhomeportal.org/diagnoses-and-conditions/tuberous-sclerosis-complex> | TSC |
| <https://www.researchgate.net/publication/325905613_Tuberous_sclerosis_complex_Review_based_on_new_diagnostic_criteria> | TSC |
| <https://www.researchgate.net/publication/325905613_Tuberous_sclerosis_complex_Review_based_on_new_diagnostic_criteria> | TSC |
| <https://www.researchgate.net/publication/325905613_Tuberous_sclerosis_complex_Review_based_on_new_diagnostic_criteria> | TSC |
| <https://www.researchgate.net/publication/325905613_Tuberous_sclerosis_complex_Review_based_on_new_diagnostic_criteria> | TSC |
| <https://www.researchgate.net/publication/325905613_Tuberous_sclerosis_complex_Review_based_on_new_diagnostic_criteria> | TSC |
| <https://radiologykey.com/pulmonary-manifestations-of-dermatologic-diseases/> | TSC* |
| <https://www.contemporarypediatrics.com/view/hypopigmented-patches-infant-history-seizures> | TSC |
| <https://www.contemporarypediatrics.com/view/hypopigmented-patches-infant-history-seizures> | TSC |
| <https://www.contemporarypediatrics.com/view/hypopigmented-patches-infant-history-seizures> | TSC |
| <https://www.contemporarypediatrics.com/view/hypopigmented-patches-infant-history-seizures> | TSC |
| <https://www.jaypeedigital.com/book/9788184485851/chapter/ch18> | TSC |
| <https://www.jaypeedigital.com/book/9788184485851/chapter/ch18> | TSC |
| <https://www.jaypeedigital.com/book/9788184485851/chapter/ch18> | TSC |
| <https://www.pinterest.com/pin/145100419217716273/> | TSC |
| <https://link.springer.com/referenceworkentry/10.1007%2F978-1-4939-2401-1_237> | TSC |
| <https://link.springer.com/referenceworkentry/10.1007%2F978-1-4939-2401-1_237> | TSC |
| <https://link.springer.com/referenceworkentry/10.1007%2F978-1-4939-2401-1_237> | TSC |
| <https://link.springer.com/referenceworkentry/10.1007%2F978-1-4939-2401-1_237> | TSC |
| <https://link.springer.com/referenceworkentry/10.1007%2F978-1-4939-2401-1_237> | TSC |
| <https://www.pinterest.com/pin/556194622703901836/> | TSC |
| <https://www.consultant360.com/articles/what-s-cause-girl-s-cutaneous-lesions> | TSC |
| <https://www.consultant360.com/articles/what-s-cause-girl-s-cutaneous-lesions> | TSC |
| <https://www.clinicaladvisor.com/home/dermatologic-look-alikes/infants-with-hypopigmented-processes/3/> | TSC |
| <https://www.bestonlinemd.com/ash-leaf-spots-new-treatments-excellent-health-information/> | TSC |
| <https://www.bestonlinemd.com/ash-leaf-spots-new-treatments-excellent-health-information/> | TSC |
| <https://www.scielo.br/pdf/abd/v88n2/0365-0596-abd-88-2-0303.pdf> | TSC |
| <https://www.cram.com/flashcards/uworld-5-13-2194534> | TSC |
| <https://www.slideserve.com/marnina/phacomatoses-powerpoint-ppt-presentation> | TSC |
| <https://journals.lww.com/continuum/Fulltext/2018/02000/Neurocutaneous_Disorders.8.aspx> | TSC* |
| <https://www.slideshare.net/IbrahimMohammed15/tuberous-sclerosis-complex-tsc> | TSC |
| <https://dermnetnz.org/topics/tuberous-sclerosis/> | TSC |
| <https://www.sciencedirect.com/science/article/pii/B9780444627025000068?via%3Dihub> | TSC* |
| <https://www.nature.com/articles/nrdp201635> | TSC* |
| <https://www.sciencedirect.com/science/article/pii/S1474442215000691?via%3Dihub> | TSC |
| <https://www.sciencedirect.com/science/article/pii/S1474442215000691?via%3Dihub> | TSC |
| <https://pubmed.ncbi.nlm.nih.gov/24053982/> | TSC |
| <https://pubmed.ncbi.nlm.nih.gov/24053982/> | TSC |
| <https://pubmed.ncbi.nlm.nih.gov/24053982/> | TSC |
| <https://casereports.bmj.com/content/2018/bcr-2018-224923> | TSC |
| <https://casereports.bmj.com/content/2018/bcr-2018-224923> | TSC |
| <https://www.revistanefrologia.com/en-tsc2-pkd1-contiguous-gene-syndrome-articulo-S2013251417301840#imagen-1> | TSC* |
| <https://pubmed.ncbi.nlm.nih.gov/4968649/> | TSC |
| <https://avgopleiding.nl/wp-content/uploads/2020/04/Tubereuze-Sclerose-Complex-samenvatting.pdf> | TSC |
| <https://avgopleiding.nl/wp-content/uploads/2020/04/Tubereuze-Sclerose-Complex-samenvatting.pdf> | TSC |
| <https://avgopleiding.nl/wp-content/uploads/2020/04/Tubereuze-Sclerose-Complex-samenvatting.pdf> | TSC |
| <https://www.sciencedirect.com/science/article/pii/S0140673608612799?via%3Dihub> | TSC |
| <https://reader.elsevier.com/reader/sd/pii/S0031395515000255?token=F2B3846504424E648DDB5D099A01BCDD76644E057F639332127DFF7F0BD46F2098C42CB09B7954166C28008CE2AFA440> | TSC |
| <https://reader.elsevier.com/reader/sd/pii/S0031395515000255?token=F2B3846504424E648DDB5D099A01BCDD76644E057F639332127DFF7F0BD46F2098C42CB09B7954166C28008CE2AFA440> | TSC |
| <https://onlinelibrary.wiley.com/doi/full/10.1111/pde.12567> | TSC |
| <https://link.springer.com/article/10.1007%2Fs00508-015-0758-y> | TSC |
| <https://link.springer.com/article/10.1007%2Fs00508-015-0758-y> | TSC |
| <https://onlinelibrary.wiley.com/doi/full/10.1111/1346-8138.14349> | TSC |
| <https://www.sciencedirect.com/science/article/pii/S0891524506002768?via%3Dihub#fig1> | TSC |
| <https://www.sciencedirect.com/science/article/pii/S0891524506002768?via%3Dihub#fig1> | TSC |
| <https://journals.sagepub.com/doi/pdf/10.1177/08830738040190090301> | TSC |
| <https://www.sciencedirect.com/science/article/pii/S0190962207008420?via%3Dihub#fig2> | TSC |
| <https://www.sciencedirect.com/science/article/pii/S0887899415303805?via%3Dihub#fig2> | TSC* |
| <https://www.ncbi.nlm.nih.gov/pmc/articles/PMC4307053/> | TSC |
| <https://jpma.org.pk/PdfDownload/2233> | TSC |
| <https://jpma.org.pk/article-details/2233?article_id=2233> | TSC |
| <https://www.mediasphera.ru/issues/zhurnal-nevrologii-i-psikhiatrii-im-s-s-korsakova/2015/10/downloads/ru/241997-729820151018> | TSC |
| <https://www.mediasphera.ru/issues/zhurnal-nevrologii-i-psikhiatrii-im-s-s-korsakova/2015/10/downloads/ru/241997-729820151018> | TSC |
| <https://www.ncbi.nlm.nih.gov/pmc/articles/PMC6531247/> | TSC* |
| <https://www.ncbi.nlm.nih.gov/pmc/articles/PMC6531247/> | TSC |
| <https://www.ncbi.nlm.nih.gov/pmc/articles/PMC6531247/> | TSC |
| <https://www.sciencedirect.com/science/article/pii/S107190910600012X?via%3Dihub#fig5> | TSC |
| <https://link.springer.com/article/10.1007%2Fs12017-015-8354-x> | TSC |
| <https://www.sciencedirect.com/science/article/pii/S0929693X10002800?via%3Dihub#fig0010> | TSC* |
| <https://www.dermatologyadvisor.com/home/decision-support-in-medicine/dermatology/cafe-au-lait-macules/> | other* |
| <https://www.dermatologyadvisor.com/home/decision-support-in-medicine/dermatology/cafe-au-lait-macules/> | other |
| <https://www.dermatologyadvisor.com/home/decision-support-in-medicine/dermatology/cafe-au-lait-macules/> | other |
| <https://onlinelibrary.wiley.com/doi/full/10.1111/pde.12936> | other |
| <https://onlinelibrary.wiley.com/doi/full/10.1111/pde.12936> | other |
| <https://onlinelibrary.wiley.com/doi/full/10.1111/pde.12936> | other |
| <https://community.whattoexpect.com/forums/may-2014-babies/topic/cafe-au-lait-spots-15.html?page=2> | other |
| <https://www.sciforschenonline.org/journals/clinical-cosmetic-dermatology/article-data/JCCD-1-111/JCCD-1-111.pdf> | other |
| <https://community.whattoexpect.com/forums/july-2018-babies/topic/cafe-au-lait-spots-72327107.html> | other |
| <https://community.whattoexpect.com/forums/july-2018-babies/topic/cafe-au-lait-spots-72327107.html> | other |
| <https://community.whattoexpect.com/forums/july-2018-babies/topic/cafe-au-lait-spots-72327107.html> | other |
| <https://www.sciencedirect.com/science/article/pii/S0190962299700757> | other* |
| <https://www.sciencedirect.com/science/article/pii/S0190962299700757> | other |
| <https://juniperpublishers.com/oajnn/images/OAJNN.MS.ID.555622.G001.png> | other |
| <https://www.fanconi.org/images/uploads/other/Alter_FA101_FA_Adult_2014_for_FARF_small.pdf> | other |
| <https://link.springer.com/article/10.1007/s00431-014-2349-8> | other |
| <https://www.consultant360.com/articles/photo-essay-hyperpigmented-macules> | other |
| <https://www.consultant360.com/articles/photo-essay-hyperpigmented-macules> | other |
| <https://quizlet.com/66940617/test> | other* |
| <https://www.aafp.org/afp/2017/1215/p797.html> | other |
| <https://www.aafp.org/afp/2017/1215/p797.html> | other |
| <https://www.childrens.com/specialties-services/conditions/congenital-nevus> | other |
| <http://medcomhk.com/hkdvb/details.asp?id=837&show=1234> | other |
| <http://medcomhk.com/hkdvb/details.asp?id=837&show=1234> | other |
| <https://www.researchgate.net/figure/Changes-of-patients-cafe-au-lait-macules-during-treatment-periods_fig1_318731439> | other |
| <https://casereports.bmj.com/content/11/1/bcr-2018-226171> | other |
| <https://pedsinreview.aappublications.org/content/39/11/e50> | other* |
| <https://www.consultant360.com/content/does-brown-patch-boy%E2%80%99s-buttock-signal-underlying-illness> | other |
| <https://indianpediatrics.net/aug2005/aug-831.htm> | other |
| <https://celibre.com/ba-birthmark-removal-7/> | other |
| <https://www.mdedge.com/clinicianreviews/quiz/7078/dermatology/macules-and-melanocytes> | other |
| <https://www.clinicaladvisor.com/slideshow/derm-dx/derm-dx-hyperpigmented-macule-on-the-abdomen/> | other |
| <http://www.idoj.in/article.asp?issn=2229-5178;year=2015;volume=6;issue=3;spage=230;epage=231;aulast=Nair> | other |
| <https://finance.yahoo.com/news/babys-rare-heart-shaped-birthmark-160804090.html> | other |
| <https://forefrontdermatology.com/consider-birthmark-removal/> | other |
| <https://blog.pregistry.com/birthmark-something-worse/> | other |
| <https://www.researchgate.net/figure/A-pigmented-macule-of-5-mm-on-the-dorsum-of-the-left-foot_fig1_45283071> | other |
| <https://www.researchgate.net/figure/Unilateral-golden-brown-macule-on-left-ankle-Presence-of-purpuric-dots-within-the-lesion_fig1_237059559> | other |
| <http://www.aworldonthebrink.com/blog/a-small-black-macule-apparently-unchanged> | other |
| <https://www.netdoctor.co.uk/conditions/heart-and-blood/a1173/aplastic-anaemia/> | other |
| <https://www.sciencedirect.com/topics/nursing-and-health-professions/cafe-au-lait-spot> | other |
| <https://link.springer.com/chapter/10.1007/978-3-319-33919-1_60> | other |
| <https://link.springer.com/chapter/10.1007/978-3-319-33919-1_60> | other |
| <https://link.springer.com/chapter/10.1007/978-3-319-33919-1_60> | other |
| <https://link.springer.com/chapter/10.1007/978-3-319-33919-1_60> | other |
| <https://link.springer.com/chapter/10.1007/978-3-319-33919-1_60> | other |
| <https://link.springer.com/chapter/10.1007/978-3-319-33919-1_60> | other |
| <https://link.springer.com/chapter/10.1007/978-3-319-33919-1_60> | other |
| <https://link.springer.com/chapter/10.1007/978-3-319-33919-1_60> | other |
| <https://link.springer.com/chapter/10.1007/978-3-319-33919-1_60> | other |
| <https://link.springer.com/chapter/10.1007/978-3-319-33919-1_60> | other |
| <https://www.sciencedirect.com/science/article/pii/S0190962207002009> | other |
| <https://obgynkey.com/disorders-of-hyperpigmentation-and-melanocytes/> | other |
| <https://obgynkey.com/disorders-of-hyperpigmentation-and-melanocytes/> | other |
| <https://www.nhs.uk/conditions/birthmarks/> | other |
| <https://www.nhs.uk/conditions/birthmarks/> | other |
| <https://www.nhs.uk/conditions/birthmarks/> | other* |
| <https://www.nhs.uk/conditions/birthmarks/> | other |
| <https://www.juliahartskinclinic.co.uk/pigmentation> | other* |
| <https://www.webmd.com/skin-problems-and-treatments/ss/slideshow-birthmarks> | other |
| <https://www.webmd.com/skin-problems-and-treatments/ss/slideshow-birthmarks> | other* |
| <https://www.parent24.com/Baby/Babycare/more-than-a-birthmark-my-familys-experience-of-infant-hemangioma-20180514> | other |
| <https://pdskin.com/service/laser-removal-of-vascular-birthmarks/premier-dermatology-before-after-port-wine-birthmark/> | other |
| <https://www.todayifoundout.com/index.php/2011/10/where-birthmarks-come-from/> | other |
| <https://factdr.com/health-conditions/birthmarks/> | other |
| <https://factdr.com/health-conditions/birthmarks/> | other |
| <https://www.laserskincare.com.au/conditions/birth-marks/> | other |
| <https://torontodermatologycentre.com/vascular-birthmarks/> | other |
| <https://torontodermatologycentre.com/vascular-birthmarks/> | other |
| <https://www.sciencephoto.com/media/148458/view/strawberry-birthmark-on-the-scalp> | other |
| <https://nextshark.com/what-exactly-is-a-mongolian-spot-anyway/> | other |
| <https://www.africaparent.com/what-is-a-birthmark> | other* |
| <https://www.mayoclinic.org/birthmarks/sls-20076683?s=4> | other |
| <https://en.wikipedia.org/wiki/Melanoma> | other |
| <https://sgsmn.com/condition/melanomaskin-cancer/> | other |
| <https://www.aad.org/public/diseases/skin-cancer/types/common/scc/symptoms> | other |
| <http://dermcast.tv/live-blog-nonmelanoma-skin-cancer-basal-cell-carcinoma-and-squamous-cell-carcinoma-scott-dinehart-md/> | other |
| <https://www.foryourbestself.com/squamous-cell-carcinoma-melbourne/> | other |
| <https://www.everydayhealth.com/hpv/guide/symptoms/> | other |
| <https://en.wikipedia.org/wiki/Molluscum_contagiosum> | other |
| [https://www.medicalnewstoday.com/articles/325189](https://www.medicalnewstoday.com/articles/325189.php) | other |
| <https://www.mirror.co.uk/news/uk-news/mums-arm-rots-away-after-14239858> | other |
| <https://www.shutterstock.com/search/birthmark> | other |
| <https://www.webmd.com/skin-problems-and-treatments/ss/slideshow-age-related-growths> | other* |
| <https://universityhealthnews.com/daily/cancer/age-spots-or-signs-of-skin-cancer/> | other |
| <https://www.cancercouncil.com.au/skin-cancer/the-signs-of-skin-cancer/> | other |
| <https://www.cancercouncil.com.au/skin-cancer/the-signs-of-skin-cancer/> | other |
| <https://www.cancercouncil.com.au/skin-cancer/the-signs-of-skin-cancer/> | other |
| <https://www.cancercouncil.com.au/skin-cancer/the-signs-of-skin-cancer/> | other |
| <https://www.cancercouncil.com.au/skin-cancer/the-signs-of-skin-cancer/> | other |
| <https://www.cancercouncil.com.au/skin-cancer/the-signs-of-skin-cancer/> | other |
| <https://www.cancercouncil.com.au/skin-cancer/the-signs-of-skin-cancer/> | other |
| <https://skincheck.com.au/skinclinic/age-spots/> | other |
| <https://skincheck.com.au/skinclinic/age-spots/> | other |
| <https://skincheck.com.au/skinclinic/seborrheic-keratosis/> | other |
| <https://skincheck.com.au/skinclinic/iec/> | other |
| <https://www.oatext.com/ringworm-on-the-legs-of-a-patient-after-long-term-intensive-care-unit-residency.php> | other |
| <https://www.sciencesource.com/archive/Ringworm--Tinea-corporis--SS2579340.html> | other |
| <https://www.sciencesource.com/archive/Ringworm--Tinea-corporis--SS2579340.html> | other |
| <https://www.webmd.com/skin-problems-and-treatments/ss/slideshow-lumps-bumps-skin> | other |
| <https://www.cancer.org/latest-news/how-to-spot-skin-cancer.html> | other |
| <https://tnoncology.com/news/2019/06/15/how-to-spot-skin-cancer/> | other |
| <https://tnoncology.com/news/2019/06/15/how-to-spot-skin-cancer/> | other |
| <https://www.healthline.com/health/skin-disorders/blue-nevus> | other |
| <https://www.medicalnewstoday.com/articles/317999.php> | other |
| <http://theconversation.com/common-skin-rashes-and-what-to-do-about-them-91518> | other |
| <http://theconversation.com/common-skin-rashes-and-what-to-do-about-them-91518> | other |
| <http://theconversation.com/common-skin-rashes-and-what-to-do-about-them-91518> | other |
| <https://www.medicalnewstoday.com/articles/319488.php> | other |
| <https://www.mdedge.com/clinicianreviews/article/87306/dermatology/unexpected-effect-crohns> | other |
| <https://www.cliniciansbrief.com/article/image-gallery-primary-skin-lesions> | other |
| <https://www.mdedge.com/dermatology/article/167458/pediatrics/red-brown-plaque-leg> | other |
| <https://www.healthline.com/health/morphea> | other |
| <https://nanopdf.com/download/taking-a-history-amp-terminology-dr-iain-henderson-gp-4_pdf> | other |
| <https://www.britishskinfoundation.org.uk/bcc> | other |
| <https://www.merckmanuals.com/home/quick-facts-skin-disorders/skin-cancers/basal-cell-carcinoma> | other |
| <https://www.healthline.com/health/basal-cell-carcinoma> | other |
| <https://www.healthline.com/health/basal-cell-carcinoma> | other |
| <https://en.wikipedia.org/wiki/Bruise> | other |
| <https://health.usnews.com/wellness/articles/2016-06-13/should-you-be-worried-about-bruising-easily> | other |
| <https://www.hellodoctor.co.za/the-causes-and-treatment-for-constant-bruising/> | other |
| <https://www.istockphoto.com/search/more-like-this/1157387369?assettype=image&family=creative&mediatype=photography> | other |
| <https://www.justlivewell.com/bruising-bleeding-gums-and-strokes/> | other |
| <https://www.thehealthy.com/first-aid/get-rid-of-bruises-faster/> | other |
| <https://www.boredpanda.com/little-superhero-barman-mask-birthmark-natalie-jackson/?utm_source=google&utm_medium=organic&utm_campaign=organic> | other |
| <https://abcnews.go.com/Lifestyle/actresss-facial-birthmark-celebrated-gorgeous-photo-series/story?id=38540434> | other |
| <https://www.tyla.com/life/real-life-mum-reveals-abuse-she-received-over-daughters-rare-birthmark-20190501> | other |
| <https://californiaskininstitute.com/conditions/birthmarks/> | other |
| <https://www.everydayhealth.com/kids-health-pictures/10-types-of-birthmarks.aspx> | other |
| <https://www.everydayhealth.com/kids-health-pictures/10-types-of-birthmarks.aspx> | other |
| <https://www.everydayhealth.com/kids-health-pictures/10-types-of-birthmarks.aspx> | other |
| <https://www.everydayhealth.com/kids-health-pictures/10-types-of-birthmarks.aspx> | other |
| <https://www.everydayhealth.com/kids-health-pictures/10-types-of-birthmarks.aspx> | other |
| <https://www.everydayhealth.com/kids-health-pictures/10-types-of-birthmarks.aspx> | other |
| <https://www.express.co.uk/life-style/life/673083/mum-hits-back-at-strangers-who-ask-about-son-s-birthmark> | other |
| <https://www.bbc.com/news/av/uk-44226184/mum-helps-stop-daughter-being-bullied-for-birthmark> | other |
| <https://www.mdedge.com/clinicianreviews/article/170615/dermatology/birthmark-ipelago> | other |
| <https://www.healthychildren.org/English/news/Pages/Preventing-Problems-From-Baby-Birthmark.aspx> | other |
| <http://www.joycelim.com/medical/birth-marks> | other |
| <http://www.joycelim.com/medical/birth-marks> | other |
| <https://fineartamerica.com/art/birthmark> | other |
| <https://birthmark.org/birthmark/port-wine-stain/> | other |
| <https://www.medicalnewstoday.com/articles/326201.php> | other |
| <https://commons.wikimedia.org/wiki/File:Skin_black_mole_on_the_face.JPG> | other |
| <https://www.express.co.uk/life-style/health/1156850/skin-cancer-signs-symptoms-melanoma-mole-pictures> | other |
| <https://www.delphineleemd.com/basal-cell-carcinoma> | other |
| <https://www.cancertherapyadvisor.com/home/cancer-topics/skin-cancer/can-vismodegib-induce-melanoma-in-patients-treated-for-basal-cell-carcinoma/> | other |
| <https://www.omicsonline.org/china/basal-cell-carcinoma-peer-reviewed-pdf-ppt-articles/> | other |
| <https://www.lymedisease.org/lyme-basics/lyme-disease/early-lyme/> | other |
| <https://www.skinsite.com/info_lyme_disease.htm> | other |
| <https://clinicalconnection.hopkinsmedicine.org/news/deep-machine-learning-can-more-accurately-identify-erythema-migrans-rashes-in-early-lyme-disease> | other |
| <https://www.express.co.uk/life-style/health/1197904/lyme-disease-symptoms-neck-stiffness-tick-rash-bite> | other |
| <https://www.nhpr.org/post/officials-2014-likely-record-year-lyme-disease#stream/0> | other |
| <http://www.independentnurse.co.uk/clinical-article/the-management-of-lyme-disease/146823/> | other |
| <https://www.pbs.org/newshour/science/4-things-know-ticks-lyme-disease-summer> | other |
| <https://www.innatoss.com/en/lyme-disease/symptoms/bullseye-rash/> | other |
| <https://radio.wosu.org/post/study-prolonged-antibiotic-treatment-gave-no-relief-lasting-lyme-symptoms#stream/0> | other* |
| <https://www.whattoexpect.com/toddler/childhood-injuries/treating-spider-bites.aspx> | other |
| <https://www.theindusparent.com/mongolian-blue-spots-babies-need-know-information> | other |
| <https://dermboard.org/birthmarks/mongolian-spot/> | other |
| <https://paulkenny.me/2014/01/09/mongolian-blue-spots/> | other* |
| <https://www.amaskincare.com/what-is-a-birthmark-does-removal-work/> | other |
| <https://www.medicinenet.com/image-collection/mongolian_spots_picture/picture.htm> | other |
| <https://www.medicinenet.com/image-collection/nevus_depigmentosus_picture/picture.htm> | other |
| <https://adc.bmj.com/content/early/2019/07/26/archdischild-2019-317497> | other |
| <https://adc.bmj.com/content/early/2019/07/26/archdischild-2019-317497> | other |
| <https://adc.bmj.com/content/early/2019/07/26/archdischild-2019-317497> | other |
| <https://www.healthline.com/health/birthmarks-red> | other |
| [https://skindelhi.com/where-does-a-birthmark-come-from/](http://skindelhi.com/where-does-a-birthmark-come-from/) | other |
| https://emedicine.medscape.com/article/1083849-clinical | other |
| <https://www.huffingtonpost.co.uk/2016/08/31/ringworm-in-babies-and-children-treatment-and-prevention_n_7355350.html?guccounter=1&guce_referrer=aHR0cHM6Ly93d3cuZ29vZ2xlLmNvbS8&guce_referrer_sig=AQAAAJ5n3qoGMLGN59x6jFAI4T8tLVhh217UHtGo5zruvSNuPcefCo_sf1WVVI1QKyGsvstdpmYcoQOHc_fCsSKHo9wy7IGNvIVX_Yn443yj_4164GeFS_Z00hIeweQYTmeaWR_Nk1O28cNEcAlDXmsx1LuMomGdl4seK0pMkHznaG-W> | other |
| <https://menstrual-cycle-calculator.com/ringworm-in-babies-causes-prevention-symptoms-treatment/> | other |
| <https://www.medicalnewstoday.com/articles/315502> | other |
| <http://www.myskinmychoice.com/app.php?name=irash> | other |
| <https://www.usdermatologypartners.com/service/keloids-scar-treatment/> | other |
| <https://xstrahl.com/xia-optimizing-radiotherapy-for-keloids/> | other |
| <https://www.quora.com/How-much-does-keloid-treatment-cost-in-India> | other |
| <https://pixels.com/featured/keloid-scar-gary-parkerscience-photo-library.html> | other |
| <https://www.sciencesource.com/archive/Close-up-of-a-keloid-scar-developed-after-surgery-SS2174675.html> | other |
| <https://www.livescience.com/51109-melanoma-mole-deadliness.html> | other |
| <https://steemit.com/cancer/@scooter77/melanoma-the-silent-killer> | other |
| <https://www.researchgate.net/figure/Patient-at-7-years-of-age-with-persistence-of-extensive-Mongolian-spot-and-superimposed_fig2_335819744> | other |
| <https://www.semanticscholar.org/paper/Blue-nevus-with-a-starburst-pattern-on-dermoscopy-Shiga-Nakajima/dbb02a35a17bf5b98d6fc1966afc7d68efa1c694> | other |
| <https://escholarship.org/uc/item/9694x4mp> | other |
| <http://dermoscopic.blogspot.com/2008/04/> | other |
| <http://dermoscopic.blogspot.com/2008/04/> | other |
| <http://dermoscopic.blogspot.com/2008/04/> | other |
| <http://dermoscopic.blogspot.com/2008/04/> | other |
| <https://dermnetnz.org/topics/blue-naevus-dermoscopy/> | other |
| <https://dermnetnz.org/topics/blue-naevus-dermoscopy/> | other |
| <https://dermnetnz.org/topics/blue-naevus-dermoscopy/> | other |
| <https://dermnetnz.org/topics/blue-naevus-dermoscopy/> | other |
| <https://dermnetnz.org/topics/blue-naevus-dermoscopy/> | other |
| <https://dermnetnz.org/topics/blue-naevus-dermoscopy/> | other |
| <https://dermnetnz.org/topics/blue-naevus-dermoscopy/> | other |
| <https://dermnetnz.org/topics/blue-naevus-dermoscopy/> | other |
| <https://dermnetnz.org/topics/blue-naevus-dermoscopy/> | other |
| <https://dermnetnz.org/topics/basal-cell-carcinoma-affecting-the-trunk-images/> | other |
| <https://www.scielo.br/scielo.php?script=sci_arttext&pid=S0004-282X2014000300241> | other |
| <https://www.scielo.br/scielo.php?script=sci_arttext&pid=S0004-282X2014000300241> | other |
| <https://www.mitchmedical.us/physical-diagnosis/info-cza.html> | other |
| <https://goldskincare.com/vitiligo/> | other* |
| <https://www.medicinenet.com/vitiligo/article.htm> | other* |
| <https://www.medicinenet.com/image-collection/vitiligo_neck_picture/picture.htm> | other* |
| <https://www.medicinenet.com/image-collection/depigmented_patch_of_skin_picture/picture.htm> | other |
| <http://www.pcds.org.uk/quick-guide/hypopigmentation-white-or-lighter-areas-of-skin-colour#!prettyPhoto> | other |
| <http://www.pcds.org.uk/quick-guide/hypopigmentation-white-or-lighter-areas-of-skin-colour#!prettyPhoto> | other |
| <http://www.pcds.org.uk/quick-guide/hypopigmentation-white-or-lighter-areas-of-skin-colour#!prettyPhoto> | other |
| <https://obgynkey.com/disorders-of-hyperpigmentation-and-melanocytes/> | other |
| <https://obgynkey.com/disorders-of-hyperpigmentation-and-melanocytes/> | other |
| <https://www.newriverdermatology.com/blog/vitiligo-awareness> | other* |
| <https://magazine.medlineplus.gov/article/treating-vitiligo-studies-look-for-long-term-options> | other |
| <https://www.cosmopolitan.com/uk/beauty-hair/a19494259/vitiligo-photo-series-instagram/> | other |
| <https://www.cosmopolitan.com/uk/beauty-hair/a19494259/vitiligo-photo-series-instagram/> | other |
| https://www.aboutkidshealth.ca/Article?contentid=2296&language=English | other* |
| <https://www.aboutkidshealth.ca/Article?contentid=2296&language=English> | other |
| <https://plasticsurgerykey.com/109-melanocytic-naevi-and-melanoma/> | other* |
| <https://plasticsurgerykey.com/109-melanocytic-naevi-and-melanoma/> | other |
| <https://plasticsurgerykey.com/109-melanocytic-naevi-and-melanoma/> | other |
| <https://www.sciencedirect.com/science/article/pii/B9780323462020000108> | other |
| <https://neoreviews.aappublications.org/content/suppl/2003/10/01/4.10.e263.DC1> | other* |
| <https://neoreviews.aappublications.org/content/suppl/2003/10/01/4.10.e263.DC1> | other |
| <https://www.dermatologyadvisor.com/home/decision-support-in-medicine/dermatology/beckers-nevus-beckers-melanosis-beckers-pigmentary-hamartoma-nevoid-melanosis-pigmented-hairy-epidermal-nevus/> | other |
| <https://cdn.mdedge.com/files/s3fs-public/Document/September-2017/073050311.pdf> | other* |
| <https://plasticsurgerykey.com/melanocytic-proliferations-and-other-pigmented-lesions/> | other |
| <http://www.jcasonline.com/viewimage.asp?img=JCutanAesthetSurg_2013_6_2_65_112665_f5.jpg> | other |
| <http://www.jcasonline.com/viewimage.asp?img=JCutanAesthetSurg_2013_6_2_65_112665_f5.jpg> | other |
| <http://www.jcasonline.com/viewimage.asp?img=JCutanAesthetSurg_2013_6_2_65_112665_f5.jpg> | other |
| <https://www.aocd.org/page/HaloMoles> | other |
| <https://link.springer.com/chapter/10.1007/978-3-319-03218-4_73> | other |
| <https://perridermatology.com/melanocytic-nevi-halo-nevus/> | other |
| <https://www.sciencephoto.com/media/616947/view/nevi-with-and-without-halo> | other |
| <https://www.sciencedirect.com/topics/nursing-and-health-professions/halo-nevus> | other |
| <https://somepomed.org/articulos/contents/mobipreview.htm?6/53/6996> | other |
| <https://www.researchgate.net/figure/Compound-nevus-Fig-2837-Suttons-halo-nevus_fig20_312930545> | other |
| <https://www.wikidoc.org/index.php/Halo_nevus> | other |
| <https://www.wikidoc.org/index.php/File:Naevus_halo02.jpg> | other |
| [https://pubmed.ncbi.nlm.nih.gov/24986573/](https://www.semanticscholar.org/paper/Multiple-halo-naevi-associated-with-tocilizumab.-Kuet-Goodfield/fc747f1e0cd8b7d2393382409a0f160a9e0b16e5/figure/1) | other |
| <https://dermateam.nl/aandoeningen/halo-moedervlek/> | other |
| <https://www.ilmelanoma.com/en/moles/moles/meyerson-naevus-2/> | other |
| <https://www.sciencesource.com/archive/Inflamed-compound-nevus--mole--SS2480991.html> | other |
| <https://www.huidziekten.nl/zakboek/dermatosen/ntxt/NaevusSuttonHaloNevus.htm> | other |
| <https://www.huidziekten.nl/afbeeldingen/halo-naevus-3.jpg> | other |
| <https://www.healthline.com/health/nevus> | other |
| <https://www.cancertherapyadvisor.com/home/decision-support-in-medicine/pediatrics/common-birthmarks/> | other |
| <https://www.cancertherapyadvisor.com/home/decision-support-in-medicine/pediatrics/common-birthmarks/> | other |
| <http://relmuisverjagen.blogspot.com/2014/05/witte-vlekken.html> | other* |
| <https://www.huidziekten.nl/afbeeldingen/piebaldisme-3.jpg> | other |
| <https://www.davinciclinic.be/aandoeningen/halo-moedervlek/> | other |
| <https://www.davinciclinic.be/aandoeningen/halo-moedervlek/> | other |
| <https://link.springer.com/referenceworkentry/10.1007%2F978-3-642-02202-9_148> | other |
| <https://link.springer.com/referenceworkentry/10.1007%2F978-3-642-02202-9_148> | other |
| <https://link.springer.com/referenceworkentry/10.1007%2F978-3-642-02202-9_148> | other |
| <https://plasticsurgerykey.com/benign-melanocytic-neoplasms/> | other |
| <http://what-when-how.com/acp-medicine/coetaneous-manifestations-of-systemic-diseases-part-4/> | other |
| <https://europepmc.org/article/pmc/pmc6536047> | other |
| <https://twitter.com/JAADjournals/status/1009518006941175808> | other |
| <https://srlotion.wordpress.com/tag/ichthyosis-treatment/> | other |
| <http://www.pcds.org.uk/quick-guide/trunk-buttocks-limbs-brown-black-blue-grey-flat-more-than-1-cm-diameter> | other |
| <http://www.pcds.org.uk/quick-guide/trunk-buttocks-limbs-brown-black-blue-grey-flat-more-than-1-cm-diameter#!prettyPhoto> | other |
| <https://www.umassmed.edu/vitiligo/blog/blog-posts1/2020/01/maybe-its-not-vitiligo/> | other |
| <https://www.umassmed.edu/vitiligo/blog/blog-posts1/2020/01/maybe-its-not-vitiligo/> | other |
| <https://www.umassmed.edu/vitiligo/blog/blog-posts1/2020/01/maybe-its-not-vitiligo/> | other |
| <https://doctorlib.info/pediatric/visual-diagnosis-treatment-pediatrics/73.html> | other |
