## Supplemental Table 3 for "Proof-of-principle neural network models for classification, attribution, creation, style-mixing, and morphing of image data for genetic conditions"

**Supplemental Table 3.** URLs to images used for disease progression in Neurofibromatosis type 1 (NF1) generated via a generative adversarial network (GAN) (links are subject to change). NF1_early, NF_intermediate, and NF1_late refer to the disease stages, which were based on visual review of cases by both a clinical geneticist and a genetic counselor using perdescribed age-related manifestations^1^ .

| URL | Condition |
| --- | --- |
| <https://dceg.cancer.gov/news-events/news/2015/doug-stewart-profile> | NF1_early |
| <https://www.mdedge.com/dermatology/article/47520/pediatrics/multiple-calms-infancy-may-signal-nf1?sso=true> | NF1_early |
| <https://www.sciencedirect.com/science/article/pii/B9780323401395001078> | NF1_early |
| <https://www.healthplexus.net/keyword/neurofibromatosis-type-1> | NF1_early |
| <https://www.researchgate.net/figure/Clinical-manifestations-of-patients-with-neurofibromatosis-type-1-in-the-Chinese-family_fig3_304808475> | NF1_early |
| <https://www.skillsplatform.org/courses/7634-neurofibromatosis-type-1-nf1-free-online-nursing-and-gp-cpd-courses> | NF1_early |
| <https://www.uab.edu/news/research/item/6935-uab-researchers-work-to-unravel-the-complex-genetic-disease-neurofibromatosis-type-1> | NF1_early |
| <http://www.pcds.org.uk/clinical-guidance/neurofibromatosis-type-1> | NF1_early |
| <https://www.researchgate.net/publication/235931471_Autoimmune_diseases_associated_with_neurofibromatosis_type_1_NF1> | NF1_early |
| <https://link.springer.com/referenceworkentry/10.1007%2F978-1-4939-2401-1_178> | NF1_early |
| <https://www.semanticscholar.org/paper/The-diagnostic-and-clinical-significance-of-Shah/c32bfce58c3cbde418dcd548ad1c8a528a6635df/figure/4> | NF1_early |
| <https://www.researchgate.net/figure/Cafe-au-lait-patches-in-neurofibromatosis-type-1-child_fig6_273389178> | NF1_early |
| <https://www.epilepsy.com/learn/epilepsy-due-specific-causes/structural-causes-epilepsy/specific-structural-epilepsies/neurofibromatosis-type-1> | NF1_early |
| <https://illnessee.com/neurofibromatosis-baby-pictures/> | NF1_early |
| <https://www.actasdermo.org/en-an-update-on-neurofibromatosis-type-articulo-S1578219016300853> | NF1_early |
| <https://www.scielo.br/scielo.php?script=sci_arttext&pid=S0365-05962013000300329> | NF1_early |
| <https://www.researchgate.net/figure/Figuur-2-Huid-kenmerken-bij-NF1-Bij-het-kind-zijn-cafe-au-lait-vlekken-op-d-e-romp_fig2_271915261> | NF1_early |
| <https://link.springer.com/referenceworkentry/10.1007%2F978-3-642-02202-9_148> | NF1_early |
| <https://media.cheggcdn.com/media/55d/55d48fb9-90d1-4e39-88f6-cdc47a11fee6/patch1327202279170.jpg> | NF1_early |
| <https://plasticsurgerykey.com/the-neurofibromatoses/> | NF1_early |
| <https://link.springer.com/chapter/10.1007/978-3-030-50823-4_11> | NF1_early |
| <https://eurjmedres.biomedcentral.com/articles/10.1186/2047-783X-14-3-102> | NF1_early |
| <https://www.karger.com/Article/FullText/346643> | NF1_early |
| <https://rjme.ro/RJME/resources/files/600219713716.pdf> | NF1_early |
| <https://rjme.ro/RJME/resources/files/600219713716.pdf> | NF1_early |
| <https://www.actasdermo.org/es-pdf-S0001731016000740> | NF1_early |
| <https://www.medicalhomeportal.org/diagnoses-and-conditions/neurofibromatosis-type-1> | NF1_early |
| <https://www.kidspot.com.au/parenting/real-life/reader-stories/we-dont-know-what-the-future-holds-for-lennox/news-story/f2eb3dde6d62a6017d5ce57e96907adb> | NF1_early |
| <https://www.wjgnet.com/2307-8960/full/v8/i22/WJCC-8-5678-g002.htm> | NF1_early |
| <https://www.epilepsy.com/learn/epilepsy-due-specific-causes/structural-causes-epilepsy/specific-structural-epilepsies/neurofibromatosis-type-1> | NF1_early |
| <https://www.touchophthalmology.com/glaucoma/journal-articles/neurofibromatosis-type-1-associated-unilateral-pediatric-glaucoma-and-proptosis-a-case-report/> | NF1_early |
| <https://www.resmedjournal.com/article/S0954-6111(19)30011-3/fulltext> | NF1_early |
| <https://www.researchgate.net/figure/Case-one-photograph-of-cafe-au-lait-spots_fig2_276885900> | NF1_early |
| <https://bestpractice.bmj.com/topics/en-us/410> | NF1_early |
| <https://www.pediatricneurosciences.com/viewimage.asp?img=JPediatrNeurosci_2010_5_1_59_66684_f2.jpg> | NF1_early |
| <https://www.amboss.com/us/knowledge/Neurocutaneous_syndromes> | NF1_early |
| <http://www.pifukezazhi.com/EN/volumn/volumn_412.shtml> | NF1_early |
| <https://radiologykey.com/pulmonary-manifestations-of-dermatologic-diseases/> | NF1_early |
| <https://www.msdmanuals.com/home/multimedia/image/v39825289> | NF1_early |
| <https://www.dermatologyadvisor.com/home/decision-support-in-medicine/dermatology/neurofibromatosis-von-recklinghausen-disease-central-neurofibromatosis-schwannomatosis-neurilemmomatosis/> | NF1_early |
| <https://www.researchgate.net/profile/Retno_Danarti/publication/278026297_Neurofibromatosis_tipe_1_Manifestasi_Dermatologis_Neurologis_dan_Psikiatris/links/55f7cd3608aec948c473e1e8/Neurofibromatosis-tipe-1-Manifestasi-Dermatologis-Neurologis-dan-Psikiatris.pdf> | NF1_early |
| <https://images.app.goo.gl/aL1x7URotqUNR9DB6> | NF1_early |
| <https://www.orthobullets.com/pediatrics/4054/neurofibromatosis> | NF1_early |
| <https://pubmed.ncbi.nlm.nih.gov/31703719/#&gid=article-figures&pid=fig-2-uid-1> | NF1_early |
| <https://ijponline.biomedcentral.com/articles/10.1186/1824-7288-39-10/figures/1> | NF1_early |
| <https://www.ncbi.nlm.nih.gov/pmc/articles/PMC6293151/> | NF1_early |
| <https://www.semanticscholar.org/paper/The-diagnostic-and-clinical-significance-of-Shah/c32bfce58c3cbde418dcd548ad1c8a528a6635df/figure/4> | NF1_early |
| <https://www.researchgate.net/figure/Patient-2-Multiple-cafe-au-lait-spots-on-the-back-and-patches-of-alopecia-areata-on-the_fig1_235931471> | NF1_early |
| <https://www.cancerjournal.net/article.asp?issn=0973-1482;year=2015;volume=11;issue=2;spage=498;epage=499;aulast=Incecik> | NF1_early |
| <https://www.e-ijd.org/article.asp?issn=0019-5154;year=2015;volume=60;issue=6;spage=573;epage=577;aulast=Chander> | NF1_early |
| <https://www.consultant360.com/articles/boy-hyperpigmented-patch-and-caf-au-lait-macules> | NF1_early |
| <https://www.consultant360.com/articles/boy-hyperpigmented-patch-and-caf-au-lait-macules> | NF1_early |
| <https://www.touchophthalmology.com/glaucoma/journal-articles/neurofibromatosis-type-1-associated-unilateral-pediatric-glaucoma-and-proptosis-a-case-report/> | NF1_early |
| <https://www.jaad.org/article/S0190-9622(09)01251-1/fulltext> | NF1_early |
| <https://www.jaad.org/article/S0190-9622(09)01251-1/fulltext> | NF1_early |
| <https://nfnetwork.org/data/uploads/pdfs/primary-care_for_patients.pdf> | NF1_early |
| <https://www.sciencedirect.com/science/article/pii/S2212555817301308#fig0005> | NF1_early |
| <https://pedsinreview.aappublications.org/content/22/3/82/tab-figures-data> | NF1_early |
| <https://pedsinreview.aappublications.org/content/22/3/82/tab-figures-data> | NF1_early |
| <https://bmcmedgenet.biomedcentral.com/articles/10.1186/s12881-018-0615-8> | NF1_early |
| <https://www.jle.com/en/revues/ejd/e-docs/neurofibromatosis_type_1_and_x_linked_ichthyosis_in_a_patient_with_a_novel_frameshift_mutation_in_the_nf1_gene_307456/article.phtml> | NF1_early |
| <https://docserv.uni-duesseldorf.de/servlets/DerivateServlet/Derivate-2353/353.pdf> | NF1_early |
| <https://www.gesundheit.de/lexika/medizin-lexikon/cafe-au-lait-fleck> | NF1_early |
| <https://www.actasdermo.org/en-rasopatias-trastornos-del-desarrollo-con-articulo-S1578219011000163> | NF1_early |
| <http://www.pediatric-orthopedics.com/Topics/Diseases/Neurofibromatosis/neurofibromatosis.html> | NF1_early |
| <http://ceo.medword.net/?signs=neurofibromatosis-type-1-nf-1-skin-scapula-alata> | NF1_early |
| <https://link.springer.com/article/10.1007/s11033-019-04888-3/figures/2> | NF1_early |
| <https://www.researchgate.net/figure/Signs-of-NF1-in-the-index-patient-of-family-2-a-c-CALS-in-the-right-inguinal-region_fig4_5990058> | NF1_early |
| <https://link.springer.com/referenceworkentry/10.1007%2F978-1-4939-2401-1_178> | NF1_early |
| <https://link.springer.com/referenceworkentry/10.1007%2F978-1-4939-2401-1_178> | NF1_early |
| <https://link.springer.com/referenceworkentry/10.1007%2F978-1-4939-2401-1_178> | NF1_early |
| <https://link.springer.com/referenceworkentry/10.1007%2F978-1-4939-2401-1_178> | NF1_early |
| <https://plasticsurgerykey.com/cafe-au-lait-macules-calms/> | NF1_early |
| <https://neupsykey.com/neurocutaneous-syndromes-3/> | NF1_early |
| <https://www.blackandbrownskin.co.uk/abdomen/caf-au-lait-neurofibromatosis-type-11> | NF1_early |
| <https://www.dermatologyadvisor.com/home/decision-support-in-medicine/dermatology/neurofibromatosis-von-recklinghausen-disease-central-neurofibromatosis-schwannomatosis-neurilemmomatosis/> | NF1_early |
| <https://juniperpublishers.com/jcmah/pdf/JCMAH.MS.ID.555650.pdf> | NF1_early |
| <https://www.scielo.br/scielo.php?script=sci_arttext&pid=S1807-59322013001201521> | NF1_early |
| <https://www.hss.edu/conditions_neurofibromatosis-complications-scoliosis-tibial-dysplasia.asp> | NF1_early |
| <https://www.nf1andpninfo.com/> | NF1_early |
| <https://onlinelibrary.wiley.com/doi/10.1002/ajmg.a.33045> | NF1_early |
| <https://www.paediatricsandchildhealthjournal.co.uk/article/S1751-7222(11)00034-5/fulltext> | NF1_early |
| <https://www.scielo.br/scielo.php?script=sci_arttext&pid=S0365-05962013000300329> | NF1_early |
| <https://swansonfamilyblog.com/2019/07/16/what-is-neurofibromatosis-type-1/> | NF1_early |
| <https://slideplayer.com/slide/12775160/> | NF1_early |
| <https://www.researchgate.net/figure/A-B-A-neurofibroma-filling-the-external-ear-canal-and-pushing-the-auricle-forward-C_fig1_316257338> | NF1_early |
| <https://www.nfaireland.ie/wp-content/uploads/2017/03/3-The-Child-With-NF1.pdf> | NF1_early |
| <https://jamanetwork.com/journals/jamapediatrics/fullarticle/516339> | NF1_early |
| <https://neoreviews.aappublications.org/content/16/1/e26> | NF1_early |
| <https://www.ijo.in/viewimage.asp?img=IndianJOphthalmol_2008_56_2_161_39128_1.jpg> | NF1_early |
| <https://www.jisppd.com/viewimage.asp?img=JIndianSocPedodPrevDent_2007_25_1_30_31987_3.jpg> | NF1_early |
| <https://www.orthopaedicsone.com/display/Review/Neurofibromatosis> | NF1_early |
| <https://www.scielo.br/scielo.php?pid=S0034-72802013000200013&script=sci_arttext&tlng=en> | NF1_early |
| <https://onlinelibrary.wiley.com/doi/full/10.1111/1346-8138.13169> | NF1_early |
| <https://onlinelibrary.wiley.com/doi/full/10.1111/1346-8138.13169> | NF1_early |
| <https://onlinelibrary.wiley.com/doi/full/10.1111/1346-8138.13169> | NF1_early |
| <https://onlinelibrary.wiley.com/doi/full/10.1111/1346-8138.13169> | NF1_early |
| <https://onlinelibrary.wiley.com/doi/full/10.1111/1346-8138.13169> | NF1_early |
| <https://onlinelibrary.wiley.com/doi/full/10.1111/1346-8138.13169> | NF1_early |
| <https://onlinelibrary.wiley.com/doi/full/10.1111/1346-8138.13169> | NF1_early |
| <http://www.clinicaterapeutica.it/2020/171/5/02_MIRAGLIA.pdf> | NF1_early |
| <https://www.ncbi.nlm.nih.gov/pmc/articles/PMC2716546/> | NF1_early |
| <https://journals.lww.com/jaaos/Fulltext/2010/06000/Orthopaedic_Manifestations_of_Neurofibromatosis.7.aspx> | NF1_early |
| <https://reader.elsevier.com/reader/sd/pii/S0248866304002437?token=D35F73468FF63B917EFB31D27F3694FE5D6E6E995C477C5B604CE9D5A1987F916B2243F2F10A5F287B50913FB78BA1D2> | NF1_early |
| <http://xbyxb.csu.edu.cn/xbwk/fileup/PDF/201807811.pdf> | NF1_early |
| <https://www.ncbi.nlm.nih.gov/pmc/articles/PMC5786411/pdf/abd-92-06-0870.pdf> | NF1_early |
| <https://jamanetwork.com/journals/jamadermatology/fullarticle/554238> | NF1_early |
| <https://www.e-ijd.org/viewimage.asp?img=IndianJDermatol_2011_56_4_375_84721_u4.jpg> | NF1_intermediate |
| <https://eurjmedres.biomedcentral.com/articles/10.1186/2047-783X-19-17> | NF1_intermediate |
| <http://www.pediatric-orthopedics.com/Topics/Diseases/Neurofibromatosis/neurofibromatosis.html> | NF1_intermediate |
| https://www.researchgate.net/publication/323297080_The_investigation_for_potential_modifier_genes_in_patients_with_neurofibromatosis_type_1_based_on_next-generation_sequencing/figures | NF1_intermediate |
| <https://www.aappublications.org/news/2019/04/22/neurofibromatosis042219> | NF1_intermediate |
| <https://www.sciencedirect.com/science/article/pii/S1578219016300853> | NF1_intermediate |
| <https://bmcophthalmol.biomedcentral.com/articles/10.1186/s12886-020-01438-5> | NF1_intermediate |
| <https://www.tandfonline.com/doi/pdf/10.3109/17453678609156819> | NF1_intermediate |
| <https://pedsinreview.aappublications.org/content/30/5/182> | NF1_intermediate |
| <https://www.dermatologyadvisor.com/home/decision-support-in-medicine/dermatology/neurofibromatosis-von-recklinghausen-disease-central-neurofibromatosis-schwannomatosis-neurilemmomatosis/> | NF1_intermediate |
| <http://www.mjdrdypu.org/viewimage.asp?img=MedJDYPatilUniv_2016_9_1_143_168004_f1.jpg> | NF1_intermediate |
| <http://www.idoj.in/viewimage.asp?img=IndianDermatolOnlineJ_2012_3_1_51_93506_u1.jpg> | NF1_intermediate |
| <https://link.springer.com/chapter/10.1007/978-3-030-50823-4_11> | NF1_intermediate |
| <https://pediatrics.aappublications.org/content/121/3/633> | NF1_intermediate |
| <https://bmcmedgenet.biomedcentral.com/articles/10.1186/s12881-018-0615-8/figures/2> | NF1_intermediate |
| <https://bmcmedgenet.biomedcentral.com/articles/10.1186/s12881-018-0615-8/figures/2> | NF1_intermediate |
| <https://bmcmedgenet.biomedcentral.com/articles/10.1186/s12881-018-0615-8/figures/2> | NF1_intermediate |
| <https://www.consultant360.com/articles/what-are-woman-s-widespread-asymptomatic-nodules?page=1> | NF1_intermediate |
| <https://www.ncbi.nlm.nih.gov/pmc/articles/PMC4917078/> | NF1_intermediate |
| <https://www.ncbi.nlm.nih.gov/pmc/articles/PMC4917078/> | NF1_intermediate |
| <https://ejcrp.org/article.asp?issn=2311-3006%3Byear=2020%3Bvolume=7%3Bissue=2%3Bspage=90%3Bepage=94%3Baulast=Chiu%3Btype=3> | NF1_intermediate |
| <https://www.sciencedirect.com/science/article/pii/S0140673620319097?via%3Dihub#fig1> | NF1_intermediate |
| <http://rc.rcjournal.com/content/56/11/1844> | NF1_intermediate |
| <https://content.sciendo.com/view/journals/bjmg/21/2/article-p45.xml?product=sciendo> | NF1_intermediate |
| <https://content.sciendo.com/view/journals/bjmg/21/2/article-p45.xml?product=sciendo> | NF1_intermediate |
| <https://casereports.bmj.com/content/2013/bcr-2013-200033> | NF1_intermediate |
| <https://drpanossian.com/neurofibromatosis/types-of-neurofibromatosis/neurofibromatosis-type-1/> | NF1_intermediate |
| <https://journal.medizzy.com/neurofibromatosis-type-1-a-family-case-report/> | NF1_intermediate |
| <https://plasticsurgerykey.com/neurofibromatosis-type-1/> | NF1_intermediate |
| <http://www.pcds.org.uk/clinical-guidance/neurofibromatosis-type-1> | NF1_intermediate |
| <http://www.pcds.org.uk/clinical-guidance/neurofibromatosis-type-1> | NF1_intermediate |
| <http://www.pcds.org.uk/clinical-guidance/neurofibromatosis-type-1> | NF1_intermediate |
| <http://www.globalskinatlas.com/upload/762_1.jpg> | NF1_intermediate |
| <https://casereports.bmj.com/content/2013/bcr-2013-200033> | NF1_intermediate |
| <https://casereports.bmj.com/content/2013/bcr-2013-200033> | NF1_intermediate |
| <https://www.researchgate.net/figure/Cafe-au-lait-spots-and-neurofibromas-in-the-umbilical-region_fig1_313776004> | NF1_intermediate |
| <https://www.neurores.org/tables/jnr358et.htm> | NF1_intermediate |
| <https://www.saudijhealthsci.org/viewimage.asp?img=SaudiJHealthSci_2017_6_1_65_210814_f1.jpg> | NF1_intermediate |
| <https://www.saudijhealthsci.org/viewimage.asp?img=SaudiJHealthSci_2017_6_1_65_210814_f1.jpg> | NF1_intermediate |
| <https://www.e-ijd.org/viewimage.asp?img=IndianJDermatol_2010_55_1_105_60366_u3.jpg> | NF1_intermediate |
| <https://www.grepmed.com/images/1994/neurofibromas-dermatology-cafeaulait-skinrash-clinical> | NF1_intermediate |
| <https://www.researchgate.net/figure/Multiple-cafe-au-lait-spots-scattered-over-the-patients-entire-body_fig1_223990602> | NF1_intermediate |
| <https://www.ncbi.nlm.nih.gov/pmc/articles/PMC7402965/> | NF1_intermediate |
| <https://www.sciencedirect.com/science/article/pii/S1474442214700638?via%3Dihub> | NF1_intermediate |
| <https://bmccancer.biomedcentral.com/articles/10.1186/s12885-019-6375-9> | NF1_intermediate |
| <https://www.e-aps.org/journal/view.php?doi=10.5999/aps.2013.40.1.57> | NF1_intermediate |
| <https://onlinelibrary.wiley.com/doi/full/10.1111/j.1346-8138.2005.tb00711.x?sid=nlm%3Apubmed> | NF1_intermediate |
| <https://www.ncbi.nlm.nih.gov/books/NBK557492/> | NF1_intermediate |
| <https://bmcmedgenet.biomedcentral.com/articles/10.1186/s12881-018-0615-8/figures/2> | NF1_intermediate |
| <https://www.ncbi.nlm.nih.gov/pmc/articles/PMC5976335/> | NF1_intermediate |
| <https://www.sciencedirect.com/science/article/pii/S2212440319303931?via%3Dihub> | NF1_intermediate |
| <https://www.sciencedirect.com/science/article/pii/S0190962207005476?via%3Dihub> | NF1_intermediate |
| <https://www.sciencedirect.com/science/article/pii/S0190962207005476?via%3Dihub> | NF1_intermediate |
| <https://www.tandfonline.com/doi/pdf/10.1586/ern.09.4?needAccess=true> | NF1_intermediate |
| <https://www.sciencedirect.com/science/article/pii/S0887899404005557?via%3Dihub> | NF1_intermediate |
| <https://reader.elsevier.com/reader/sd/pii/S1071909198800028?token=0AA8D0CF7587735FA27B958409D7538E0BA083F72D57DF97EA403F0E48E35DB3282ECEB1CDDFD618F46B31CBF76AF0CD> | NF1_intermediate |
| <http://xbyxb.csu.edu.cn/xbwk/fileup/PDF/201807811.pdf> | NF1_intermediate |
| <https://academic.oup.com/qjmed/article/110/9/583/3084728> | NF1_intermediate |
| <https://link.springer.com/article/10.1007/BF03347397> | NF1_intermediate |
| <https://journals.lww.com/co-ophthalmology/Fulltext/2012/09000/Type_I_neurofibromatosis___a.5.aspx> | NF1_intermediate |
| <https://www.geneticsmr.com/sites/default/files/articles/year2016/vol15-2/pdf/gmr7572_1.pdf> | NF1_intermediate |
| <https://www.geneticsmr.com/sites/default/files/articles/year2016/vol15-2/pdf/gmr7572_1.pdf> | NF1_intermediate |
| <https://www.sciencedirect.com/science/article/pii/S0378111920303279?via%3Dihub> | NF1_intermediate |
| <https://www.sciencedirect.com/science/article/pii/S0378111920303279?via%3Dihub> | NF1_intermediate |
| <https://onlinelibrary.wiley.com/doi/full/10.1111/j.1346-8138.2001.tb00090.x?sid=nlm%3Apubmed> | NF1_intermediate |
| <https://www.sciencedirect.com/science/article/pii/S0929693X15002341?via%3Dihub#fig0005> | NF1_intermediate |
| <https://www.sciencedirect.com/science/article/pii/S1553465011001294?via%3Dihub#fig1> | NF1_intermediate |
| <https://www.ncbi.nlm.nih.gov/pmc/articles/PMC5708999/> | NF1_intermediate |
| <https://www.geneticsmr.com/sites/default/files/articles/year2014/vol13-3/pdf/gmr3532.pdf> | NF1_intermediate |
| <https://www.geneticsmr.com/sites/default/files/articles/year2014/vol13-3/pdf/gmr3532.pdf> | NF1_intermediate |
| <https://www.geneticsmr.com/sites/default/files/articles/year2014/vol13-3/pdf/gmr3532.pdf> | NF1_intermediate |
| <https://bmcmedgenet.biomedcentral.com/articles/10.1186/s12881-018-0615-8> | NF1_late |
| <https://www.merckmanuals.com/professional/pediatrics/neurocutaneous-syndromes/neurofibromatosis> | NF1_late |
| <https://www.researchgate.net/figure/Clinical-manifestations-of-patients-with-neurofibromatosis-type-1-in-the-Chinese-family_fig3_304808475> | NF1_late |
| <https://www.sciencedirect.com/science/article/pii/S2212440319303931> | NF1_late |
| <https://bmcmedgenet.biomedcentral.com/articles/10.1186/s12881-018-0615-8> | NF1_late |
| <https://www.researchgate.net/figure/A-70-year-old-woman-with-neurofibromatosis-type-1-Ectoscopic-examination-shows-multiple_fig2_330446967> | NF1_late |
| <http://www.scielo.br/scielo.php?pid=S0034-72802013000200013&script=sci_arttext&tlng=en> | NF1_late |
| <http://ceo.medword.net/?signs=neurofibromatosis-type-1-nf-1-skin> | NF1_late |
| <https://www.scielo.br/scielo.php?script=sci_arttext&pid=S0365-05962013000300329> | NF1_late |
| <https://jmedicalcasereports.biomedcentral.com/articles/10.1186/1752-1947-6-179> | NF1_late |
| <https://jmedicalcasereports.biomedcentral.com/articles/10.1186/1752-1947-6-179> | NF1_late |
| <https://www.researchgate.net/figure/A-view-of-the-back-of-the-patient-Numerous-cafe-au-lait-macules-and-cutaneous_fig1_229552432> | NF1_late |
| <https://www.huidziekten.nl/afbeeldingen/neurofibromatosis-6.jpg> | NF1_late |
| <https://www.researchgate.net/figure/Image-du-pere-de-notre-patient-montrant-de-multiples-neurofibromes-cutanes_fig2_324222416> | NF1_late |
| <https://drpanossian.com/before-and-after/neurofibromatosis-photos-8712/> | NF1_late |
| <https://www.wjgnet.com/1007-9327/full/v24/i33/3806.htm> | NF1_late |
| https://www.wjgnet.com/1007-9327/full/v24/i33/3806.htm | NF1_late |
| <https://www.ncbi.nlm.nih.gov/books/NBK539707/> | NF1_late |
| <https://www.sciencedirect.com/science/article/pii/S1769721217302896?via%3Dihub> | NF1_late |
| <https://www.ncbi.nlm.nih.gov/pmc/articles/PMC6848415/> | NF1_late |
| <https://www.sciencedirect.com/science/article/pii/S109291341300155X?via%3Dihub#f0005> | NF1_late |
| <https://casereports.bmj.com/content/12/11/e230637.long> | NF1_late |
| <https://www.sciencedirect.com/science/article/pii/S1878875018328687?via%3Dihub#fig1> | NF1_late |
| <https://www.e-aps.org/journal/view.php?doi=10.5999/aps.2013.40.1.57> | NF1_late |
| <https://www.annalsafrmed.org/article.asp?issn=1596-3519;year=2020;volume=19;issue=2;spage=150;epage=152;aulast=Yahya> | NF1_late |
| https://www.annalsafrmed.org/article.asp?issn=1596-3519;year=2020;volume=19;issue=2;spage=150;epage=152;aulast=Yahya | NF1_late |
| <https://www.tandfonline.com/doi/pdf/10.3109/17453678609156819> | NF1_late |
| <https://www.semanticscholar.org/paper/Neurofibromatosis-Type-1-(von-Recklinghausen's-in-Lungu-Baican/bf711d669e893775dcef0a135f6f8f34aaa22880> | NF1_late |
| <https://www.researchgate.net/figure/The-patient-has-multiple-cutaneous-neurofibromas-on-the-trunk_fig1_330547353> | NF1_late |
| <https://www.wjgnet.com/1007-9327/full/v24/i4/537.htm> | NF1_late |
| <https://www.dovemed.com/diseases-conditions/neurofibromatosis-type-1/> | NF1_late |
| <https://bmcmedgenet.biomedcentral.com/articles/10.1186/s12881-018-0615-8/figures/2> | NF1_late |
| <https://bmcmedgenet.biomedcentral.com/articles/10.1186/s12881-018-0615-8/figures/2> | NF1_late |
| <http://atlasgeneticsoncology.org/Tumors/NeurofibromaID5098.html> | NF1_late |
| <https://www.ncbi.nlm.nih.gov/books/NBK459358/figure/article-25786.image.f1/> | NF1_late |
| <https://www.nejm.org/doi/full/10.1056/NEJMicm1103859> | NF1_late |
| <https://www.nejm.org/doi/full/10.1056/NEJMicm1103859> | NF1_late |
| <https://eurekalert.org/multimedia/pub/183856.php?from=409748> | NF1_late |
| <https://www.nature.com/articles/s41416-018-0073-2> | NF1_late |
| <https://www.nature.com/articles/s41416-018-0073-2> | NF1_late |
| <https://www.nature.com/articles/s41416-018-0073-2> | NF1_late |
| <https://onlinelibrary.wiley.com/doi/pdf/10.1002/ajmg.a.37045> | NF1_late |
| <https://onlinelibrary.wiley.com/doi/pdf/10.1002/ajmg.a.37045> | NF1_late |
| <https://www.researchgate.net/figure/Clinical-features-of-NF1-probands_fig3_51568778> | NF1_late |
| <https://thasso.com/first-therapy-for-children-with-debilitating-nf-1-approved-in-the-usa/> | NF1_late |
| <https://www.researchgate.net/figure/Multiple-cafe-au-lait-spots-scattered-over-the-patients-entire-body_fig1_223990602> | NF1_late |
| <https://www.researchgate.net/figure/Multiple-skin-neurofibromas-with-freckles-and-cafe-au-lait-spots_fig1_269538723> | NF1_late |
| <https://www.asianjns.org/article.asp?issn=1793-5482;year=2015;volume=10;issue=4;spage=344;epage=347;aulast=Varghese> | NF1_late |
| <https://ejcrp.org/article.asp?issn=2311-3006%3Byear=2020%3Bvolume=7%3Bissue=2%3Bspage=90%3Bepage=94%3Baulast=Chiu%3Btype=3> | NF1_late |
| <https://www.stepwards.com/?page_id=1997> | NF1_late |
| <https://www.jprasurg.com/article/S1748-6815(07)00245-8/fulltext> | NF1_late |
| <https://www.jprasurg.com/article/S1748-6815(07)00245-8/fulltext> | NF1_late |
| <https://www.jprasurg.com/article/S1748-6815(07)00245-8/fulltext> | NF1_late |
| <http://www.globalskinatlas.com/upload/865_1.jpg> | NF1_late |
| <http://www.globalskinatlas.com/upload/865_2.jpg> | NF1_late |
| <https://www.ijdvl.com/article.asp?issn=0378-6323;year=2017;volume=83;issue=2;spage=231;epage=233;aulast=Lin> | NF1_late |
| <http://hjog.org/wp-content/pdf/2018/Kalmantis-L.pdf> | NF1_late |
| <http://studymedicalphotos.blogspot.com/2017/07/a-26-year-old-man-with-lesions-on-his.html> | NF1_late |
| <https://www.pharmazeutische-zeitung.de/krankheit-mit-vielen-gesichtern/> | NF1_late |
| <https://deximed.de/home/b/neurologie/patienteninformationen/erbliche-und-angeborene-erkrankungen/neurofibromatose-typ1/> | NF1_late |
| <https://d-nb.info/1167274407/34> | NF1_late |
| <https://www.ncbi.nlm.nih.gov/pmc/articles/PMC4753888/> | NF1_late |
| <https://www.ncbi.nlm.nih.gov/pmc/articles/PMC7584462/> | NF1_late |
| <https://www.ncbi.nlm.nih.gov/pmc/articles/PMC5026286/> | NF1_late |
| <https://www.ecronicon.com/ecop/pdf/ECOP-10-00433.pdf> | NF1_late |
| <https://link.springer.com/referenceworkentry/10.1007%2F978-1-4939-2401-1_178> | NF1_late |
| <https://cdn.mdedge.com/files/s3fs-public/Document/September-2017/6108JFP_PhotoRounds.pdf> | NF1_late |
| <https://journals.lww.com/amjforensicmedicine/Fulltext/2016/09000/Can_Severe_Kyphoscoliosis_Lead_to_Aorta_Rupture_.17.aspx> | NF1_late |
| <https://www.ncbi.nlm.nih.gov/pmc/articles/PMC6744821/> | NF1_late |
| <https://www.mdpi.com/2296-3529/8/1/3> | NF1_late |
| <https://www.sciencedirect.com/science/article/pii/S1748681516300146?via%3Dihub#fig4> | NF1_late |
| <https://repository.kulib.kyoto-u.ac.jp/dspace/bitstream/2433/153012/1/58_17.pdf> | NF1_late |
| <https://www.sciencedirect.com/science/article/pii/S2210261212002258?via%3Dihub#fig0015> | NF1_late |
| <https://link.springer.com/referenceworkentry/10.1007%2F978-1-4939-2401-1_178> | NF1_late |
| <http://www.brainkart.com/article/Neurofibromatosis_26641/> | NF1_late |
| <https://neupsykey.com/neurocutaneous-syndromes-3/> | NF1_late |
| <https://jmedicalcasereports.biomedcentral.com/articles/10.1186/s13256-019-2292-4> | NF1_late |
| <https://jmedicalcasereports.biomedcentral.com/articles/10.1186/s13256-019-2292-4> | NF1_late |
| <https://academic.oup.com/neurosurgery/article/31/CN_suppl_1/417/4099502> | NF1_late |
| <https://www.thieme-connect.com/products/ejournals/pdf/10.1055/s-2008-1035910.pdf> | NF1_late |
| <https://www.ncbi.nlm.nih.gov/pmc/articles/PMC4907302/> | NF1_late |
| <https://ykhoa.org/d/image.htm?imageKey=PEDS%2F69606%7EDERM%2F82613%7EDERM%2F67532> | NF1_late |
| <https://www.thieme-connect.com/products/ejournals/html/10.1055/s-0033-1335433> | NF1_late |
| <https://www.thieme-connect.com/products/ejournals/html/10.1055/s-0033-1335433> | NF1_late |
| <https://content.sciendo.com/view/journals/bjmg/21/2/article-p45.xml?product=sciendo> | NF1_late |
| <https://content.sciendo.com/view/journals/bjmg/21/2/article-p45.xml?product=sciendo> | NF1_late |
| <https://casereports.bmj.com/content/2013/bcr-2013-200033> | NF1_late |
| <https://jmedicalcasereports.biomedcentral.com/articles/10.1186/s13256-015-0533-8> | NF1_Late |
| <https://www.dovepress.com/the-investigation-for-potential-modifier-genes-in-patients-with-neurof-peer-reviewed-fulltext-article-OTT> | NF1_late |
| <https://www.almeka.in/skin_neurofibroma/> | NF1_late |
| <http://www.pcds.org.uk/clinical-guidance/neurofibromatosis-type-1> | NF1_late |
| <https://www.ojoonline.org/viewimage.asp?img=OmanJOphthalmol_2015_8_3_208_169894_f2.jpg> | NF1_late |
| <http://www.clinicaterapeutica.it/2020/171/5/02_MIRAGLIA.pdf> | NF1_late |
| <http://www.clinicaterapeutica.it/2020/171/5/02_MIRAGLIA.pdf> | NF1_late |
| <http://www.clinicaterapeutica.it/2020/171/5/02_MIRAGLIA.pdf> | NF1_late |
| <https://www.sciencedirect.com/science/article/pii/S1878875020305465?via%3Dihub#fig2> | NF1_late |
| <https://link.springer.com/article/10.1007%2Fs12031-018-1128-9> | NF1_late |
| <https://www.ncbi.nlm.nih.gov/pmc/articles/PMC6925767/> | NF1_late |
| <https://journals.lww.com/jaaos/Fulltext/2010/06000/Orthopaedic_Manifestations_of_Neurofibromatosis.7.aspx> | NF1_late |
| <https://www.nature.com/articles/gim20101> | NF1_late |
| <https://www.nature.com/articles/gim20101> | NF1_late |
| <https://reader.elsevier.com/reader/sd/pii/S0248866304002437?token=D35F73468FF63B917EFB31D27F3694FE5D6E6E995C477C5B604CE9D5A1987F916B2243F2F10A5F287B50913FB78BA1D2> | NF1_late |
| <https://www.geneticsmr.com/sites/default/files/articles/year2014/vol13-3/pdf/gmr3532.pdf> | NF1_late |
