## Supplemental material (survey) for "Proof-of-principle neural network models for classification, attribution, creation, style-mixing, and morphing of image data for genetic conditions"

### Directions and Demographic Questions

Thank you for participating in this study – we are truly grateful. This project involves using "artificial intelligence" to see whether we can help build useful tools that may be relevant for different genetic conditions.

We will first ask a few general questions about your background (e.g., what kind of clinician you are and where you practice). We will then ask you a series of questions asking you to "classify" or diagnose some data related to genetic conditions. For example, we may show you images and ask you to choose which condition the images represent. The images you see will be randomly selected from a larger dataset. We anticipate that participation will take most people less than 30 minutes.

Some of the items may be difficult to identify, while others may be easier. For example, this may be because some of images are cropped or magnified in certain ways. This can help us understand which types of images are easier or harder to classify.

You may take this survey on your smart device or your desktop/laptop computer. You are encouraged to adjust your zoom to best view the image and select your answer. Please do not use the internet or textbooks. We are interested in your instinctive response to each question. Once you answer a question, you will not be able to go back.

No identifiers will be collected as part of the survey. All individual results will be maintained in an anonymous format. You will not receive your individual results. We plan to describe/publish only deidentified results.

Your participation is voluntary and will not be compensated. We have been granted exemption from IRB review by the National Institutes of Health (NIH). By continuing with the survey, you agree to take part.

If you have questions, encounter any difficulties or change your mind about participation, please contact Rebekah Waikel at or 301.435.6558.

What is your area of specialty?

- Family Medicine
- Medical Genetics
- Pediatrics

For how long (since your last residency or fellowship) have you been practicing?

- < 1 year
- 1 to 5 years
- 5 to 10 years
- > 10 years

Where do you currently practice medicine?

- Africa
- Asia
- Australia/Oceania
- Europe
- North America
- South America

### Skin Manifestation Questions

For the following questions you will view photographs of skin lesions. Below each image you will select the condition that best describes the skin lesion(s). Possible conditions include Hypomelanosis of Ito, Incontinentia Pigmenti, McCune-Albright Syndrome, Noonan Syndrome with Multiple Lentigines, Neurofibromatosis Type 1, and Tuberous Sclerosis Complex. For skin lesions that cannot be classified in the listed 6 conditions, select the last box indicating that the lesion is not one of the listed conditions. You are encouraged to adjust your zoom (in or out) to best view the image and select your answer. After you have selected your answer, click the arrow at the bottom right of the window to enter your answer and progress to the next question.

### Set 1

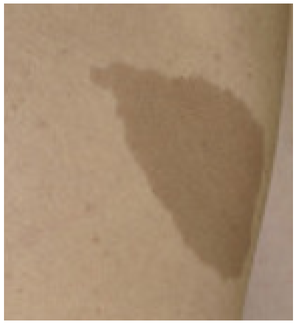

| Hypomelanosis of Ito | Incontinentia Pigmenti | McCune-Albright Syndrome | Noonan with Multiple Lentigines | Neurofibromatosis Type 1 | Tuberous Sclerosis Complex | Not one of the listed conditions |
|---|---|---|---|---|---|---|
| ○ | ○ | ○ | ○ | ○ | ○ | ○ |

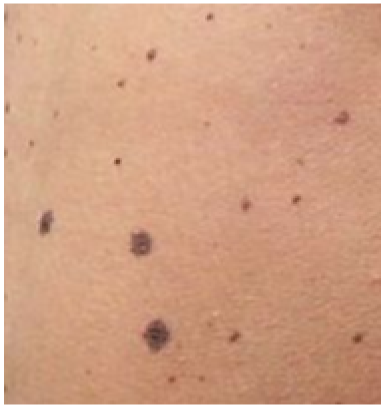

| Hypomelanosis of Ito | Incontinentia Pigmenti | McCune-Albright Syndrome | Noonan with Multiple Lentigines | Neurofibromatosis Type 1 | Tuberous Sclerosis Complex | Not one of the listed conditions |
|---|---|---|---|---|---|---|
| ○ | ○ | ○ | ○ | ○ | ○ | ○ |

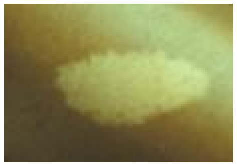

| Hypomelanosis of Ito | Incontinentia Pigmenti | McCune-Albright Syndrome | Noonan with Multiple Lentigines | Neurofibromatosis Type 1 | Tuberous Sclerosis Complex | Not one of the listed conditions |
|---|---|---|---|---|---|---|
| ○ | ○ | ○ | ○ | ○ | ○ | ○ |

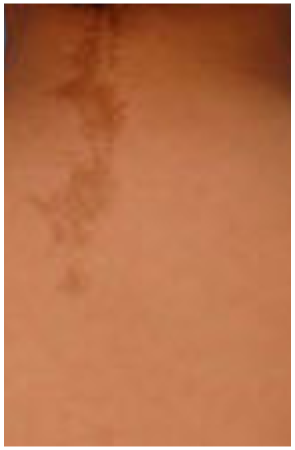

| Hypomelanosis of Ito | Incontinentia Pigmenti | McCune-Albright Syndrome | Noonan with Multiple Lentigines | Neurofibromatosis Type 1 | Tuberous Sclerosis Complex | Not one of the listed conditions |
|---|---|---|---|---|---|---|
| ○ | ○ | ○ | ○ | ○ | ○ | ○ |

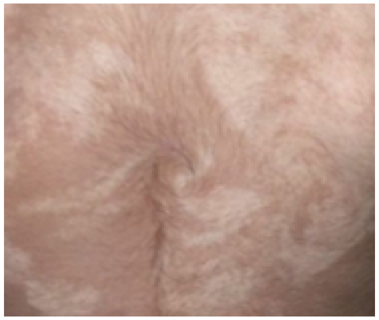

| Hypomelanosis of Ito | Incontinentia Pigmenti | McCune-Albright Syndrome | Noonan with Multiple Lentigines | Neurofibromatosis Type 1 | Tuberous Sclerosis Complex | Not one of the listed conditions |
|---|---|---|---|---|---|---|
| ○ | ○ | ○ | ○ | ○ | ○ | ○ |

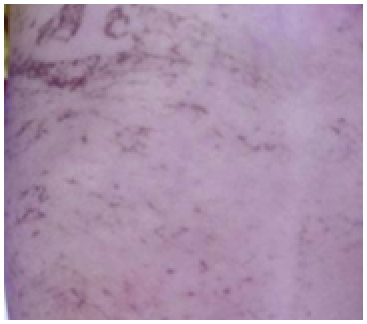

| Hypomelanosis of Ito | Incontinentia Pigmenti | McCune-Albright Syndrome | Noonan with Multiple Lentigines | Neurofibromatosis Type 1 | Tuberous Sclerosis Complex | Not one of the listed conditions |
|---|---|---|---|---|---|---|
| ○ | ○ | ○ | ○ | ○ | ○ | ○ |

### Set 2

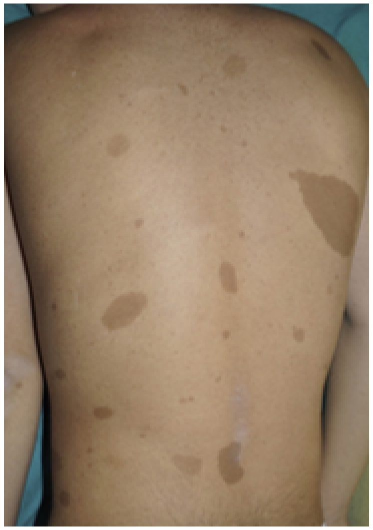

| Hypomelanosis of Ito | Incontinentia Pigmenti | McCune-Albright Syndrome | Noonan with Multiple Lentigines | Neurofibromatosis Type 1 | Tuberous Sclerosis Complex | Not one of the listed conditions |
|---|---|---|---|---|---|---|
| ○ | ○ | ○ | ○ | ○ | ○ | ○ |

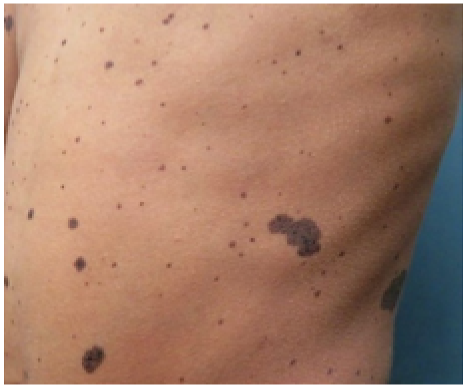

| Hypomelanosis of Ito | Incontinentia Pigmenti | McCune-Albright Syndrome | Noonan with Multiple Lentigines | Neurofibromatosis Type 1 | Tuberous Sclerosis Complex | Not one of the listed conditions |
|---|---|---|---|---|---|---|
| ○ | ○ | ○ | ○ | ○ | ○ | ○ |

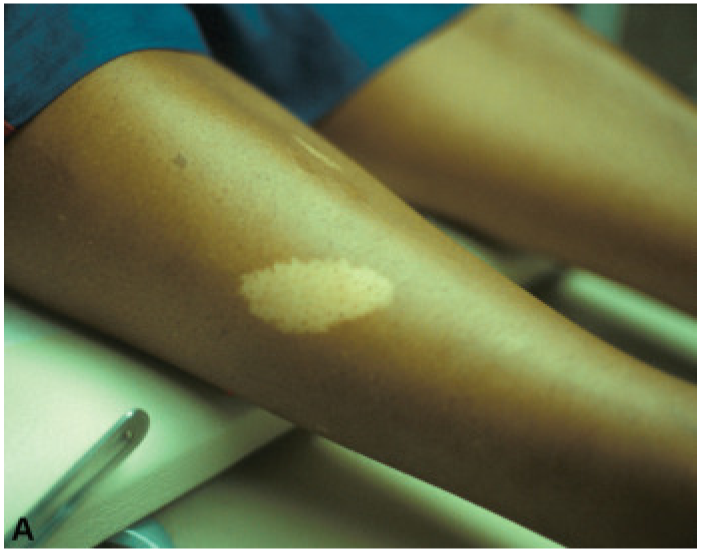


| Hypomelanosis of Ito | Incontinentia Pigmenti | McCune-Albright Syndrome | Noonan with Multiple Lentigines | Neurofibromatosis Type 1 | Tuberous Sclerosis Complex | Not one of the listed conditions |
|---|---|---|---|---|---|---|
| ○ | ○ | ○ | ○ | ○ | ○ | ○ |

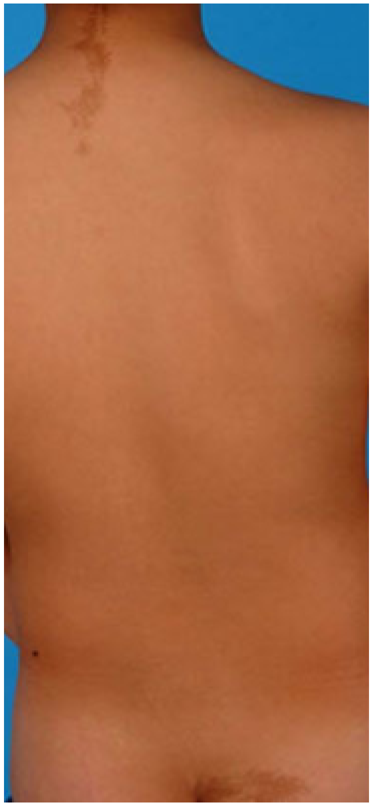

| Hypomelanosis of Ito | Incontinentia Pigmenti | McCune-Albright Syndrome | Noonan with Multiple Lentigines | Neurofibromatosis Type 1 | Tuberous Sclerosis Complex | Not one of the listed conditions |
|---|---|---|---|---|---|---|
| ○ | ○ | ○ | ○ | ○ | ○ | ○ |

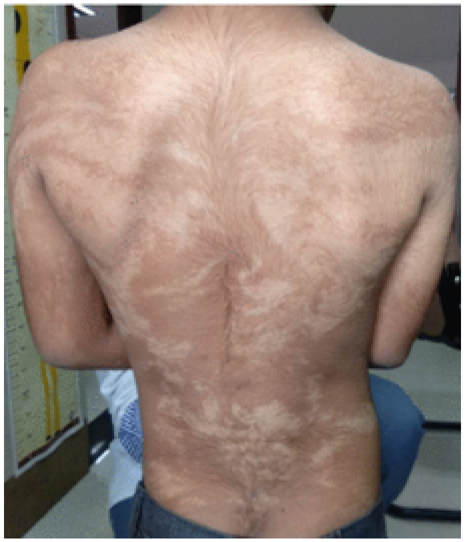

| Hypomelanosis of Ito | Incontinentia Pigmenti | McCune-Albright Syndrome | Noonan with Multiple Lentigines | Neurofibromatosis Type 1 | Tuberous Sclerosis Complex | Not one of the listed conditions |
|---|---|---|---|---|---|---|
| ○ | ○ | ○ | ○ | ○ | ○ | ○ |

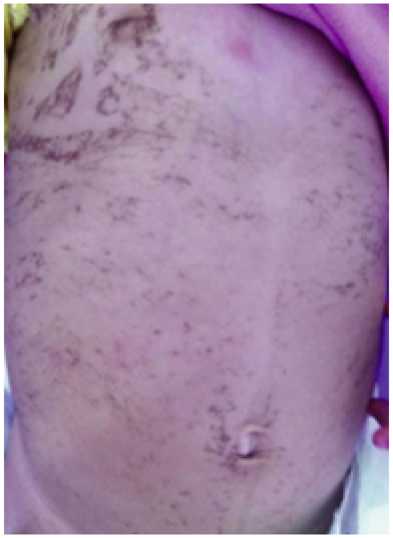

| Hypomelanosis of Ito | Incontinentia Pigmenti | McCune-Albright Syndrome | Noonan with Multiple Lentigines | Neurofibromatosis Type 1 | Tuberous Sclerosis Complex | Not one of the listed conditions |
|---|---|---|---|---|---|---|

○ ○ ○ ○ ○ ○ ○

### Thank you

End of survey. Thank you.

Powered by Qualtrics
