## Supplementary material for "Proof-of-principle neural network models for classification, attribution, creation, style-mixing, and morphing of image data for genetic conditions": Image permission

BMJ PUBLISHING GROUP LTD. LICENSE
TERMS AND CONDITIONS

Mar 28, 2021

---

This Agreement between Benjamin Solomon ("You") and BMJ Publishing Group Ltd. ("BMJ Publishing Group Ltd.") consists of your license details and the terms and conditions provided by BMJ Publishing Group Ltd. and Copyright Clearance Center.

| | |
|---|---|
| License Number | 5037700979686 |
| License date | Mar 28, 2021 |
| Licensed Content Publisher | BMJ Publishing Group Ltd. |
| Licensed Content Publication | BMJ Case Reports |
| Licensed Content Title | Hypomelanosis of Ito: streaks and whorls |
| Licensed Content Author | Daisy Khera,Saurabh Singh,Priya Gupta |
| Licensed Content Date | Apr 1, 2019 |
| Licensed Content Volume | 12 |
| Licensed Content Issue | 4 |
| Type of Use | Journal/Magazine |
| Requestor type | Government/Public Sector |
| Format | Print and electronic |

| | |
|---|---|
| Portion | Figure/table/extract |
| Number of figure/table/extracts | 1 |
| Descriptionof figure/table/extracts | Figure 1 |
| Will you be translating? | No |
| Circulation/distribution | 100000 |
| Order reference number | 12 |
| Title of new article | A proof-of-principle neural network model for classification, attribution, creation, and morphing of image data for genetic conditions |
| Lead author | Benjamin Solomon |
| Title of targeted journal | medRxiv |
| Publisher | Cold Spring Harbor Laboratory |
| Expected publication date | Apr 2021 |
| Order reference number | 12 |
| Portions | Figure 1 |
| Requestor Location | Benjamin Solomon<br>Building 35, Room 1B-207<br>National Institutes of Health<br>MSC 3717<br>Bethesda, MD 20892 |

| | |
|---|---|
| | United States<br>Attn: Benjamin Solomon |
| Publisher Tax ID | GB674738491 |
| Total | 0.00 USD |

Terms and Conditions

**BMJ Group Terms and Conditions for Permissions**

When you submit your orderyou are subject to the terms and conditions set out below. You will also haveagreed to the Copyright Clearance Center's ("CCC") terms and conditionsregarding billing and payment https://s100.copyright.com/App/PaymentTermsAndConditions.jsp. CCC are acting as the BMJ Publishing Group Limited's ("BMJ Group's") agent.

Subject to the terms set outherein, the BMJ Group hereby grants to you (the Licensee) a non-exclusive,non-transferable licence to re-use material as detailed in your request forthis/those purpose(s) only and in accordance with the following conditions:

1) **Scope of Licence:** Useof the Licensed Material(s) is restricted to the ways specified by you duringthe order process and any additional use(s) outside of those specified in thatrequest, require a further grant of permission.

2) **Acknowledgement:** Inall cases, due acknowledgement to the original publication with permission fromthe BMJ Group should be stated adjacent to the reproduced Licensed Material.The format of such acknowledgement should read as follows:

"Reproduced from [publicationtitle, author(s), volume number, page numbers, copyright notice year] withpermission from BMJ Publishing Group Ltd."

3) **Third Party Material**:BMJ Group acknowledges to the best of its knowledge, it has the rights to licenceyour reuse of the Licensed Material, subject always to the caveat thatimages/diagrams, tables and other illustrative material included within, whichhave a separate copyright notice, are presumed as excluded from the licence. Therefore,you should ensure that the Licensed Material you are requesting is original toBMJ Group and does not carry the copyright of another entity (as credited inthe published version). If the credit line on any part of the material you haverequested in any way indicates that it was reprinted or adapted by BMJ Groupwith permission from another source, then you should seek permission from thatsource directly to re-use the Licensed Material, as this is outside of thelicence granted herein.

4) **Altering/ModifyingMaterial:** The text of any material for which a licence is granted may notbe altered in any way without the prior express permission of the BMJ Group. Subject to Clause 3 above however, single figure adaptations do not require BMJGroup's approval; however, the adaptation should be credited as follows:

"Adapted by permission fromBMJ Publishing Group Limited. [publication title, author, volume number, pagenumbers, copyright notice year]

5) **Reservation of Rights:**The BMJ Group reserves all rights not specifically granted in the combinationof (i) the licence details provided by you and accepted in the course of thislicensing transaction, (ii) these terms and conditions and (iii) CCC's Billing and Payment Terms and Conditions.

6) **Timing of Use:** Firstuse of the Licensed Material must take place within 12 months of the grant ofpermission.

7). **Creation of Contractand Termination:** Once you have submitted an order via Rightslink and thisis received by CCC, and subject to you completing accurate details of yourproposed use, this is when a binding contract is in effect and our acceptanceoccurs. As you are ordering rights from a periodical, to the fullest extentpermitted by law, you will have no right to cancel the contract from this pointother than for BMJ Group's material breach or fraudulent misrepresentation oras otherwise permitted under a statutory right. Payment must be made inaccordance with CCC's Billing and Payment Terms and conditions. In the eventthat you breach any material condition of these terms and condition or any of CCC's Billing and Payment Terms and Conditions, the license is automatically terminated uponwritten notice from the BMJ Group or CCC or as otherwise provided for in CCC's Billing and Payment Terms and Conditions, where these apply.. Continued use of materials wherea licence has been terminated, as well as any use of the Licensed Materialsbeyond the scope of an unrevoked licence, may constitute intellectual propertyrights infringement and BMJ Group reserves the right to take any and all actionto protect its intellectual property rights in the Licensed Materials.

8. **Warranties:** BMJGroup makes no express or implied representations or warranties with respect tothe Licensed Material and to the fullest extent permitted by law this isprovided on an "as is" basis. For the avoidance of doubt BMJ Group does notwarrant that the Licensed Material is accurate or fit for any particularpurpose.

9. **Limitation ofLiability:** To the fullest extent permitted by law, the BMJ Group disclaimsall liability for any indirect, consequential or incidental damages (includingwithout limitation, damages for loss of profits, information or interruption)arising out of the use or inability to use the Licensed Material or theinability to obtain additional rights to use the Licensed Material. To thefullest extent permitted by law, the maximum aggregate liability of the BMJGroup for any claims, costs, proceedings and demands for direct losses causedby BMJ Group's breaches of its obligations herein shall be limited to twice theamount paid by you to CCC for the licence granted herein.

10. **Indemnity:** You hereby indemnify and holdharmless the BMJ Group and their respective officers, directors, employees andagents, from and against any and all claims, costs, proceeding or demands arisingout of your unauthorised use of the Licensed Material.

11. **No Transfer of License:** This licence ispersonal to you, and may not be assigned or transferred by you without priorwritten consent from the BMJ Group or its authorised agent(s). BMJ Groupmay assign or transfer any of its rights and obligations under this Agreement,upon written notice to you.

12. **No Amendment Except inWriting:** This licence may not be amended except in a writing signed by bothparties (or, in the case of BMJ Group, by CCC on the BMJ Group's behalf).

13. **Objection to Contraryterms:** BMJ Group hereby objects to any terms contained in any purchaseorder, acknowledgment, check endorsement or other writing prepared by you,which terms are inconsistent with these terms and conditions or CCC's Billing and Payment Terms and Conditions. These terms and conditions, together with CCC's Billing and Payment Terms and Conditions (which to the extent they are consistent areincorporated herein), comprise the entire agreement between you and BMJ Group (and CCC) and the Licensee concerning this licensing transaction. In the event ofany conflict between your obligations established by these terms and conditionsand those established by CCC's Billing and Payment Terms and Conditions, theseterms and conditions shall control.

14. **Revocation:** BMJGroup or CCC may, within 30 days of issuance of this licence, deny thepermissions described in this licence at their sole discretion, for any reasonor no reason, with a full refund payable to you should you have not been ableto exercise your rights in full. Notice of such denial will be made using thecontact information provided by you. Failure to receive such notice from BMJGroup or CCC will not, to the fullest extent permitted by law alter orinvalidate the denial. For the fullest extent permitted by law in no event willBMJ Group or CCC be responsible or liable for any costs, expenses or damageincurred by you as a result of a denial of your permission

request, other thana refund of the amount(s) paid by you to BMJ Group and/or CCC for denied permissions.

15. **Restrictions to thelicense**:

**15.1 Promotion:** BMJ Group will not give permission to reproduce infull or in part any Licensed Material for use in the promotion of thefollowing:

a) non-medical products thatare harmful or potentially harmful to health: alcohol, baby milks and/or, sunbeds

b) medical products that donot have a product license granted by the Medicines and Healthcare productsRegulatory Agency (MHRA) or its international equivalents. Marketing of theproduct may start only after data sheets have been released to members of themedical profession and must conform to the marketing authorization contained inthe product license.

**16. Translation**: This permission is granted for non-exclusive worldEnglish language rights only unless explicitly stated in your licence. Iftranslation rights are granted, a professional translator should be employedand the content should be reproduced word for word preserving the integrity ofthe content.

17. **General:**Neither party shall be liable for failure, default or delay in performingits obligations under this Licence, caused by a Force Majeure event which shallinclude any act of God, war, or threatened war, act or threatened act ofterrorism, riot, strike, lockout, individual action, fire, flood, drought,tempest or other event beyond the reasonable control of either party.

17.1 Inthe event that any provision of this Agreement is held to be invalid, theremainder of the provisions shall continue in full force and effect.

17.2 Thereshall be no right whatsoever for any third party to enforce the terms andconditions of this Agreement. The Parties hereby expressly wish to exclude theoperation of the Contracts (Rights of Third Parties) Act 1999 and any otherlegislation which has this effect and is binding on this agreement.

17.3 Tothe fullest extent permitted by law, this Licence will be governed by the lawsof England and shall be governed and construed in accordance with the laws of England. Any action arising out of or relating to this agreement shall be brought in courtssituated in England save where it is necessary for BMJ Group for enforcement tobring proceedings to bring an action in an alternative jurisdiction.

**Questions? or +1-855-239-3415 (toll free in the US) or +1-978-646-2777.**
