## Supplementary material for "Proof-of-principle neural network models for classification, attribution, creation, style-mixing, and morphing of image data for genetic conditions": Image permission

This page is available in the following languages:

### Creative Commons License Deed

**Attribution-NonCommercial 3.0 Unported (CC BY-NC 3.0)**

This is a human-readable summary of (and not a substitute for) the license.

#### You are free to:

**Share** — copy and redistribute the material in any medium or format

**Adapt** — remix, transform, and build upon the material

The licensor cannot revoke these freedoms as long as you follow the license terms.

#### Under the following terms:

**Attribution** — You must give appropriate credit, provide a link to the license, and indicate if changes were made. You may do so in any reasonable manner, but not in any way that suggests the licensor endorses you or your use.

**NonCommercial** — You may not use the material for commercial purposes.

**No additional restrictions** — You may not apply legal terms or technological measures that legally restrict others from doing anything the license permits.

#### Notices:

You do not have to comply with the license for elements of the material in the public domain or where your use is permitted by an applicable exception or limitation.

No warranties are given. The license may not give you all of the permissions necessary for your intended use. For example, other rights such as publicity, privacy, or moral rights may limit how you use the material.
