## Supplementary material for "Proof-of-principle neural network models for classification, attribution, creation, style-mixing, and morphing of image data for genetic conditions": Image permission

ELSEVIER LICENSE
TERMS AND CONDITIONS

Mar 28, 2021

---

This Agreement between Benjamin Solomon ("You") and Elsevier ("Elsevier") consists of your license details and the terms and conditions provided by Elsevier and Copyright Clearance Center.

**All payments must be made in full to CCC. For payment instructions, please see information listed at the bottom of this form.**

| | |
|---|---|
| License Number | 5037701361065 |
| License date | Mar 28, 2021 |
| Licensed Content Publisher | Elsevier |
| Licensed Content Publication | Actas Dermo-Sifiliográficas (English Edition) |
| Licensed Content Title | An Update on Neurofibromatosis Type 1: Not Just Café-au-Lait Spots, Freckling, and Neurofibromas. An Update. Part I. Dermatological Clinical Criteria Diagnostic of the Disease |
| Licensed Content Author | A. Hernández-Martín,A. Duat-Rodríguez |
| Licensed Content Date | July–August 2016 |
| Licensed Content Volume | 107 |
| Licensed Content Issue | 6 |
| Licensed Content Pages | 11 |

| | |
|---|---|
| Start Page | 454 |
| End Page | 464 |
| Type of Use | reuse in a journal/magazine |
| Requestor type | academic/educational institute |
| Portion | figures/tables/illustrations |
| Number of figures/tables/illustrations | 1 |
| Format | both print and electronic |
| Are you the author of this Elsevier article? | No |
| Will you be translating? | No |
| Title of new article | A proof-of-principle neural network model for classification, attribution, creation, and morphing of image data for genetic conditions |
| Lead author | Benjamin Solomon |
| Title of targeted journal | medRxiv |
| Publisher | Cold Spring Harbor Laboratory |
| Expected publication date | Apr 2021 |
| Order reference number | 15 |
| Portions | Figure 1 |
| Requestor Location | Benjamin Solomon<br>Building 35, Room 1B-207 |

National Institutes of Health
MSC 3717
Bethesda, MD 20892
United States
Attn: Benjamin Solomon

| | |
|---|---|
| Publisher Tax ID | 98-0397604 |
| Billing Type | Invoice |
| Billing Address | Benjamin Solomon<br>Building 10, Room 3-2551<br>National Institutes of Health<br><br>BETHESDA, MD 20892<br>United States<br>Attn: Benjamin Solomon |
| Total | 48.30 USD |

Terms and Conditions

**INTRODUCTION**

1. The publisher for this copyrighted material is Elsevier. By clicking "accept" in connection with completing this licensing transaction, you agree that the following terms and conditions apply to this transaction (along with the Billing and Payment terms and conditions established by Copyright Clearance Center, Inc. ("CCC"), at the time that you opened your Rightslink account and that are available at any time at http://myaccount.copyright.com).

**GENERAL TERMS**

2. Elsevier hereby grants you permission to reproduce the aforementioned material subject to the terms and conditions indicated.

3. Acknowledgement: If any part of the material to be used (for example, figures) has appeared in our publication with credit or acknowledgement to another source, permission must also be sought from that source. If such permission is not obtained then that material may not be included in your publication/copies. Suitable acknowledgement to the source must be made, either as a footnote or in a reference list at the end of your publication, as follows:

"Reprinted from Publication title, Vol /edition number, Author(s), Title of article / title of chapter, Pages No., Copyright (Year), with permission from Elsevier [OR APPLICABLE SOCIETY COPYRIGHT OWNER]." Also Lancet special credit - "Reprinted from The Lancet, Vol. number, Author(s), Title of article, Pages No., Copyright (Year), with permission from Elsevier."

4. Reproduction of this material is confined to the purpose and/or media for which permission is hereby given.

5. Altering/Modifying Material: Not Permitted. However figures and illustrations may be altered/adapted minimally to serve your work. Any other abbreviations, additions, deletions and/or any other alterations shall be made only with prior written authorization of Elsevier Ltd. (Please contact Elsevier's permissions helpdesk here). No modifications can be made to any Lancet figures/tables and they must be reproduced in full.

6. If the permission fee for the requested use of our material is waived in this instance, please be advised that your future requests for Elsevier materials may attract a fee.

7. Reservation of Rights: Publisher reserves all rights not specifically granted in the combination of (i) the license details provided by you and accepted in the course of this licensing transaction, (ii) these terms and conditions and (iii) CCC's Billing and Payment terms and conditions.

8. License Contingent Upon Payment: While you may exercise the rights licensed immediately upon issuance of the license at the end of the licensing process for the transaction, provided that you have disclosed complete and accurate details of your proposed use, no license is finally effective unless and until full payment is received from you (either by publisher or by CCC) as provided in CCC's Billing and Payment terms and conditions. If full payment is not received on a timely basis, then any license preliminarily granted shall be deemed automatically revoked and shall be void as if never granted. Further, in the event that you breach any of these terms and conditions or any of CCC's Billing and Payment terms and conditions, the license is automatically revoked and shall be void as if never granted. Use of materials as described in a revoked license, as well as any use of the materials beyond the scope of an unrevoked license, may constitute copyright infringement and publisher reserves the right to take any and all action to protect its copyright in the materials.

9. Warranties: Publisher makes no representations or warranties with respect to the licensed material.

10. Indemnity: You hereby indemnify and agree to hold harmless publisher and CCC, and their respective officers, directors, employees and agents, from and against any and all claims arising out of your use of the licensed material other than as specifically authorized pursuant to this license.

11. No Transfer of License: This license is personal to you and may not be sublicensed, assigned, or transferred by you to any other person without publisher's written permission.

12. No Amendment Except in Writing: This license may not be amended except in a writing signed by both parties (or, in the case of publisher, by CCC on publisher's behalf).

13. Objection to Contrary Terms: Publisher hereby objects to any terms contained in any purchase order, acknowledgment, check endorsement or other writing prepared by you, which terms are inconsistent with these terms and conditions or CCC's Billing and Payment terms and conditions. These terms and conditions, together with CCC's Billing and Payment terms and conditions (which are incorporated herein), comprise the entire agreement between you and publisher (and CCC) concerning this licensing transaction. In the event of any conflict between your obligations established by these terms and conditions and those established by CCC's Billing and Payment terms and conditions, these terms and conditions shall control.

14. Revocation: Elsevier or Copyright Clearance Center may deny the permissions described in this License at their sole discretion, for any reason or no reason, with a full refund payable to you. Notice of such denial will be made using the contact information provided by you. Failure to receive such notice will not alter or invalidate the denial. In no event will Elsevier or Copyright Clearance Center be responsible or liable for any costs, expenses or damage incurred by you as a result of a denial of your permission request, other than a refund of the amount(s) paid by you to Elsevier and/or Copyright Clearance Center for denied permissions.

**LIMITED LICENSE**

The following terms and conditions apply only to specific license types:

15. **Translation**: This permission is granted for non-exclusive world **English** rights only unless your license was granted for translation rights. If you licensed translation rights you may only translate this content into the languages you requested. A professional translator must perform all translations and reproduce the content word for word preserving the integrity of the article.

16. **Posting licensed content on any Website**: The following terms and conditions apply as follows: Licensing material from an Elsevier journal: All content posted to the web site must maintain the copyright information line on the bottom of each image; A hyper-text must be included to the Homepage of the journal from which you are licensing at http://www.sciencedirect.com/science/journal/xxxxx or the Elsevier homepage for books at http://www.elsevier.com; Central Storage: This license does not include permission for a scanned version of the material to be stored in a central repository such as that provided by Heron/XanEdu.

Licensing material from an Elsevier book: A hyper-text link must be included to the Elsevier homepage at http://www.elsevier.com . All content posted to the web site must maintain the copyright information line on the bottom of each image.

**Posting licensed content on Electronic reserve**: In addition to the above the following clauses are applicable: The web site must be password-protected and made available only to bona fide students registered on a relevant course. This permission is granted for 1 year only. You may obtain a new license for future website posting.

17. **For journal authors:** the following clauses are applicable in addition to the above:

**Preprints:**

A preprint is an author's own write-up of research results and analysis, it has not been peer-reviewed, nor has it had any other value added to it by a publisher (such as formatting, copyright, technical enhancement etc.).

Authors can share their preprints anywhere at any time. Preprints should not be added to or enhanced in any way in order to appear more like, or to substitute for, the final versions of articles however authors can update their preprints on arXiv or RePEc with their Accepted Author Manuscript (see below).

If accepted for publication, we encourage authors to link from the preprint to their formal publication via its DOI. Millions of researchers have access to the formal publications on ScienceDirect, and so links will help users to find, access, cite and use the best available version. Please note that Cell Press, The Lancet and some society-owned have different preprint policies. Information on these policies is available on the journal homepage.

**Accepted Author Manuscripts:** An accepted author manuscript is the manuscript of an article that has been accepted for publication and which typically includes author-incorporated changes suggested during submission, peer review and editor-author communications.

Authors can share their accepted author manuscript:

- immediately
  - via their non-commercial person homepage or blog
  - by updating a preprint in arXiv or RePEc with the accepted manuscript
  - via their research institute or institutional repository for internal institutional uses or as part of an invitation-only research collaboration work-group
  - directly by providing copies to their students or to research collaborators for their personal use
  - for private scholarly sharing as part of an invitation-only work group on commercial sites with which Elsevier has an agreement
- After the embargo period
  - via non-commercial hosting platforms such as their institutional repository
  - via commercial sites with which Elsevier has an agreement

In all cases accepted manuscripts should:

- link to the formal publication via its DOI
- bear a CC-BY-NC-ND license - this is easy to do
- if aggregated with other manuscripts, for example in a repository or other site, be shared in alignment with our hosting policy not be added to or enhanced in any way to appear more like, or to substitute for, the published journal article.

**Published journal article (JPA):** A published journal article (PJA) is the definitive final record of published research that appears or will appear in the journal and embodies all value-adding publishing activities including peer review co-ordination, copy-editing, formatting, (if relevant) pagination and online enrichment.

Policies for sharing publishing journal articles differ for subscription and gold open access articles:

**Subscription Articles:** If you are an author, please share a link to your article rather than the full-text. Millions of researchers have access to the formal publications on ScienceDirect, and so links will help your users to find, access, cite, and use the best available version.

Theses and dissertations which contain embedded PJAs as part of the formal submission can be posted publicly by the awarding institution with DOI links back to the formal publications on ScienceDirect.

If you are affiliated with a library that subscribes to ScienceDirect you have additional private sharing rights for others' research accessed under that agreement. This includes use for classroom teaching and internal training at the institution (including use in course packs and courseware programs), and inclusion of the article for grant funding purposes.

**Gold Open Access Articles:** May be shared according to the author-selected end-user license and should contain a CrossMark logo, the end user license, and a DOI link to the formal publication on ScienceDirect.

Please refer to Elsevier's posting policy for further information.

18. **For book authors** the following clauses are applicable in addition to the above: Authors are permitted to place a brief summary of their work online only. You are not allowed to download and post the published electronic version of your chapter, nor may you scan the printed edition to create an electronic version. **Posting to a repository:** Authors are permitted to post a summary of their chapter only in their institution's repository.

19. **Thesis/Dissertation**: If your license is for use in a thesis/dissertation your thesis may be submitted to your institution in either print or electronic form. Should your thesis be published commercially, please reapply for permission. These requirements include permission for the Library and Archives of Canada to supply single copies, on demand, of the complete thesis and include permission for Proquest/UMI to supply single copies, on demand, of the complete thesis. Should your thesis be published commercially, please reapply for permission. Theses and dissertations which contain embedded PJAs as part of the formal submission can be posted publicly by the awarding institution with DOI links back to the formal publications on ScienceDirect.

### Elsevier Open Access Terms and Conditions

You can publish open access with Elsevier in hundreds of open access journals or in nearly 2000 established subscription journals that support open access publishing. Permitted third party re-use of these open access articles is defined by the author's choice of Creative Commons user license. See our open access license policy for more information.

**Terms & Conditions applicable to all Open Access articles published with Elsevier:**

Any reuse of the article must not represent the author as endorsing the adaptation of the article nor should the article be modified in such a way as to damage the author's honour or reputation. If any changes have been made, such changes must be clearly indicated.

The author(s) must be appropriately credited and we ask that you include the end user license and a DOI link to the formal publication on ScienceDirect.

If any part of the material to be used (for example, figures) has appeared in our publication with credit or acknowledgement to another source it is the responsibility of the user to ensure their reuse complies with the terms and conditions determined by the rights holder.

**Additional Terms & Conditions applicable to each Creative Commons user license:**

**CC BY:** The CC-BY license allows users to copy, to create extracts, abstracts and new works from the Article, to alter and revise the Article and to make commercial use of the Article (including reuse and/or resale of the Article by commercial entities), provided the user gives appropriate credit (with a link to the formal publication through the relevant DOI), provides a link to the license, indicates if changes were made and the licensor is not represented as endorsing the use made of the work. The full details of the license are available at http://creativecommons.org/licenses/by/4.0.

**CC BY NC SA:** The CC BY-NC-SA license allows users to copy, to create extracts, abstracts and new works from the Article, to alter and revise the Article, provided this is not done for commercial purposes, and that the user gives appropriate credit (with a link to the formal publication through the relevant DOI), provides a link to the license, indicates if changes were made and the licensor is not represented as endorsing the use made of the work. Further, any new works must be made available on the same conditions. The full details of the license are available at http://creativecommons.org/licenses/by-nc-sa/4.0.

**CC BY NC ND:** The CC BY-NC-ND license allows users to copy and distribute the Article, provided this is not done for commercial purposes and further does not permit distribution of the Article if it is changed or edited in any way, and provided the user gives appropriate credit (with a link to the formal publication through the relevant DOI), provides a link to the license, and that the licensor is not represented as endorsing the use made of the work. The full details of the license are available at http://creativecommons.org/licenses/by-nc-nd/4.0. Any commercial reuse of Open Access articles published with a CC BY NC SA or CC BY NC ND license requires permission from Elsevier and will be subject to a fee.

Commercial reuse includes:

- Associating advertising with the full text of the Article
- Charging fees for document delivery or access
- Article aggregation
- Systematic distribution via e-mail lists or share buttons

Posting or linking by commercial companies for use by customers of those companies.

20. **Other Conditions**:

v1.10

**You will be invoiced within 48 hours of this transaction date. You may pay your invoice by credit card upon receipt of the invoice for this transaction. Please follow instructions provided at that time.**
**To pay for this transaction now; please remit a copy of this document along with your payment. Payment should be in the form of a check or money order referencing your account number and this invoice number RLNK504003136.**

**Make payments to "COPYRIGHT CLEARANCE CENTER" and send to:**

**Copyright Clearance Center**
**29118 Network Place**
**Chicago, IL 60673-1291**
**Please disregard electronic and mailed copies if you remit payment in advance**

**Questions? or +1-855-239-3415 (toll free in the US) or +1-978-646-2777.**
