## Supplementary material for "Proof-of-principle neural network models for classification, attribution, creation, style-mixing, and morphing of image data for genetic conditions": Image permission

ELSEVIER LICENSE
TERMS AND CONDITIONS

Apr 08, 2021

---

This Agreement between Rebekah Waikel ("You") and Elsevier ("Elsevier") consists of your license details and the terms and conditions provided by Elsevier and Copyright Clearance Center.

**All payments must be made in full to CCC. For payment instructions, please see information listed at the bottom of this form.**

| | |
|---|---|
| License Number | 5041311003448 |
| License date | Apr 03, 2021 |
| Licensed Content Publisher | Elsevier |
| Licensed Content Publication | The American Journal of Medicine |
| Licensed Content Title | Have You Ever Seen a LEOPARD? |
| Licensed Content Author | Lucien Marchand |
| Licensed Content Date | Sep 1, 2019 |

| | |
|---|---|
| Licensed Content Volume | 132 |
| Licensed Content Issue | 9 |
| Licensed Content Pages | 1 |
| Start Page | e711 |
| End Page | |
| Type of Use | reuse in a journal/magazine |
| Requestor type | academic/educational institute |
| Portion | figures/tables/illustrations |
| Number of figures/tables/illustrations | 1 |
| Format | electronic |
| Are you the author of this Elsevier article? | No |
| Will you be translating? | No |
| Title of new article | A proof-of-principle neural network model for classification, |

| | |
|---|---|
| | attribution, creation, and morphing of image data for genetic conditions |
| Lead author | Dat Duong, PhD |
| Title of targeted journal | MedRxiv |
| Publisher | CSHL |
| Expected publication date | May 2021 |
| Portions | Figure B |
| Requestor Location | Dr. Rebekah Waikel<br>31 Center Dr<br><br>Bethesda, MD 20894<br>United States<br>Attn: Dr. Rebekah Waikel |
| Publisher Tax ID | 98-0397604 |
| Billing Type | Invoice |
| Billing Address | Dr. Rebekah Waikel<br>31 Center Dr<br><br>Bethesda, MD 20894<br>United States<br>Attn: Rebekah Waikel |
